# Autophagy regulator ATG7 links lipid metabolism to cell-fate decisions in kidney tubule health and disease

**DOI:** 10.1101/2025.03.26.25324675

**Authors:** Daniela Nieri, Svenja Aline Keller, Louise Pierre, Federica Carloni, Roberto Pili, Zhiyong Chen, Adia M. Ouellette, Marine Berquez, Patrick Krohn, Andrea Raimondi, Valeria Berno, Martina Zanella, Michelle Elisabeth Reid, Alaa Othman, Andrew M. Schaefer, Eric Gregory Olinger, John A. Sayer, Alina Ustiugova, Mikhail Korzinkin, Stephan C.F. Neuhauss, Christian Münz, Robert McFarland, Robert W. Taylor, Louis R. Lapierre, Elisa Araldi, Olivier Devuyst, Alessandro Luciani

## Abstract

Homeostasis in the kidney proximal tubule (PT) requires coordination between metabolism and differentiation, yet the mechanisms governing this balance remain elusive. Here, we integrate model organisms, multiomics profiling, and human genetics to identify the autophagy regulator ATG7 as a key determinant of cell-fate decisions, sustaining PT specialization in health and contributing to dysfunction in disease. In mice, PT-specific deletion of ATG7 reprograms differentiated cells into anabolic, proliferative states, impairing their specialized function and causing kidney tubulopathy. Mechanistically, loss of ATG7-dependent autophagy hinders lipid droplet clearance and restricts fatty-acid oxidation (FAO), leading to energy depletion and functional decline. In zebrafish pronephros, re-expression of wild-type ATG7 restores homeostasis in *atg7* mutants, while pharmacological FAO inhibition triggers dysfunction. In humans, *ATG7* variants associate with cardio-renal-metabolic traits and increased disease risk, whereas low *ATG7* expression correlates with transcriptional signatures of metabolic reprogramming, loss of epithelial markers, and poor prognosis in renal cell carcinoma. These findings establish a conserved genetic paradigm that links autophagy to kidney epithelial cell-fate specialization, with implications for disease, cancer, and metabolic health.

## Introduction

Specialized epithelial cells must tightly coordinate metabolic programs with differentiation cues to execute their unique physiological function and maintain homeostasis.^1^ A prototypical example of this integration is observed in the epithelial cells of the kidney’s proximal tubule (PT). These cells recover ultrafiltered plasma proteins and solutes via receptor-mediated endocytosis — capturing low-molecular-weight ligands through LRP2/megalin and CUBN/cubilin — and transport them to lysosomes for degradation, while receptors are recycled back to the membrane.^2–4^

This constant cycle of endocytosis, intracellular trafficking, and processing imposes a significant energetic burden, demanding sustained adenosine triphosphate (ATP) production and energy consumption. Dysregulation of these processes — whether due to congenital or acquired defects — leads to PT dysfunction, triggering the appearance in the urine of essential nutrients and life-threatening metabolic complications.^2,3^

Simultaneously, macroautophagy (hereafter referred to as autophagy) facilitates the sequestration and lysosomal degradation of intracellular macromolecules and organelles, thereby supporting energy metabolism and maintaining cellular activities. While defects in either autophagy or lysosomal function disrupt PT differentiation and reabsorptive capacity, they operate at distinct steps: lysosomal dysfunction hinders cargo degradation, whereas the loss of core autophagy regulators prevents autophagosome formation or maturation.

A central mediator of autophagosome biogenesis is ATG7, a conserved E1-like enzyme that facilitates conjugation of ATG12 to ATG5 and lipidation of MAP1LC3-I to MAP1LC3-II — an established marker of autophagic flux in cells and tissues.^8–11^ Global *Atg7* knockout in mice results in neonatal lethality and metabolic collapse,^12–13^ while PT-specific deletion overwhelms adaptive stress responses and promotes macromolecular damage and dysfunction.^14,15^ Although ATG7-dependent autophagy is well-recognised for its role in cellular stress resilience and repair mechanisms, its contribution to cell-fate decisions and specialization during the differentiation of the kidney tubule remains largely unexplored.

The kidney is among the most metabolically active organs, requiring continuous energy production to sustain ATP-dependent processes such as reabsorption and transport of nutrients and multiple solutes.^16^ Tubular epithelial cells are densely packed with mitochondria, reflecting their reliance on fatty-acid β-oxidation (FAO) and oxidative phosphorylation to meet high energetic demands.^17,18^ Disruption of mitochondrial homeostasis — characterized by reduced electron-transport chain activity, reduced FAO, altered oxidative phosphorylation and diminished ATP production, and increased reactive oxygen species (ROS) — compromises cellular bioenergetics, PT differentiation and function, contributing to the onset and progression of various kidney diseases.^6,7,19^

In preclinical models, both genetic and pharmacological activation of autophagy improves mitochondrial quality control, restores FA metabolism and energy production, and mitigates dysfunction and the progression of kidney disease.^18,20^ In humans, biallelic pathogenic variants in *ATG7* lead to profound autophagy impairment, accompanied by neurodevelopmental defects and systemic metabolic dysregulation, underscoring its essential role in human health.^21^ Together, these findings raise the possibility that ATG7-mediated autophagy may modulate the metabolic cues governing kidney epithelial cell-fate specification and state transitions in health and disease.

Here, we used genetically engineered model organisms with physiologically relevant epithelial PT cellular systems and combine these approaches with lipid metabolism assays, multiomics profiles, and phenome-wide genetic analysis across multiple populations to investigate how the autophagy regulator ATG7 links lipid homeostasis to energy metabolism to direct epithelial cell-fate specialization during the differentiation of the kidney tubule. We demonstrate that dysregulation of the ATG7-driven autophagy–metabolism axis underlies a spectrum of kidney diseases and influences the progression of clear cell renal cell carcinoma, thus offering insights into mechanisms of homeostasis and highlighting new avenues for therapeutic intervention.

## Results

### ATG7 deficiency impairs autophagy in kidney PT epithelial cells

To explore the role of ATG7 in PT autophagy (Supplementary Fig.1a), we crossed mice^13^ homozygous for floxed alleles of *Atg7* with *Ggt1-Cre* mice^22^ expressing Cre recombinase under the control of the rat *Ggt1*, gamma-glutamyltransferase 1, gene promoter (Supplementary Fig.1b-1e).^22^ This strategy generated *Atg7*^fl/fl^; γ*Gt1*^cre/cre^ mice (hereafter referred to as *Atg7*^PT-KO^), in which exon 14 of *Atg7* was efficiently excised (Supplementary Fig. 1f-1g), and corresponding control littermates (*Atg7*^fl/fl^; γ*Gt1*^wt/wt^, hereafter CTR).

Conditional *Atg7*^PT-KO^ mice were viable with expected Mendelian frequencies and gender ratios. As expected, the expression of Atg7 was significantly reduced at the mRNA and protein levels (Supplementary Fig. 2a-2b). In line, *Atg7*^PT-KO^ kidneys exhibited markedly reduced levels of the lipidated, autophagosome-associated form of Map1lc3b (Map1lc3b-II; Supplementary Fig. 2c) and accumulation of Sequestosome 1 (Sqstm1; ref.11; Supplementary Fig. 2c-2d), indicating autophagy impairment in PT epithelial cells.

### Defective ATG7-driven autophagy triggers dyshomeostasis and dysfunction

To examine how loss of ATG7-driven autophagy deficiency affects PT homeostasis and normal physiology, we longitudinally tracked a cohort of *Atg7*^PT-KO^ mice and CTR littermates up to 24 weeks of age (Fig. 1a; Table S1). Beginning at 12 weeks of age, *Atg7*^PT-KO^ mice displayed reduced body weight and impaired growth compared to CTR littermates (Fig. 1b-1c). Urinary levels of the LMW Clara cell secretory protein 16 (CC16), a PT dysfunction marker, were significantly increased in *Atg7*^PT-KO^ compared to CTR mice from 4 weeks of age, irrespective of gender (Fig. 1d; Supplementary Fig. 3a). Other indicators of PT dysfunction including albumin, transferrin (TF) and vitamin D binding protein (VDBP) (Fig. 1e-1f), along with abnormal loss of glucose, phosphate, calcium, and amino acids, were observed in *Atg7*^PT-KO^ mice compared to CTR littermates (Fig. 1g-1i; Supplementary Fig. 3b- 3c; Table S1). Remarkably, despite clear PT functional deficits, *Atg7*^PT-KO^ kidneys exhibited no evidence of fibrosis, apoptosis, or overt structural abnormalities (Supplementary Fig. 3d-3h), and blood urea nitrogen (BUN), plasma creatinine, and survival rates remained comparable to controls (Supplementary Fig. 3i-3k; Table S2). Together, these findings establish that ATG7-dependent autophagy is critical for sustaining PT function and homeostasis, and that its disruption leads to a global proximal tubulopathy in the absence of structural kidney damage.

**Fig. 1.**
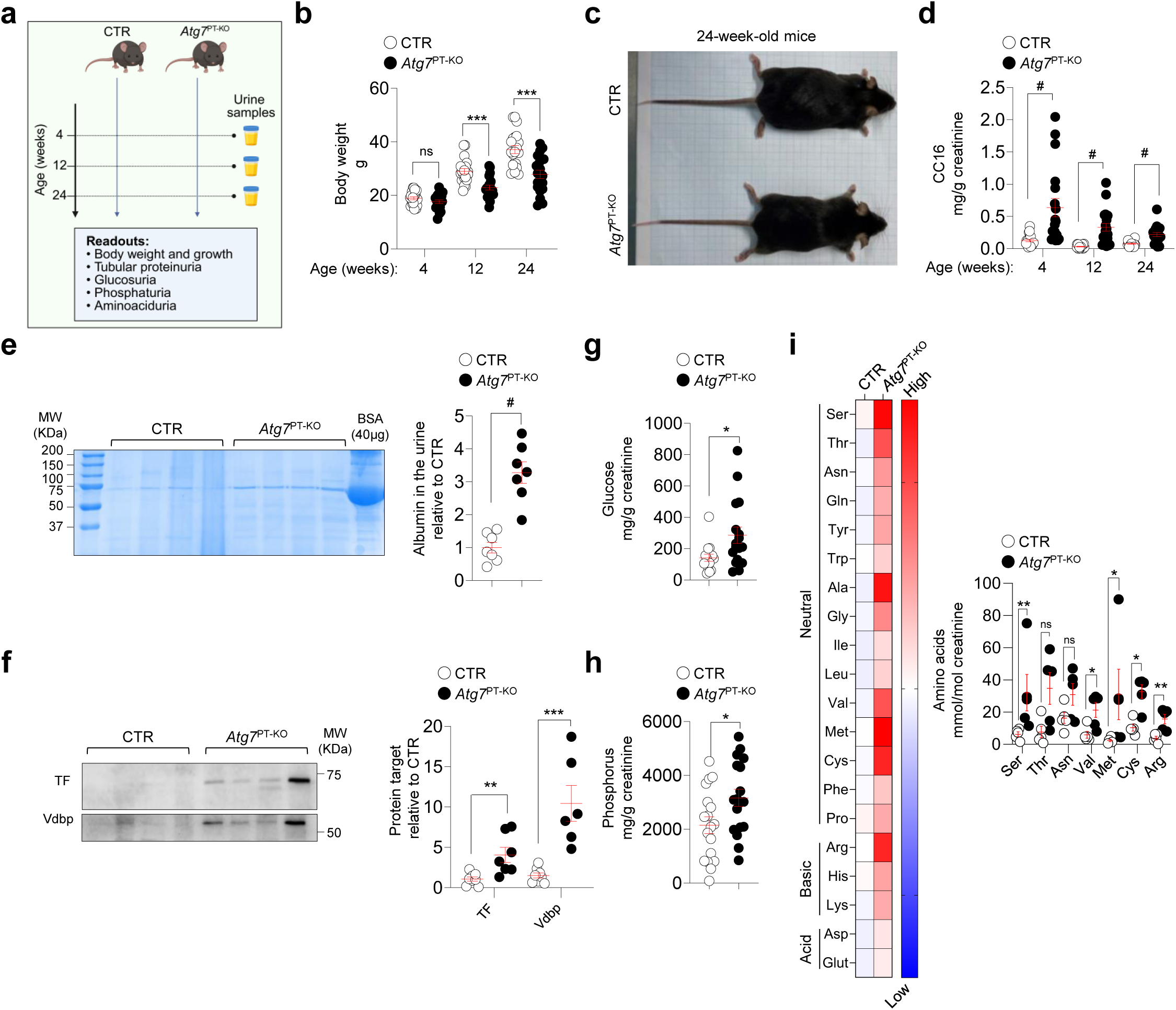
Loss of ATG7 in the adult mouse kidneys triggers growth defects, metabolic dyshomeostasis and proximal tubulopathy. (**a**) Schematic of the experimental workflow. (**b**) Body weight measurements of mice at 4, 12, and 24 weeks, n=20 mice per genotype. (**c**) Representative images of *Atg7* mice at 24 weeks. (**d**) Urinary excretion levels of the low-molecular-weight CC16 protein at the indicated times. At 4 weeks of age: n=19 CTR and n=20 *Atg7*^PT-KO^ mice; at 12 and 24 weeks of age: n=20 mice per genotype. (**e**) Quantification of urinary albumin in 24-week-old mice; nL=L8 mice per genotype. (**f**) Representative immunoblots and quantification of the indicated proteins in 24-week-old mice, n=6 animals per genotype. (**g**-**h**) Quantification of urinary (**g**) glucose and (**h**) phosphorus in 24-week-old *Atg7* mice. For (**g**), n=15 CTR and 17 *Atg7*^PT-KO^ mice; for (**h**), n=19 mice per genotype. (**i**) Quantification of urinary amino acid levels in 24-week-old mice, n=5 animals per genotype. Ala, alanine; Arg, arginine; Asn, asparagine; Asp, aspartate; Cys, cysteine; Gln, glutamine; Glu, glutamate; Gly, glycine; His, histidine; Hyp, hydroxyproline; Ile, isoleucine; Leu, leucine; Lys, lysine; Met, methionine; Orn, ornithine; Phe, phenylalanine; Pro, proline; Ser, serine; Thr, threonine; Trp, tryptophan; Tyr, tyrosine; Val, valine. All urinary parameters were normalised to creatinine concentration. Bar graphs show mean ± SEM. Statistics were calculated using unpaired two-tailed Student’s *t*-test, *P < 0.05, **P < 0.01, ***P < 0.001, and ^#^P < 0.0001 relative to CTR. NS, not significant.

### ATG7 deficiency diverts PT towards abnormal growth and proliferation

To assess how ATG7 deficiency alters PT cell identity and phenotype, we examined endocytic capacity, differentiation, and proliferation markers in *Atg7*^PT-KO^ versus control mice (Fig. 2a). Fluorescent tracer studies showed a marked impairment in both receptor-mediated (Cy5-lactoglobulin) and fluid-phase (Alexa647-dextran) endocytosis in *Atg7*^PT-KO^ PTs (Fig. 2b). Consistently, mRNA levels of key solute transporters (*Slc34a2*, *Slc5a2*) and endocytic receptors (*Lrp2*, *Cubn*) were significantly reduced (Fig. 2c), and apical localization of LRP2/megalin and transferrin receptor was diminished (Fig. 2d; Supplementary Fig. 4a), despite preserved expression of core endosomal–lysosomal components (Supplementary Fig. 4b). In parallel, *Atg7*^PT-KO^ PTs upregulated genes associated with cell cycle and division (*Ccna2*, *Ccnb2*, *Cdc20*, *Pcna*, *Ki67*; Fig. 2e), increased proliferation rates (Fig. 2f), accompanied by an increased kidney-to-body-weight ratio (Supplementary Fig. 4c).

**Fig. 2.**
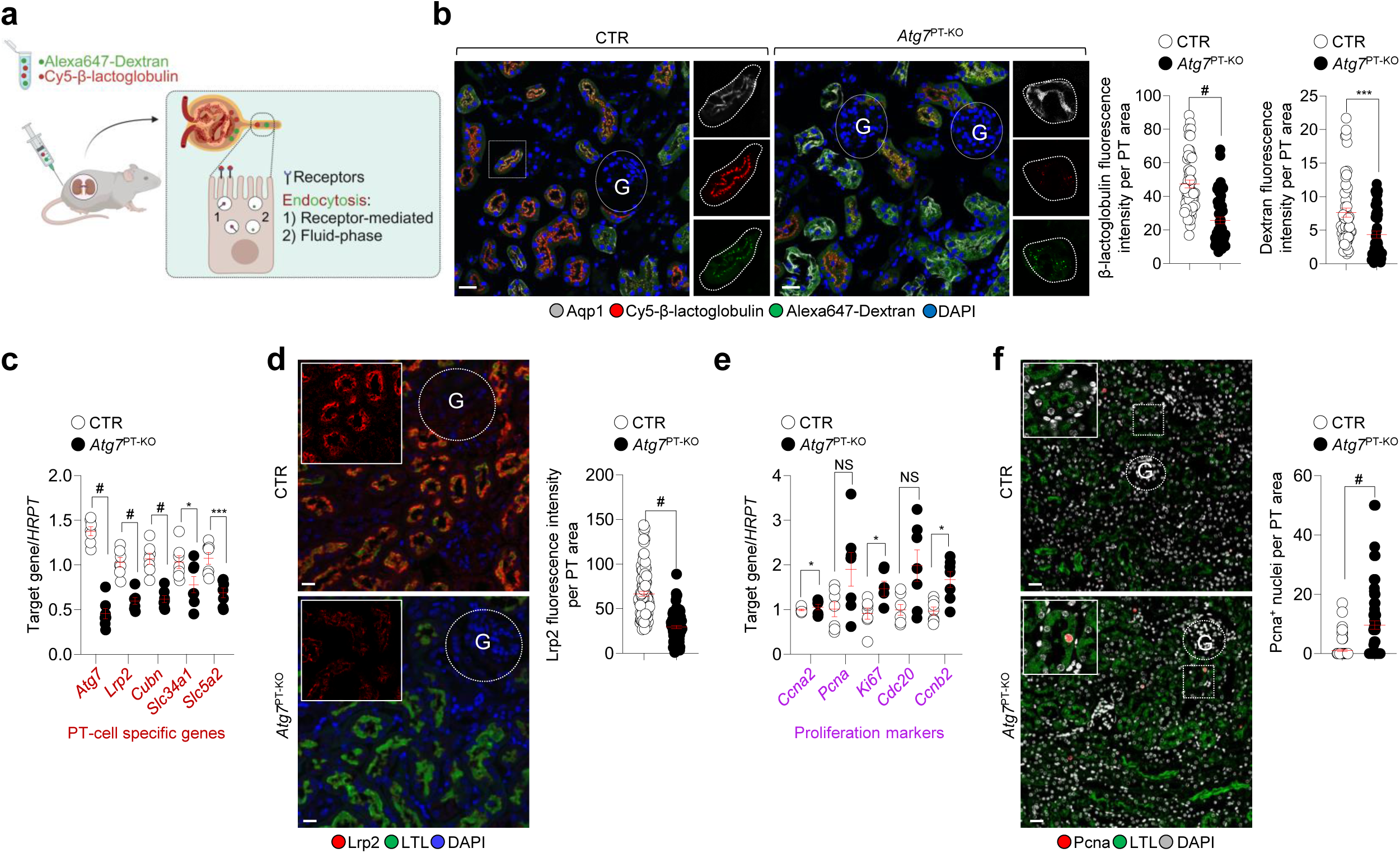
Loss of ATG7 diverts the functioning states of PT cells towards abnormal growth and proliferation. (**a**) Schematic of the experimental workflow used to assess endocytosis. (**b**) Confocal imaging and quantification of Cy5-(red) or Alexa 647-Dextran (green) fluorescence intensity in Aqp1^+^ PT cells (grey) of the mouse kidneys; n = 60 PT segments pooled from 2 mice per genotype. (**c**) mRNA expression of the indicated genes in the kidney cortex of 24-week-old mice, n=7 animals per genotype. (**d**) Confocal imaging and quantification of Lrp2 (red) fluorescence intensity in the LTL^+^ (green) PT of mouse kidneys; n = 100 PT segments pooled from 2 mice per genotype. (**e**) mRNA expression of the indicated genes in the kidney cortex of 24-week-old mice, n=7 animals per genotype. (**f**) Confocal microscopy and quantification of the percentage of PCNA^+^ (red) nuclei in LTL^+^ (green) PT segments of the mouse kidneys; n=90 PT segments pooled from 2 animals per genotype. Nuclei counterstained with DAPI (blue or grey). Bar graphs show mean ± SEM. Statistics were calculated using unpaired two-tailed Student’s *t*-test, *P < 0.05, ***P < 0.001, and ^#^P < 0.0001 relative to CTR. NS, not significant. Scale bars, 25µm.

Morphologically, PT epithelial cells transitioned from a cuboidal to a columnar phenotype (Supplementary Fig. 4d), even though transcription factors that normally couple metabolism to differentiation (*Esrra*, *Hnf1a*, *Hnf4a*, *Ppara*^19,24^) remained unchanged (Supplementary Fig. 4e-4f). Together, these findings reveal that ATG7-dependent autophagy preserves the specialized, endocytic phenotype of PT cells. Loss of ATG7 triggers a shift toward anabolic and proliferative states, disrupting epithelial identity and impairing receptor-mediated endocytosis.

### ATG7 deficiency alters autophagy, metabolism, and function in cultured PT cells

To delineate how ATG7 loss disrupts PT-cell phenotype and homeostasis, we analyzed primary mouse PT cells (mPTCs^6,7^) isolated from microdissected PT segments of *Atg7*^PT-KO^ and control mouse kidneys (Fig. 3a). Compared to control cells, *Atg7*^PT-KO^ mPTCs exhibited impaired conversion of Map1lc3b-I to its lipidated form, Map1lc3b-II, and reduced formation of Map1lc3b-positive autophagosomes under both fed and starved conditions (Fig. 3b-3f). However, treatment with the lysosomal inhibitor Bafilomycin A1 (BafA1) increased Map1lc3b-II levels in both starved control and ATG7-deficient cells (Fig. 3c-3e), suggesting activation of alternative autophagy pathways in response to nutrient/lysosomal stress.^12^ In line, transmission electron microscopy revealed a lower abundance of autophagic vacuoles in BafA1-treated, starved *Atg7*^PT-KO^ mPTCs (Fig. 3g). These autophagic defects were accompanied by altered lysosomal dynamics and catabolic function, dysregulated anabolic programs for cell growth and proliferation, and impaired receptor-mediated uptake of fluorescent ligands — phenotypes that mirror the in vivo observations (Fig. 3h-3j; Supplementary Fig. 5a-5b). These cellular abnormalities occurred independently of changes in the expression of other core autophagy genes (Supplementary Fig. 5c) or major nutrient-sensing pathways, including mTORC1 and AMPK (Supplementary Fig. 5d-5e).^25^ Together, these data establish that ATG7-driven autophagy is indispensable for maintaining metabolic trajectories of PT cells and their specialized function, acting through mechanisms that are at least partially independent of canonical nutrient-sensing and signalling pathways.

**Fig. 3.**
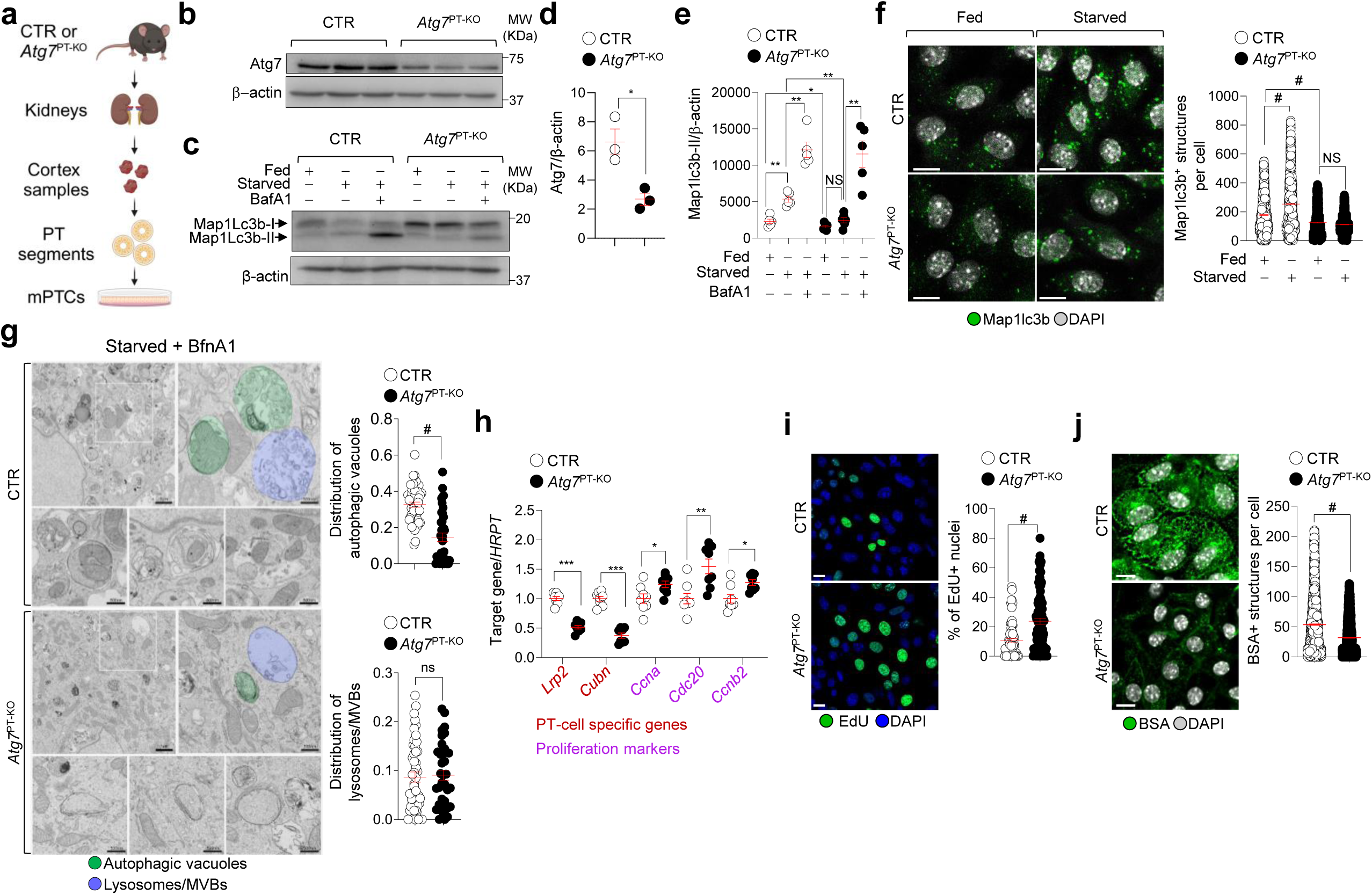
ATG7 deficiency impairs autophagy, leading to abnormal proliferation and loss of epithelial differentiation in cultured PT cells. (**a**) Schematic of the workflow for isolating and culturing primary cells derived from PT segments of the mouse kidneys (mPTCs). (**b,d**) Immunoblots (**b**) and quantification (**d**) of the indicated proteins in PTCs from 24-week-old mouse kidneys, n=3 biologically independent experiments. (**c,e-g**) mPTCs were cultured under fed or starved conditions, or starved conditions in the presence of BafA1 (250 nM for 4 h). (**c,e**) Immunoblots (**c**) and quantification (**e**) of the indicated proteins, n=6 biologically independent experiments. (**f**) Confocal imaging and quantification of the number of Map1lc3^+^ structures. Fed conditions: n=1570 CTR and 1413 *Atg7*^PT-KO^ cells. Starved conditions: n=910 CTR and 1548 *Atg7*^PT-KO^ cells, pooled from 3 biologically independent experiments. (**g**) Representative electron micrographs and quantification of cytoplasm area occupied by autophagy vacuoles or lysosome/multivesicular bodies (expressed as the percentage of the total area). AVs distribution: n = 48 CTR and 40 *Atg7*^PT-KO^ cells pooled from 2 biologically independent experiments. Lysosomes/MVBs distribution: n = 48 CTR and 38 *Atg7*^PT-KO^ cells pooled from 2 biologically independent experiments. Green and purple indicate EM-compatible AVs or Lys/MVBs, respectively. (**h**) mRNA expression of the indicated genes in mPTCs from 24-week-old *Atg7* mice, n = 8 biologically independent experiments. (**i**) Confocal imaging and quantification of EdU^+^ cells (expressed as a percentage of total cells); n = 84 CTR and 95 *Atg7*^PT-KO^ cells from 3 biologically independent experiments. (**j**) Confocal microscopy and quantification of BSA+ structures per cell; n = 2096 CTR and 1833 *Atg7*^PT-KO^ cells from 3 biologically independent experiments. Nuclei counterstained with DAPI (blue or grey). Bar graphs show mean ± SEM. Statistics were calculated by unpaired two-tailed Student’s t-test *P < 0.05, **P < 0.01, ***P < 0.001, ^#^P < 0.0001 relative to fed CTR or starved CTR or starved *Atg7*^PT-KO^. NS not significant. Scale bars, 25µm in (**f**), (**i**) and (**j**).

### Loss of ATG7-driven autophagy impairs lipid droplet turnover, triggering lipid storage

To uncover factors driving loss of epithelial cell identity and dysregulated homeostasis downstream of ATG7/autophagy deficiency, we performed proteomic and metabolomic profiling of control and *Atg7*-deficient mPTCs (Supplementary Fig. 6a-6d). Protein-protein interaction (PPI) network analysis of differentially expressed proteins revealed significant enrichment in mitochondria and metabolic pathways, including amino acid and fatty acid (FA) metabolism, the TCA cycle, electron transport, FA oxidation, and energy generation in *Atg7*^PT-KO^ mPTCs (Supplementary Fig. 6e) compared to CTR cells. Combined clustering of metabolites and proteins further highlighted enrichment in FA elongation/degradation, amino acid metabolism, autophagy–lysosome degradation systems, and signatures associated with senescence and cancer metabolism in *Atg7*-deficient mPTCs (Fig. 4a). Based on these findings, we hypothesized that ATG7 loss disrupts PT-cell phenotype and homeostasis by impairing the autophagic degradation of lipid droplets (LDs) — a conserved process known as lipophagy.^26^ In control mPTCs, fasting protocols triggered a transient increase in LDs at 4–8 hours that normalized by 16 hours, consistent with LD turnover via autophagy, as shown by BODIPY^493/503^ staining (Supplementary Fig. 7a). A similar LD clearance pattern was observed following oleic acid (OA) treatment, which promotes triacylglycerol (TAG) accumulation within LDs^27^ (Supplementary Fig. 7b). Blocking catabolic autophagy with BafA1 or the VPS34 inhibitor SAR405 impaired LD clearance and increased LD size (Supplementary Fig. 7c-7e). Treatment with BafA1 increased colocalization of LD markers (Plin2, LipidTOX) with the lysosomal marker LAMP1, while SAR405 reduced it (Supplementary Fig. 7c,7f-7g), underscoring the reliance of LD turnover and homeostasis on autophagy and lysosome-related pathways.^26^ In contrast, when compared to control cells, *Atg7*-deficient mPTCs exhibited elevated baseline LD levels that failed to decrease during starvation, irrespective of BafA1 (Fig. 4b). These cells also accumulated Sqstm1⁺ LDs and showed reduced colocalization between LipidTOX and LAMP1 (Fig. 4b; Supplementary Fig. 8a-8b), indicating defective lipophagy. Lipidomic profiling confirmed the accumulation of TAGs and phosphatidylcholines (PCs), as well as altered levels of lysophosphatidylethanolamines (LPEs) and sphingomyelins (SMs) in *Atg7*^PT-KO^ cells (Fig. 4c; Supplementary Fig. 9a-9d). Notably, expression of FA transporters, lipolytic enzymes, mitochondrial/peroxisomal FAO genes, and mitochondrial biogenesis markers remained unchanged (Supplementary Fig. 10a-10c), suggesting that defects in lipid homeostasis stem specifically from impaired lipophagy. Together, these findings demonstrate that ATG7 is required for lipid droplet turnover and metabolic homeostasis in PT cells through a lipophagy-dependent mechanism.

**Fig. 4.**
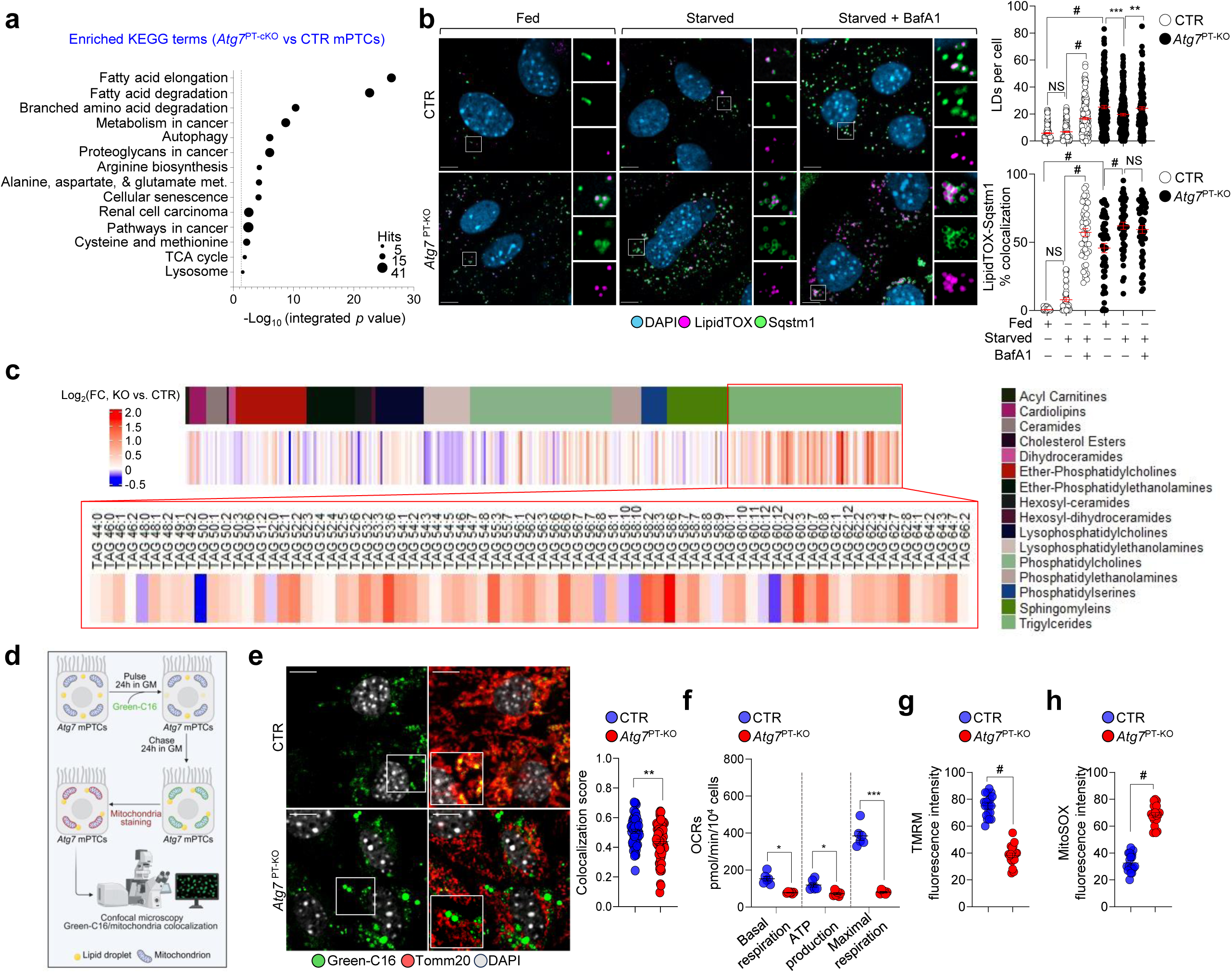
Loss of ATG7 impairs the autophagic lipid degradation, compromising FA metabolism and energy production in PT cells. (**a**) Multiomics integration and KEGG pathway enrichment analysis. The black dashed line marks the threshold for significance. Black circles indicate the enriched proteins and metabolites in *Atg7* mPTCs compared to CTR, n=4 biologically independent experiments. *P* values were calculated using Omicsnet2.0. (**b**) Confocal imaging and quantification of the number of LDs (LipidTOX, magenta) and LipidTOX/Sqstm1 (green) colocalization (%). Number: n = 158 fed, 242 starved, and 200 BafA1-treated, starved CTR cells; and n = 212 fed, 228 starved, and 208 BafA1-treated, starved *Atg7*^PT-KO^ cells. Colocalization (%): n = 42 fed, 43 starved, 50 BafA1-treated, starved CTR cells; and n = 51 fed, 50 starved, 51 BafA1-treated, starved *Atg7*^PT-KO^ cells. (**c**) Lipidomics and heat map showing lipid changes in *Atg7*^KO^ mPTCs vs. control cells. (**d**) Schematic of the experimental workflow for tracking FA (Green-C16) transport into the mitochondria. (**e**) Confocal imaging and quantification of Bodipy-C16-TOMM20^+^ colocalization score (n=15 fields from 3 biologically independent experiments). Insets show high-magnification views. (**f**) Oxygen consumption rates (OCRs) of basal respiration, mitochondrial ATP production and maximal respiration; n = 9 replicates from 3 biologically independent experiments. (**g, h**) Quantification of TMRM or MitoSOX fluorescence intensity; nL=L20 cells from 2 biologically independent experiments. Representative images shown in **Supplementary Fig.10d-10e**. Bar graphs show mean ± SEM. Statistics were calculated by one-way ANOVA followed by Tukey’s multiple comparisons test in (**b**) and by unpaired two-tailed Student’s t-test in **(e, f, g, h)** *P < 0.05, **P < 0.01, ***P < 0.001, ^#^P < 0.0001 relative to fed/starved CTR or fed/starved *Atg7*^PT-KO^ cells. NS, not significant Nuclei counterstained with DAPI (blue or grey). Scale bars, 10µm.

### ATG7-driven autophagy couples lipid metabolism to mitochondrial energy production

Lipid catabolism releases fatty acids (FAs) that fuel mitochondrial β-oxidation and ATP production.^26^ To test whether ATG7-mediated autophagy governs this process, we tracked the intracellular fate of a fluorescent FA analogue, BODIPY C16, in *Atg7*^PT-KO^ and control mPTCs (Fig. 4d). Following a 24-hour pulse and 24-hour chase, control cells efficiently mobilized Green-C16 from LDs to mitochondria, while the tracer remained trapped in LDs in *Atg7*^PT-KO^ cells (Fig. 4e), indicating impaired FA release and reduced mitochondrial availability. Consistently, seahorse-based metabolic profiling revealed reduced basal respiration, mitochondrial ATP-linked respiration, and maximal oxygen consumption rates (OCRs) in mutant cells (Fig. 4f), accompanied by decreased mitochondrial membrane potential (Fig. 4g; Supplementary Fig. 10d) and elevated mitochondrial reactive oxygen species (ROS; Fig. 4h; Supplementary Fig. 10e), despite unchanged levels of mitochondrial proteins (Supplementary Fig. 10f). To test whether pharmacological enhancement of lipid metabolism could restore PT cell-fate specialization and differentiation, we treated *Atg7*-deficient cells with fenofibrate, a PPARα agonist known to enhance cytosolic lipolysis and FAO pathways.^19^ Fenofibrate induced expression of *Pnpla2*, *Lipe*, *Mgll*, *Cpt1*, and *Cpt2* (Supplementary Fig. 11a), rescued LD homeostasis without reversing autophagy defects (Supplementary Fig. 11b-c), suppressed aberrant proliferation and rescued PT-specific gene expression, differentiation markers, and endocytic uptake in *Atg7*^PT-KO^ cells (Supplementary Fig. 11d-11h). Conversely, inhibition of mitochondrial FA import using etomoxir^27,28^ in control cells phenocopied ATG7 deficiency — leading to impaired ligand uptake and LD accumulation (Supplementary Fig. 11h-11i). Together, these findings demonstrate that ATG7-dependent autophagy is essential for mobilizing LD-derived FAs to mitochondria, sustaining energy metabolism, and hence the specialized function of PT cells and homeostasis. Importantly, enhancing FA metabolism through PPARα activation can compensate for autophagy/lipophagy failure and restore core aspects of normal PT physiology.

### Cell-autonomous role of ATG7 in maintaining PT metabolism and differentiation

To confirm the cell-autonomous role of ATG7, we cultured primary cells derived from microdissected PT segments of *Atg7*^fl/fl^ kidneys and acutely deleted *Atg7* using adenoviral Cre-recombinase. This intervention effectively reduced *Atg7* mRNA and protein levels (Fig. 5a and Supplementary Fig. 12a) and impaired autophagy, as shown by decreased Map1lc3b-I to Map1lc3b-II conversion and reduced autophagosome formation under both fed and starved conditions (Fig. 5b; Supplementary Fig. 12b). Consistent with in vivo observations, *Atg7*-deleted mPTCs accumulated lipid droplets (LDs) that failed to clear upon starvation or Bafilomycin A1 treatment, indicating defective lipophagy (Fig. 5c). Following oleic acid overload and starvation in the presence and absence of BafA1(Fig. 5d), *Atg7*-deleted cells exhibited impaired autophagic delivery of LDs to lysosomes for degradation, evidenced by increased Plin2–Sqstm1 colocalization and reduced LipidTOX–LAMP1 colocalization (Fig. 5e). Mitochondrial morphology and bioenergetic function were compromised in these mutant cells (Fig. 5f; Supplementary Fig. 12c), accompanied by augmented proliferation (Fig. 5g), loss of PT-specific gene expression, enhanced dedifferentiation (Fig. 5h and Supplementary Fig. 12d), and impaired endocytic uptake (Fig. 5i; Supplementary Fig. 12e-12f). Deletion of *Atg5* phenocopied these defects (Supplementary Fig. 13a-13e), and loss of Fip200 — required for canonical autophagy but not for alternative trafficking pathways, such as LC3-associated phagocytosis (LAP^29^) and endocytosis (LANDO^30^) — similarly reduced ligand uptake (Supplementary Fig. 13a-13c, 13f-13h). Together, these findings demonstrate that ATG7-dependent autophagy acts in a cell-autonomous manner to coordinate metabolic fate, differentiation of PT cells, and their functional specialization in the kidney tubule.

**Fig. 5.**
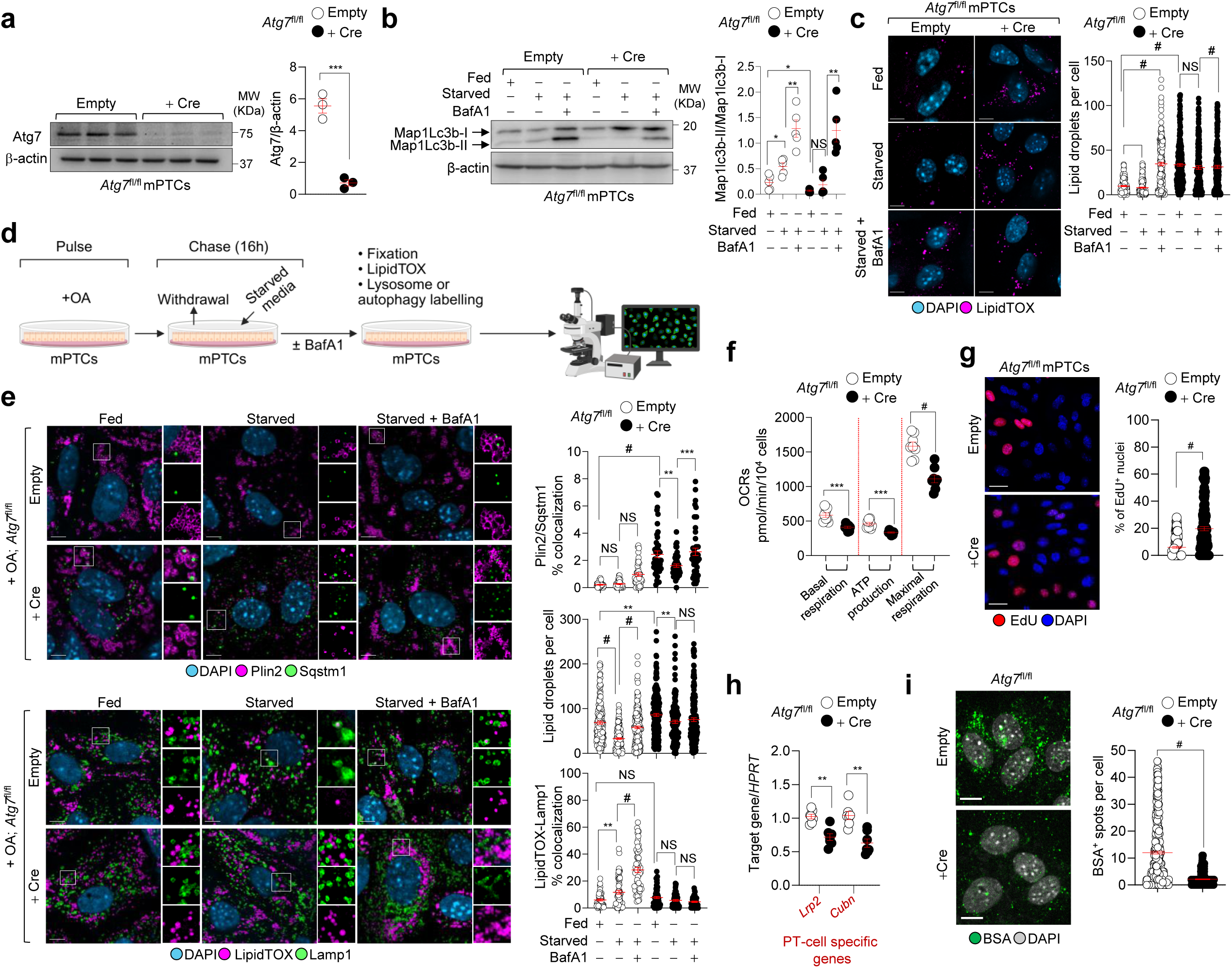
Cell-autonomous role of ATG7 in PT metabolism and differentiation. (**a-b**) Quantification of the indicated proteins transduced with empty vector or Cre recombinase; n = 3 (**a**) and n = 5 (**b**) biologically independent experiments. (**c**) Confocal imaging and quantification of the number of LDs (LipidTOX, magenta). Empty: n = 167 fed, 223 starved , 220 BafA1-treated, starved cells; + Cre: n = 287 fed, 210 starved, and 242 BafA1-treated, starved cells. (**d**) Schematic of the experimental workflow used to assess autophagic degradation of LDs. (**e**) Confocal imaging and quantification of LD number (LipidTOX, magenta), Plin2 (magenta)-Sqstm1 (green), and LipidTOX/Lamp1 (green) colocalization (%). Plin2/Sqstm1 colocalization: Empty: n = 39 fed, 48 starved, 43 BafA1-treated, starved cells; + Cre: n = 45 fed or starved, 48 BafA1-treated, starved cells. Number: Empty: n = 180 fed, 169 starved, and 198 BafA1-treated, starved cells; + Cre: 225 fed and 210 starved, and 242 BafA1-treated, starved cells. LipidTOX/Lamp1 colocalization: Empty: n = 56 fed, 60 starved, and 61 BafA1-treated, starved cells; + Cre: n= 64 fed, 52 starved, and 53 BafA1-treated cells. (**f**) Oxygen consumption rates (OCRs) of basal respiration, mitochondrial ATP production and maximal respiration; n = 8 replicates from 2 biologically independent experiments. (**g**) Confocal imaging and quantification of EdU^+^ cells; n = 133 (Empty) and 130 (+ Cre) cells from 3 biologically independent experiments. (**h**) mRNA expression of the indicated genes, n = 7 biologically independent experiments. (**i**) Confocal imaging and quantification of BSA^+^ structures per cell; n = 466 (Empty) and 387 cells (+ Cre) from 4 biologically independent experiments. Nuclei counterstained with DAPI (blue). Plots show mean ± SEM. Statistics were calculated by one-way ANOVA followed by Tukey’s multiple comparisons test in (**b**,**c**,**e**) and unpaired two-tailed Student’s *t*-test in (**a**,**f-i**), *P < 0.05, **P < 0.01, ***P < 0.001, ^#^P < 0.0001 relative to fed/starved cells transduced with Empty or with Cre. NS, not significant. Scale bars, 10µm.

### Metabolic and functional conservation of ATG7 in nematodes and zebrafish

To evaluate the evolutionary conservation of ATG7 function, we examined autophagy and epithelial homeostasis in *Caenorhabditis elegans* and *Danio rerio* (zebrafish). In *C. elegans*, we employed two established reporters^31^: SQST-1::RFP to monitor autophagy flux and DHS-3::GFP to visualize intestinal LDs (Fig. 6a). *atg-7* mutants showed increased SQST-1 accumulation (Fig. 6b), elevated intestinal LD content (Fig. 6c), and impaired mitochondrial bioenergetics, as reflected by reduced whole-organism oxygen consumption (Fig. 6d).

**Fig. 6.**
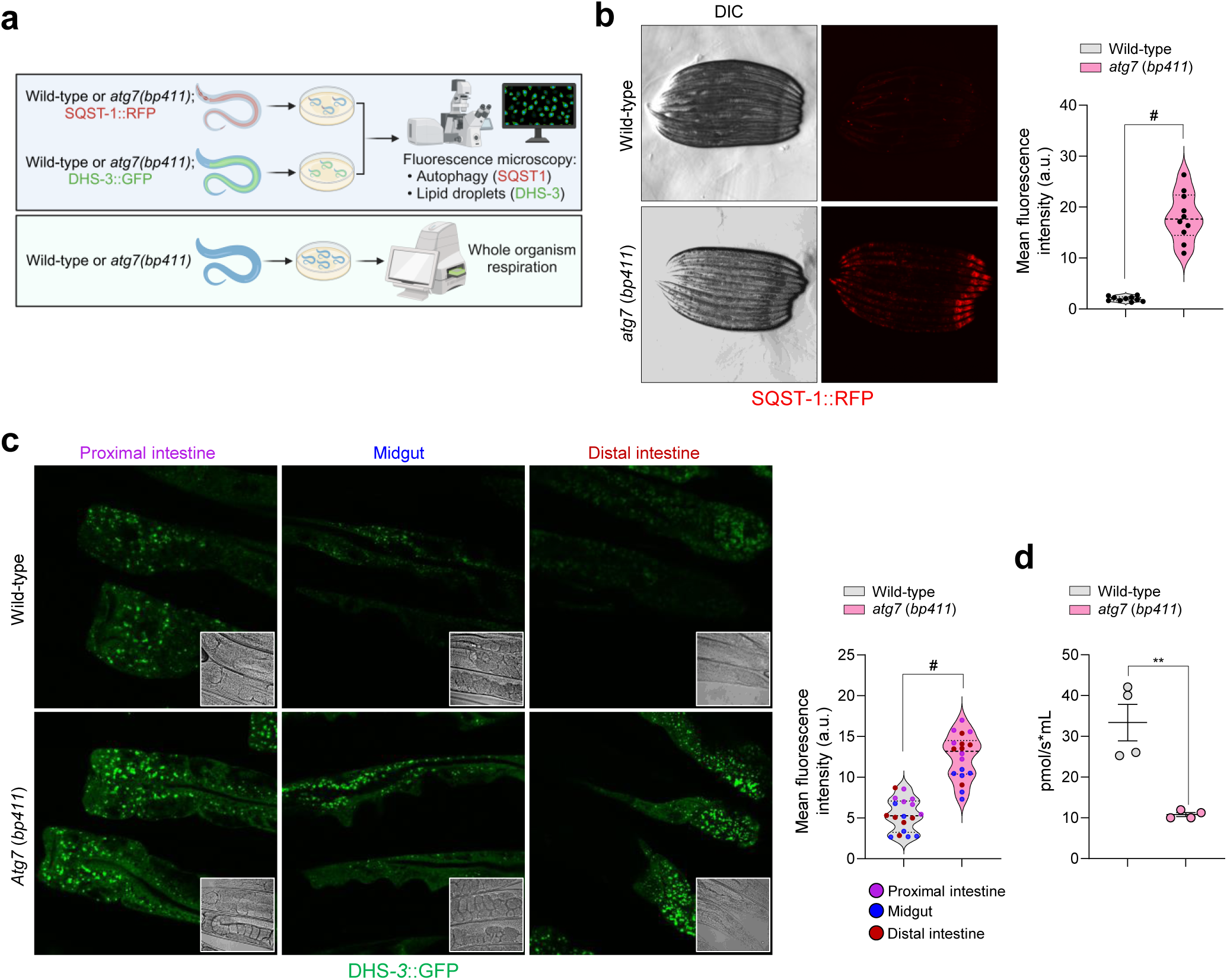
Autophagy regulator atg7 has a conserved role in *C. elegans*. (**a**) Schematic of the experimental workflow used to measure autophagy, LDs, and whole-organism respiration in nematodes. (**b**) Light microscopy and quantification of RFP fluorescence intensity in wild-type and *atg7 (bp411)* mutant nematodes, n = 10 animals per group. (**c**) Light microscopy and quantification of GFP fluorescence intensity in the intestine of both WT and *atg7 (bp411)* mutant nematodes, n = 6 animals per group. (**d**) Oxygen consumption analysis in wild-type and *atg7(bp411)* nematodes at day 4 of adulthood, normalised to the number of nematodes, n = 4 biologically independent experiments. Bar graphs show mean ± SEM. Statistics were calculated by unpaired two-tailed Student’s t-test *P < 0.05, **P < 0.01, ***P < 0.001, ^#^P < 0.0001 relative to wild-type nematodes.

The zebrafish pronephros mimics the mammalian kidney PT in structure and function, including reliance on endolysosomal trafficking, FAO, and receptor-mediated endocytosis.^32^ We generated whole-body *atg7*-null zebrafish using CRISPR/Cas9 editing (Supplementary Fig. 14a-14c). Although early development was unaffected (Supplementary Fig. 14d), *atg7*^KO^ larvae showed reduced motility from 14 days post-fertilization (dpf) and increased mortality beyond 20 dpf (Fig. 7a-7c). To visualize autophagy in vivo, we generated a transgenic zebrafish line expressing the autophagosome marker mCherry-MAP1LC3B (Fig. 7d). In *atg7*-deficient larvae, autophagosome formation in the pronephros was significantly reduced (Fig. 7e; Supplementary Fig. 14e), a defect confirmed by electron microscopy showing aberrant phagophore structures and reduced autophagic vacuoles (Supplementary Fig. 14f). To assess pronephros function, we utilized a low-molecular-weight proteinuria biosensor line (lfabp10a::½vdbp-mCherry).^33^ At 14 dpf, *atg7*^KO^ larvae exhibited elevated urinary mCherry levels, indicative of tubular dysfunction and impaired reabsorption (Fig. 7g). Expression of wild-type human ATG7 in *atg7*-deficient zebrafish rescued both pronephros function and LMW proteinuria (Fig. 7h; Supplementary Fig. 14g-14h). Furthermore, pharmacological inhibition of mitochondrial FAO with non-toxic doses of etomoxir in wild-type larvae induced swimming defects and tubular proteinuria (Fig. 7i-7j), phenocopying *atg7* deficiency. Together, these findings highlight an evolutionarily conserved role of ATG7-driven autophagy in sustaining metabolic homeostasis and functions of PT cells.

**Fig. 7.**
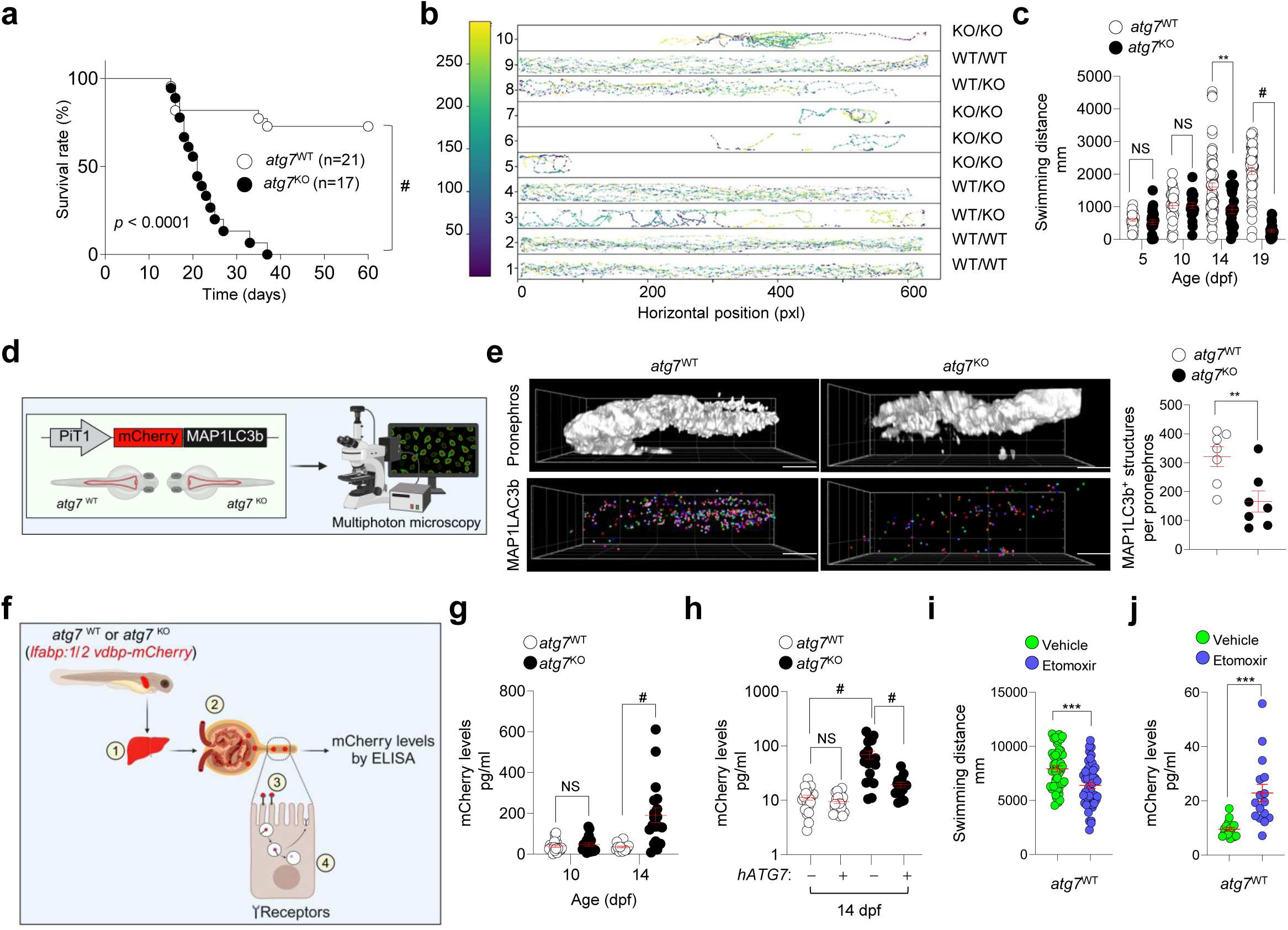
Loss of atg7-dependent autophagy impairs functionality and homeostasis of the zebrafish pronephros. (**a**) Kaplan-Meier survival curves of *atg7*^WT^ (n = 21) and *atg7*^KO^ (n = 20) zebrafish larvae. (**b**) Representative swimming tracks and (**c**) quantification of the total distance quantification swum by zebrafish larvae at 5,10,14, and 19 dpf; n = 29 *atg7*^WT^ and 27 *atg*7^KO^ zebrafish larvae at 5dpf, n = 32 *atg7*^WT^ and 20 *atg*7^KO^ zebrafish larvae at 10dpf, n = 64 *atg7*^WT^ and 21 *atg*7^KO^ zebrafish larvae at 14dpf, n = 52 *atg7*^WT^ and 24 *atg*7^KO^ zebrafish larvae at 19dpf. (**d**) Schematic of the experimental workflow used to assess autophagy in zebrafish larvae. (**e**) Multiphoton imaging and quantification of the number of MAP1LC3B^+^ structures, n = 7 zebrafish larvae per genotype. (**f**) Schematic of the experimental workflow used to measure PT function/tubular proteinuria in zebrafish larvae: ½vdbp-mCherry (∼50 kDa) is produced in the liver, filtered by the glomerulus, reabsorbed by PT cells, and processed via the endolysosomal pathway (EE = early endosome, LE = late endosome, Lys = lysosome). (**g,h,j**) Quantification of urinary mCherry levels by ELISA; in (**g**), n = 19 *atg7*^WT^ and 20 *atg*7^KO^ zebrafish larvae at 10 dpf; and n = 19 zebrafish larvae at 14 dpf; in (**h**), n = 20 *atg7*^WT^, 19 *atg*7^WT^ + *hATG7*, 19 *atg*7^KO^, and 16 *atg*7^KO^ + h*ATG7* zebrafish larvae; in (**j**) n = 16 zebrafish larvae per condition. (**i**) Swimming tracking and distance quantification in treated zebrafish; n= 46 in vehicle- and etomoxir-treated zebrafish larvae. Bar graphs show mean ± SEM. Statistics were calculated using unpaired two-tailed Student’s *t*-test, **P < 0.01, ***P < 0.001 and ^#^P < 0.0001 relative to *atg7*^WT^ or *atg7*^KO^. NS, not significant. Scale bar, 200µm.

### Association of ATG7 variants with kidney function and human diseases

Our findings across multiple model systems highlight the essential role of ATG7 in sustaining PT homeostasis and normal physiology. While autophagy dysfunction has long been implicated in disease pathogenesis,^5^ direct associations between human *ATG7* variants and kidney phenotypes have remained elusive. To address this gap, we used large-scale phenome-wide association meta-analysis (PheWAS) involving 635,969 individuals from the VA Million Veteran Program^34^ and identified a genome-wide significant association (p=2×10^11^) with a rare intronic *ATG7* variant (rs375156407; MAF=0.06%), linked to multiple kidney-related conditions (Fig. 8a; Supplementary Fig. 15a-15b), suggesting a pathogenic or risk-modifying role for autophagy gene variants in human disease.^5,35^ Further, analysis of Human Genetic Evidence (HuGE) scores from the Common Metabolic Diseases Knowledge Portal (https://md.hugeamp.org/) — which integrates genome-wide association studies across diverse populations and phenotypes — revealed genome-wide significant associations between *ATG7* variants and multiple kidney-related traits, including blood urea nitrogen (BUN), estimated glomerular filtration rate (eGFR) by creatinine and cystatin C, and cardiometabolic traits known to influence kidney function, such as blood pressure regulation, hypertension, waist-hip ratio (a surrogate for visceral adiposity), and specific circulating lipid species, including TAGs (Fig. 8b).

**Fig. 8.**
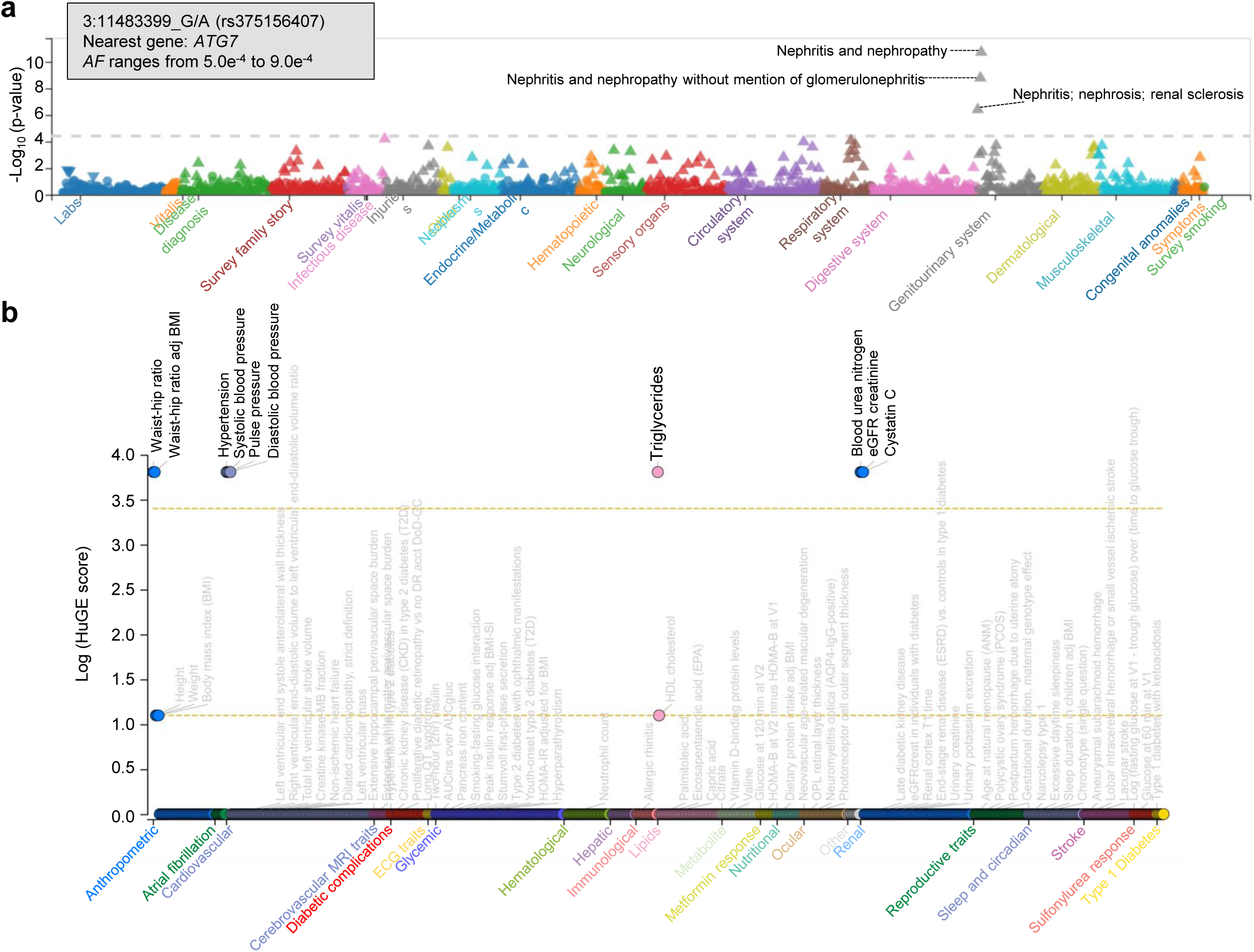
Association of *ATG7* variants with cardio-renal-metabolic traits and increased kidney disease risk. (**a**) Phenome-wide (PheWAS) for variant rs375156407, highlighting phenome-significant associations with nephritis, nephropathy, and related traits in the Million Veteran Program (MVP) biobank. Traits are grouped and color-coded by biological categories. The horizontal grey line denotes the phenome-wide significance threshold. Direction of effect is indicated by triangles: upward-facing for positive associations and downward-facing for negative associations with the alternative allele. (**b**) HuGe score plot showing genome-wide significant associations between *ATG7* and traits catalogued from the Common Metabolic Diseases Knowledge Portal.

At the other end of the genetic spectrum, ultra-rare biallelic ATG7 variants were recently reported in five families presenting with neurodevelopmental disorders.^21^ Two adult female siblings (P1 and P2) from one family carried compound heterozygous loss-of-function variants (c.1975C>T [p.Arg659]; c.2080-2A>G, NM_006395.2), resulting in undetectable ATG7 protein in multiple tissues.^21^ This prompted us to test whether a congenital lack of ATG7 in humans might also cause tubular proteinuria and kidney tubulopathy. While classical markers of tubular proteinuria (e.g., RBP, aminoaciduria) were absent in both siblings, P2 showed elevated urinary CC16 — a sensitive LMW PT dysfunction marker — compared to P1 and healthy controls (Supplementary Fig. 16a). Urine metabolomic profiling revealed distinct metabolic signatures in both siblings relative to controls (Supplementary Fig. 16b-16c). Notably, P2 exhibited enrichment in pathways related to glucose, amino acid, and nucleotide metabolism, fatty acid degradation, the TCA cycle, and cancer metabolism (Supplementary Fig. 16d), paralleling the metabolic abnormalities observed in ATG7-deficient preclinical models.

### ATG7 impacts metabolism, differentiation and prognosis in renal cell carcinoma

Clear cell renal cell carcinoma (ccRCC), the most prevalent form of kidney cancer, originates from proximal tubule (PT) cells and is characterized by lipid droplet (LD) accumulation.^36,37^ Given the enrichment of cancer metabolism signatures and LD accumulation in ATG7-deficient mouse PT cells (mPTCs) and metabolic alterations in human urine samples, we investigated the potential role of ATG7 in ccRCC. In the UK Biobank cohort (n = 394,841), predicted loss-of-function variants in *ATG7* were significantly associated with malignant neoplasms of the kidney (ICD-10 C64; p=2.16×10⁻^4^, β=0.097; Supplementary Fig. 17a).

Consistently, analysis of The Cancer Genome Atlas (TCGA-KIRC) revealed that ccRCC patients with the lowest tertile of *ATG7* expression had significantly worse overall survival compared to those with higher expression (p=2.23e^-02^; Fig. 9a). To further explore this relationship, we conducted a meta-analysis of six transcriptomic datasets from ccRCC tumors^40^ (656 tumors vs.146 controls; Fig. 9b). This analysis revealed coordinated upregulation of proliferation and dedifferentiation markers, and downregulation of FA metabolism and PT-cell-specific genes (all combined q < 0.001, except *SOX9*; Fig. 9c).

**Fig. 9.**
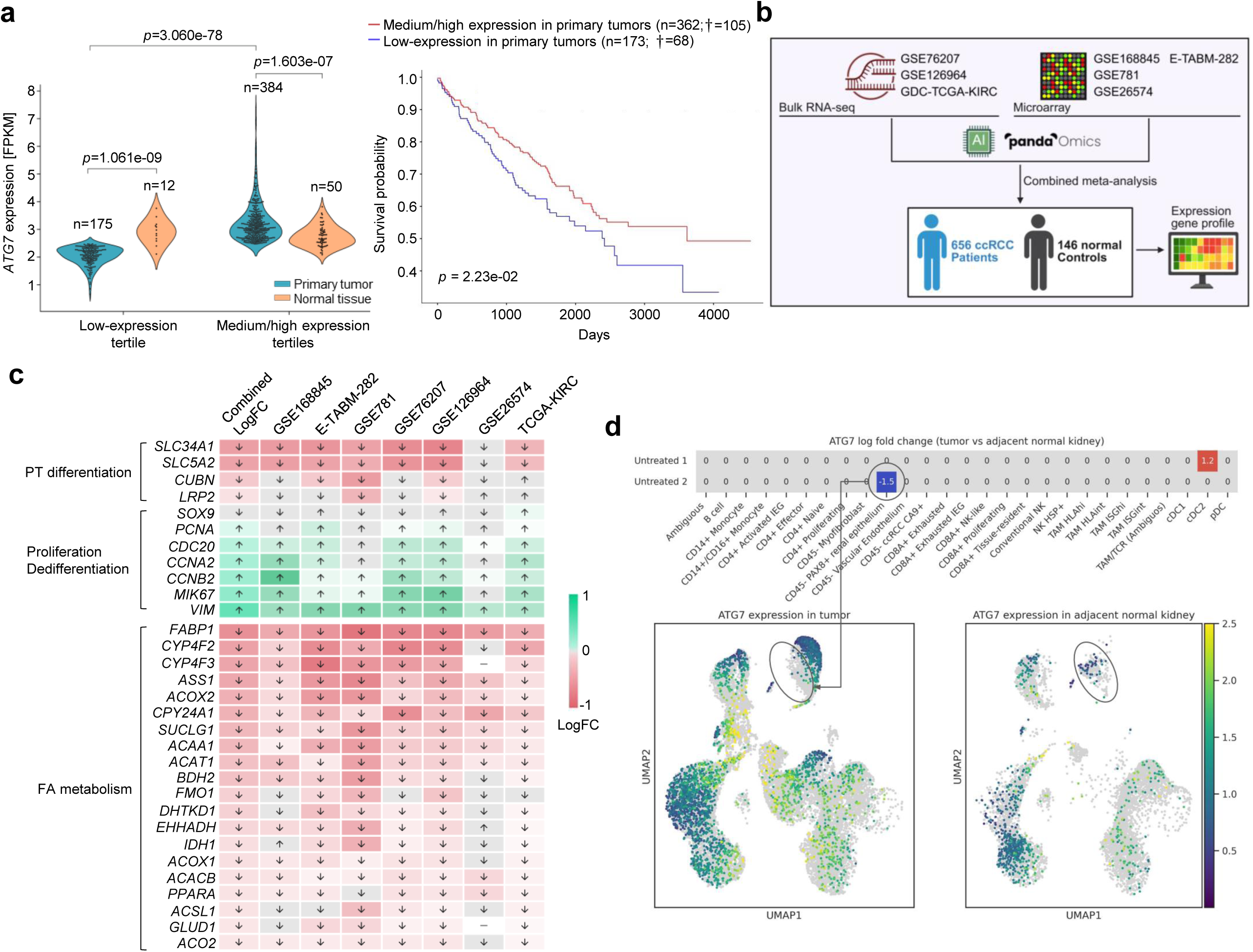
Low *ATG7* expression correlates with transcriptional signatures of metabolic reprogramming, loss of PT identity, and poor prognosis in patients with renal cell carcinoma. (**a,** right) *ATG7* mRNA expression in normal solid tissues versus primary tumor tissues from ccRCC individuals stratified by ATG7 expression tertiles (low vs. medium/high). Statistical comparisons were performed using the Wilcoxon test. Left: Kaplan–Meier survival analysis of patients with kidney renal clear cell carcinoma (KIRC), stratified by tumor ATG7 expression tertiles. Patients with low ATG7 expression (blue) were compared to those with medium/high expression (red). Significance was assessed using the log-rank test. (**b**) Schematic of the transcriptomic analysis workflow. Publicly available datasets from patients with clear cell renal cell carcinoma (ccRCC) were analyzed using PandaOmics, followed by meta-analysis and pathway enrichment analysis. (**c**) Heat maps showing differential mRNA expression of PT-specific markers, fatty acid oxidation (FAO) genes, and genes associated with proliferation and dedifferentiation in ccRCC tumor tissue versus matched normal tissue. Expression patterns and log fold changes were derived from the PandaOmics ‘Expression’ tool. (**d**) Heat map and UMAPs displaying *ATG7* mRNA expression across different cell populations in untreated ccRCC tumour tissues compared to adjacent normal kidney samples. Statistical significance was assessed using the Wilcoxon test (P < 0.05).

Single-cell RNA sequencing (scRNA-seq) from two treatment-naïve ccRCC patients corroborated these findings, showing significantly reduced *ATG7* expression in *PAX8*⁺ tumour cells compared to adjacent normal PT epithelia [log_2_ fold change (FC)= –1.20; p=0.05; Fig. 9d). Notably, the patient with the most pronounced *ATG7* downregulation also exhibited the highest tumour burden (Supplementary 17b-17c). In tumour cells, this loss of *ATG7* was associated with activation of epithelial-to-mesenchymal transition (EMT) pathways, suppression of nutrient transporters, downregulation of mitochondrial metabolism, and loss of differentiation programs (Supplementary 17d-17e). Collectively, these data implicate that ATG7 deficiency may contribute to the lipid-rich, dedifferentiated, and proliferative phenotype characteristic of ccRCC, positioning ATG7 as a prognostic marker and potential candidate therapeutic target in kidney cancer.

## Discussion

Coordination of metabolism and differentiation is fundamental to tissue homeostasis and organ function. Here, we identify ATG7-dependent autophagy as a central regulator of epithelial cell fate in PT of the kidney — a metabolically demanding nephron segment specialized for nutrient reabsorption and transport. We demonstrate that ATG7 is essential for LD metabolism through the maintenance of lipophagy flux, enabling the mobilization of FAs to mitochondria to support oxidative phosphorylation and meet the high energetic demands required for PT-specific homeostatic functions, including transport and reabsorptive activities (Fig. 10).

**Fig. 10.**
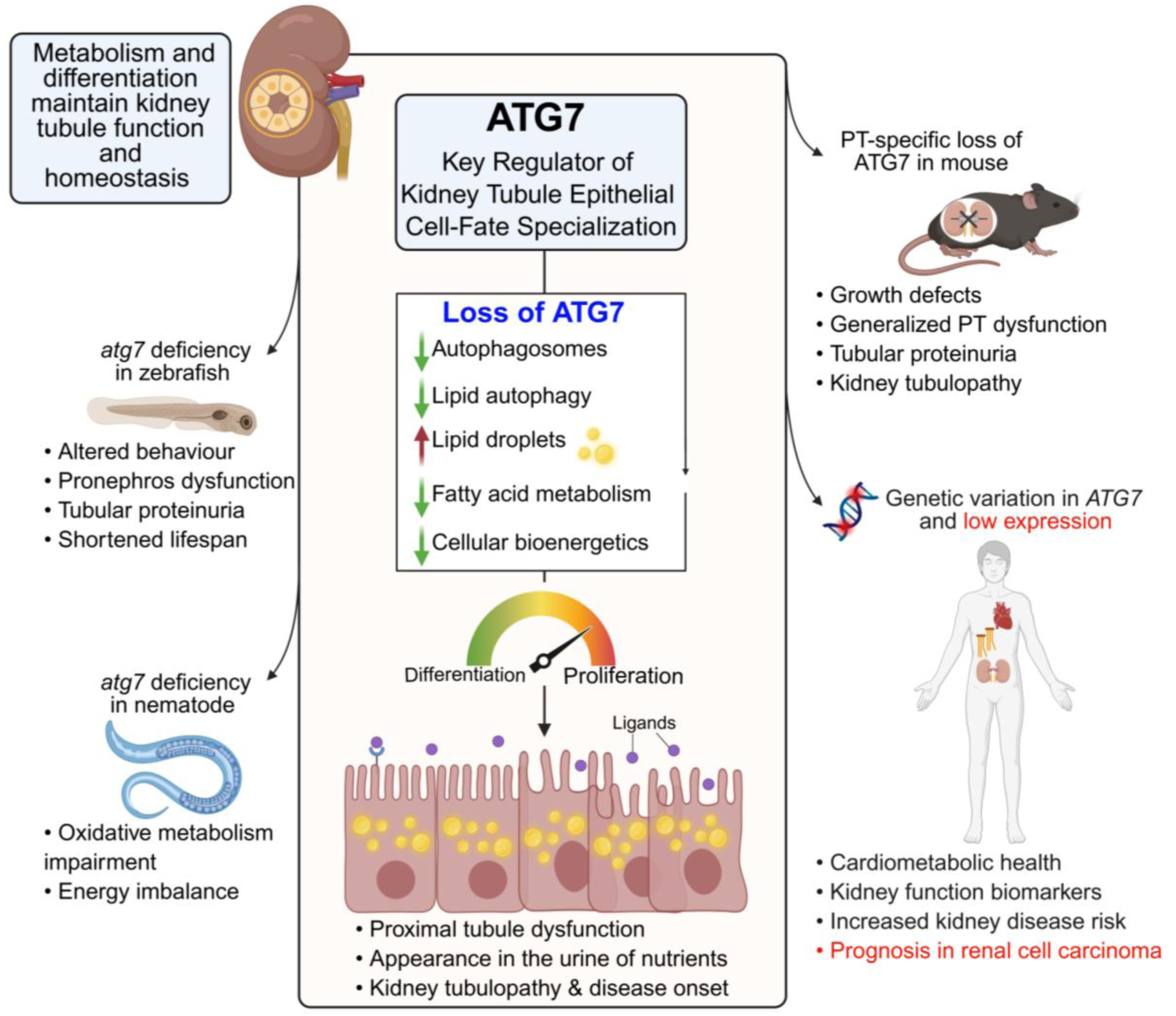
Autophagy regulator ATG7 functions as a conserved metabolic switch governing cell-fate decisions in kidney tubule health and disease. Specialized epithelial cells must couple metabolic programs with differentiation signals to fulfil their physiological roles and maintain homeostasis. In the kidney proximal tubule, ATG7-dependent autophagy acts as a central integrator of these processes, directing metabolic and differentiation trajectories that sustain epithelial specialization and reabsorptive function. Loss of ATG7 disrupts this integration, shifting cells toward an anabolic, proliferative, and dedifferentiated state. Mechanistically, ATG7 deficiency impairs lipophagy, resulting in lipid droplet accumulation and insufficient fatty acid supply to mitochondria, thereby compromising oxidative metabolism, depleting energy production, and ultimately triggering functional decline. These defects are observed in both nematode and murine cells, underscoring the evolutionary conservation of this metabolic axis. Across vertebrate models — from zebrafish to mice — autophagy failure results in phenotypic alterations, including solute transport defects, nutrient-wasting proximal tubulopathy, and systemic metabolic imbalance. Clinically, ATG7 genetic variants are associated with cardio-renal-metabolic traits and increased risk of kidney disease. In patients with clear cell renal cell carcinoma, low ATG7 expression correlates with transcriptional signatures of metabolic reprogramming, epithelial dedifferentiation, and poor prognosis, identifying ATG7 as a potential prognostic biomarker and therapeutic target in kidney disease and cancer.

Mechanistically, ATG7 deficiency — induced via genetic deletion in mice, adenoviral knockdown in primary cells, or CRISPR-based disruption in zebrafish — consistently led to LD accumulation, mitochondrial dysfunction, and a shift toward anabolic, proliferative states that impairs differentiation and receptor-mediated endocytosis. These phenotypes are conserved across species and are functionally linked to the loss of epithelial specialization.

In humans, common genetic variants in ATG7 are associated with kidney function biomarkers, cardiometabolic traits, and increased disease risk in large-scale phenome-wide genetic analysis across multiple populations. At the other end of the genetic spectrum, ultra-rare biallelic loss-of-function mutations human *ATG7* lead to mitochondrial abnormalities and tubular dysfunction — as observed in an affected family member. In clear cell renal cell carcinoma (ccRCC), reduced ATG7 expression correlates with metabolic reprogramming, loss of differentiation markers, and poor clinical outcomes, supporting the clinical relevance of this autophagy-lipid metabolism axis for kidney disease, cancer, and metabolic health (Fig. 10). Collectively, our findings position ATG7 as a gatekeeper of metabolic fidelity and epithelial identity, and a promising therapeutic target for restoring physiological homeostasis in a broad spectrum of currently intractable diseases.

In genetically engineered models, high-resolution imaging and functional analyses revealed that loss of ATG7 in PT cells results in the accumulation of damaged mitochondria and SQSTM1-positive protein aggregates — canonical hallmarks of autophagy failure. This buildup of cellular stress drives metabolic reprogramming and diminishes reabsorptive capacity, culminating in urinary loss of essential nutrients and causing proximal tubulopathy. Thus, beyond maintaining quality control,^41^ ATG7-mediated autophagy maintains the metabolic trajectories of PT cells, preserving their identity and specialized functions.

Strikingly, ATG7-deficient murine PTs recapitulate key features of inherited proximal tubulopathies, including defective endocytosis, aberrant growth signalling, and loss of key nutrient transporters and endocytic receptors.^2,6,7^ While ATG7 is best known for canonical autophagy, it may also engage in non-canonical pathways such as LC3-associated phagocytosis (LAP^29^) and LC3-associated endocytosis (LANDO^30^). However, loss of FIP200 — a core autophagy component required for canonical but not non-canonical pathways — phenocopies the epithelial dysfunction seen in ATG7-deficient cells, emphasizing the critical role of canonical autophagy in maintaining epithelial specialization and homeostatic function.

Supporting the evolutionary conservation of this role, zebrafish larvae lacking *atg7* exhibit hallmark features of autophagy-deficient mice^10,13^ — including impaired motility, low molecular weight proteinuria, and early lethality, all of which are rescued by re-expression of wild-type human ATG7, confirming its conserved function in kidney tubule epithelial differentiation and normal physiology.

How does loss of ATG7 compromise PT cell-fate specialization and physiological homeostasis? Tubular epithelial cells rely on FAO and oxidative phosphorylation to sustain their high metabolic demands required for nutrient transport and solute reabsorption.^17,19,20^ Lipid droplets serve as energy reserves, releasing fatty acids (FAs) via cytosolic lipolysis and autophagy-dependent degradation (lipophagy).^19,26,42^ Using integrated multi-omics landscapes and lipid metabolism functional assays, we show that ATG7 deficiency impairs lipophagy, thereby limiting FA release, impairs FAO and ATP production, and drives mitochondrial dysfunction — evidenced by organelle fragmentation, membrane potential loss, oxidative stress, and impaired respiratory capacity. These defects underscore the requirement of autophagy for mitochondrial fitness and metabolic energy supply in differentiated PT epithelial cells. This energy crisis, in turn, induces a shift toward anabolic and proliferative programs at the expense of epithelial differentiation, ultimately manifesting as nutrient-wasting tubulopathy.

Lipid dysmetabolism and LD accumulation — whether from defective lipophagy or increased de novo lipogenesis — are emerging as unifying features of diverse kidney diseases, both metabolic and non-metabolic.^42^ Spatial metabolomics of diseased human kidneys reveals reduced FAO, lipid buildup, and mitochondrial dysfunction in PT cells, implicating dysregulated kidney lipid metabolism as a central pathogenic mechanism driving disease progression.^42^ Preclinical studies have shown that activation of lipid-catabolic regulators, including PPARα, ERRα, and CPT1A, mitigates kidney injury and fibrosis in chronic kidney disease (CKD) models.^42^ In line with these findings, we show that pharmacologic activation of cytosolic lipolysis via the PPARα agonist fenofibrate restores mitochondrial energy metabolism and function, PT-cell-specific gene expression, receptor-mediated endocytosis, and structural features of differentiation — including primary cilia^43^ — in ATG7-deficient cells, despite persistent autophagy impairment. These results define a conserved ATG7-lipophagy-lipid metabolism axis as a critical licensing mechanism that governs epithelial cell-fate specialization and differentiation in the kidney tubule.

Translational relevance is supported by integrative human genetics. Our genome- and phenome-wide association studies link common *ATG7* variants with kidney function, cardiometabolic traits, and kidney disease risk. Longitudinal lipidomics in >91,000 individuals further implicates disrupted lipid metabolism — marked by elevated VLDL, triglycerides, and cholesteryl esters — as early predictors of chronic kidney disease (CKD).^44^ Additionally, deep clinical phenotyping of a family with biallelic pathogenic *ATG7* variants unveils mitochondrial dysfunction and subtle PT defects, including proteinuria in an affected family member — subtle phenotypes that may be missed in standard clinical assessments — suggesting the existence of alternative degradative pathways or yet-to-be-discovered compensatory mechanisms in humans.^45^

Notably, *ATG7* downregulation also marks poor prognosis in clear cell renal cell carcinoma (ccRCC) — the most prevalent kidney cancer subtype arising from proximal tubule (PT) cells and characterized by lipid droplet accumulation.^36,37^ Transcriptomic analyses show that low *ATG7* expression associates with enhanced transcriptional signatures related to epithelial-to-mesenchymal transition and suppression of FA metabolism, metabolic reprogramming, and differentiation — paralleling our findings in ATG7-deficient models and inherited proximal tubulopathies such as cystinosis.^6,7^ These shared signatures support a unifying model in which ATG7 couples lipid turnover and homeostasis with mitochondrial bioenergetics to maintain PT-cell identity and epithelial specialization. Disruption of this genetic and metabolic axis contributes not only to proximal tubulopathy and disease development but may also be co-opted by malignant kidney cells during tumorigenesis. Lipid metabolic dysfunction and defective lipophagy — defining features of ATG7 loss — are emerging as shared drivers of CKD and cancer cell adaptation.^46^

In summary, our findings define an evolutionarily conserved genetic paradigm linking autophagy to energy metabolism in directing cell-fate decisions and epithelial specialization during kidney tubule differentiation. This genetic–metabolic axis deepens our understanding of epithelial tube development and function, with broad implications for health, as well as the pathogenesis of kidney disorders and cancer. Moreover, it provides a mechanistic framework with translational relevance, offering new avenues for therapeutic discovery and metabolic intervention in currently intractable diseases.

## MATERIAL AND METHODS

### Ethical compliance

All the animal experiments were approved by the Cantonal Veterinary Office Zurich (license no. ZH195/2020 and ZH139/2022) and performed following the guidelines of the University of Zurich.

### Human studies

Patients with an established diagnosis of ATG7-related spinocerebellar ataxia (MIM #619422), i.e. with biallelic pathogenic *ATG7* variants were recruited (Family 1; ref. 21). Written informed consent was obtained from the parents of the siblings in the study and controls following the Declaration of Helsinki protocols, and the experimental protocol was approved by the local institutional review board. Urine samples were collected in the morning and kept at 4°C before it was frozen at -80 °C. For immunoblot analysis, the loading volume was indexed to creatinine to correct for variations in urine concentration. Urinary creatinine levels were measured using UniCel DxC 800 proSynchron (Beckman Coulter, Fullerton, CA, USA), according to manufacturer instructions.

### Antibodies and reagents

The following antibodies were used in this study: rabbit anti-Atg7 (Sigma-Aldrich, A2856; 1:200); rabbit anti-Apg5l/Atg5 (Abcam, 108327, 1:500); mouse anti-AQP1(Abcam, ab9566, 1:400); mouse anti-AMPKα (Cell Signaling Technology, 2793; 1:500); rabbit anti-phospho-AMPKα (Thr172) (Cell Signaling Technology, 2535; 1:500); mouse anti-β-actin (Sigma-Aldrich, A5441, 1:1000); rabbit anti-cleaved caspase 3 (Cell Signaling Technology, 9662, 1:400); rabbit anti-caspase 3 (Cell Signaling Technology, 9661, 1:400); rabbit anti-phospho-4E-BP1 (Ser65; Cell Signaling Technology, 9451, 1:500); rabbit anti-4E-BP1 (Cell Signaling Technology, 9644, 1:500); rabbit anti-FIP200 (Cell Signaling Technology, 12436S, 1:200); sheep anti-Lrp2/megalin (1:1000) was kindly provided by P. Verroust and R. Kozyraki (INSERM, Paris, France); rabbit anti-Laminin gamma 1 (clone A5; Thermo Fisher Scientific, MA5-14649, 1:400); rat anti-LAMP1 (Santa Cruz Biotechnology, sc-19992, 1:1000); rabbit anti-LC3 (PM036, MBL, 1:200); rabbit anti-NCoR1 (Cell Signaling Technology, S9448S; 1:500); anti-PPARα (Abcam 126289: 1:500); mouse anti-PCNA (Dako, M0879, 1:500); guinea pig anti-Plin2 (Progen, GP40; 1:100); anti-SDH (Abcam, ab14715, 1:500); anti-PDHA1 (Abcam, ab110334, 1:500); anti-VDAC1 (Cell Signaling Technology, 4866, 1:500); anti-COX IV (Abcam, ab14744, 1:500); anti-cytochrome *C* (Abcam, ab110325, 1:500); rabbit anti-Rab5 (Abcam, ab218624; 1:500); rabbit anti-Rab7 (Abcam, ab137029; 1:500); rabbit anti-S6 ribosomal protein (Cell Signaling Technology, 2217; 1:500); rabbit anti-phospho-S6 ribosomal protein (Ser235/236) (Cell Signaling Technology, 4858; 1:500); mouse anti-SQTM1 (Abcam, ab56416; 1:500); rabbit anti-SQTM1 (Abcam, ab109012; 1:400); mouse anti-transferrin receptor (H68.4; Thermo Fisher Scientific 13-6800; 1:400); anti-acetylated tubulin (Sigma Aldrich T7451; 1:400); rabbit anti-VDBP (Dako A/S , Glostrup Danmark A0021, 1:500); human anti-Transferrin (Dako, A0061; 1:500); mouse anti-TOM20 (Abcam; ab186735; 1:400); rabbit anti-uteroglobin (also known as CC16, Abcam, ab40873; 1:500); sheep anti-uromodulin (K90071C, Meridian Life Science Inc., Cincinnati, OH, USA; 1:300); donkey-anti rabbit IgG, cross-absorbed secondary antibody, Alexa Fluor 488 (Thermo Fischer Scientific, A10040, 1:400); goat-anti mouse IgG, cross-absorbed secondary antibody, Alexa Fluor 633 (Thermo Fischer Scientific, A21052, 1:400); goat-anti mouse IgG, cross-absorbed secondary antibody, Alexa Fluor 546 (Thermo Fischer Scientific, A11003, 1:400); goat-anti rat IgG, cross-absorbed secondary antibody, Alexa Fluor 633 (Thermo Fischer Scientific, A21094, 1:400); donkey-anti goat IgG, cross-absorbed secondary antibody, Alexa Fluor 633 (Thermo Fischer Scientific, A21094, 1:400); donkey-anti sheep IgG, cross-absorbed secondary antibody, Alexa Fluor 633 (Thermo Fischer Scientific, A21100, 1:400). Compounds included Bafilomycin A1 (BafA1; Enzo Life Sciences, ALX-380-030, 250 nM), the biotinylated Lotus Tetragonolobus Lectin (LTL; Vector Laboratories, B-1325), and 4′,6-diamino-2-phenylindole dihydrochloride (DAPI; D1306, Thermo Fischer Scientific; 1:1000), Etomoxir (Merck; 23620), and Fenofibrate (Sigma Aldrich; F620), PIK3C3/Vps34 inhibitor SAR-405 (A8883, APExBIO). All molecules were of at least 98% purity and dissolved in dimethylsulfoxide (DMSO) for experiments unless specified. BSA-conjugated oleic acid (OA; Cayman Chemical, 29557; 300 µM) was diluted in the culture medium. The cells used in this study were negatively tested for mycoplasma contamination using MycoAlert™ Mycoplasma Detection Kit (LT07-118, Lonza, Switzerland).

### *C. elegans* strain maintenance

For all experiments using invertebrate *C. elegans*, hermaphroditic animals were used (wild-type and *atg7* mutant). Worms from mutant deletion strains were genotyped by PCR, and the PCR amplicon sizes were used to check for the presence of the deletion. The ages of adult animals are specifically defined in the figures and figure legends. The nematodes were raised at 20 °C on nematode growth medium (NGM) agar (51.3 mM NaCl, 0.25% peptone, 1.7% agar, 1 mM CaCl_2_, 1 mM MgSO_4_, 25 mM KPO_4_ and 12.9 µM cholesterol, pH 6.0). Fed worms were maintained on NGM agar plates that had been seeded with *Escherichia coli* OP50 bacteria. Synchronous populations of worms were obtained by bleaching young adult hermaphrodites. Briefly, adult hermaphrodites were exposed to 1 mL bleaching solution (0.5 M NaOH and 20% bleach) for 5 min to isolate eggs, and eggs were then washed three times in M9 buffer (22 mM KH_2_PO_4_, 42 mM Na_2_HPO_4_, 85.5 mM NaCl and 1 M MgSO_4_) before plating.

### Quantification of lipid droplets and autophagy in *C. elegans*

To visualize LDs, we utilised a reporter strain expressing DHS-3 fused to GFP [LIU1: (N2; IdrIs1 (dhs-3p::dhs-3::GFP; unc-76+)], LRL262: [*atg-*7*(bp411)*; IdrIs1 (dhs-3p::dhs-3::GFP; unc-76+)], a well-established marker for LDs in *C. elegans*.^31^ DHS-3 is orthologous to the mammalian 17β-HSD11 and is among the most abundant LD-associated proteins identified via mass spectrometry in *C. elegans*.^47^ Its localisation to the LD membrane was confirmed through fluorescence microscopy. To visualise autophagy, we employed a reporter strain expressing SQST-1 fused to RFP [(LRL160: [N2; llcIs12 (p62p::p62::RFP::unc-54 3’UTR; myo-2::GFP)] and LRL256: [*atg-7(bp411);* llcIs12 (p62p::p62::RFP::unc-54 3’UTR; myo-2::GFP)], which is orthologous to the mammalian autophagy receptor SQSTM1/p62. Imaging was performed on day 3 or 4 of adulthood. Approximately 10 worms were imaged per condition for each experiment. Worms were mounted on a 2% agar pad, anaesthetised with 50 mM sodium azide, and covered with a glass coverslip for imaging. Confocal images were captured using an Olympus Fluoview FV3000 microscope equipped with the FV316-SW software (version 2.3.2.169). A 60% oil objective (UPLAN 60x oil, 2.35NA Olympus, Japan) and a 1.90-μm pinhole (resulting in an optical section thickness of 2.2μm) were used. SQST-1::RFP reporter strain imaging was performed using a Zeiss V20 Discovery microscope equipped with an AxioCam 820 mono camera, Achromat S 1.0x FWD 63mm objective, and the Zeiss Zen 3.8 software. Imaging conditions, such as laser power and exposure time, were kept consistent across all samples. The mean fluorescence intensity was quantified using Fiji (version 2.0.0) by applying uniform thresholds to all images for consistency.

### Oxygen consumption measurements in *C. elegans*

Worms were synchronized using a standard hypochlorite treatment protocol. Eggs were placed on OP50-seeded NGM plates and grown to day 2 of adulthood. Adult worms were hand-picked and transferred to P10 plates to control population density, with two plates containing 500 worms each and two plates containing 250 worms each. Worms were allowed to acclimate at 20°C for 30 minutes to minimize stress before measurements. Worms were harvested by rinsing the plates twice with 7 mL of M9 buffer, ensuring thorough collection. The suspensions were centrifuged at 1,000 rpm for 90 seconds, and the supernatant was discarded. Worms were washed three times with 13 mL of M9 buffer to eliminate residual bacteria and debris. After the final wash, 100μL of the worm suspension was transferred into a pre-calibrated Oroboros chamber using a Pasteur pipette, ensuring minimal loss and preventing worms from adhering to surfaces. Oxygen consumption was continuously monitored using the Oroboros O2k system. Measurements were recorded until the nematodes exhibited signs of mortality. Data were normalized to the initial number of worms introduced into the chamber. All analyses were conducted using DatLab software.

### Generation and maintenance of *atg7* zebrafish

The pT7-gRNA and pT3TS-nCas9n plasmids were sourced from Addgene (#46759 and #46757, respectively). To monitor CRISPR-Cas9-mediated mutations and facilitate genotyping, we selected the CRISPR-Cas9-targeted region (5′-GGCGAACTGAAGCTTCAGACTGG-3′), which contains the HindIII restriction site (AAGCTT). Oligonucleotides containing the sgRNA-targeting sequence were annealed and cloned into the Esp3I (BsmBI)-digested *pT7-gRNA* vector, resulting in *pT7-gRNA-atg7*. The oligonucleotide sequences were CRISPR-atg7-S: 5′-TTAGGCGAACTGAAGCTTCAGAC-3′ and CRISPR-atg7-AS: 5′-AAACGTCTGAAGCTTCAGTTCG-3′. Cas9 messenger RNA was produced by utilizing the XbaI-linearized *pT3TS-nCas9n* vector with the mMESSAGE T3 kit (Invitrogen). Both sgRNA-*atg7* and Cas9 mRNA were co-injected into one cell stage AB zebrafish embryos (*Danio rerio*). Genomic DNA was extracted from 2 dpf embryos that developed normally to detect CRISPR-Cas9-induced mutation. Mosaic AB embryos injected with CRISPR-Cas9 were raised to adulthood (F0) and outcrossed with wild-type AB zebrafish. The resulting embryos (F1) were raised to adulthood, and heterozygous carriers were screened. We identified a heterozygous carrier with the mutation of 6bp-deletion + 2bp insertion (*atg7*^+/del4^), and subsequent generations (F1) were crossed to obtain homozygous mutants carrying *atg7*^del4/del4^. The CAS-OFFinder analysis confirmed that there were no off-target edits in other genes. Zebrafish larvae (AB background) were maintained on a 14/10 h day/night cycle at 28°C and fed a standard diet (Zebrafeed 100-200, SPAROS, Portugal) from 5 dpf. Experiments were primarily conducted on larvae aged 5-14 dpf, with animal sex not specified at this developmental stage. Stable zebrafish lines expressing mCherry-tagged half vitamin D binding protein (½vdbp-mCherry) in the liver or hMAPLC3B in the kidney were established. These lines were subsequently outcrossed with *atg7*^+/del4^ zebrafish to generate a transgenic mutant line. The latter was further crossed with *atg7*^+/del4^ zebrafish to produce homozygous larvae. The measurement of ½vdbp-mCherry and mCherry-hMAPLC3B-flagged autophagic vesicles was assessed by ELISA and multiphoton microscopy, respectively.

### Complementation studies in zebrafish

For the generation of transgenic lines with a proximal tubule-specific expression of human *ATG7*, multisite Gateway cloning technology was used to generate the expression vector *PiT1::EGFP-hATG7* using plasmids coming from Tol2 kit v1.2. The cloning of the promoter of the zebrafish phosphate transporter 1/*slc20a1a* (*p5E-PiT1*) genes was previously described.^6,7^ Human *ATG7* cDNA was amplified from the vector *pCMV-myc-ATG7* (kindly provided by Toren Finkel (Addgene plasmid # 24921)) and subcloned into the *pENTR/D-TOPO vector* (Invitrogen) after N-Terminal Tagging with EGFP. The final expression vector *PiT1::EGFP-hATG7* was realized after LR Reaction (Gateway LR Clonase II Enzyme Mix, Invitrogen), which utilizes the LR Clonase for vector construction through plasmids *p5E-PiT1*, *pED-EGFP-hATG7*, *p3E-polyA* and destination vector *pDestTol2CG2* with *cmlc2::EGFP* transgenesis marker. Plasmid DNA of *PiT1::EGFP-hATG7* vector was co-injected with Tol2 transposase mRNA into zebrafish embryo at the one-cell stage to generate a stable transgenic line. Zebrafish *atg7*^+/del4^, *Tg(PiT1::EGFP-hATG7)* was generated after crossing the *Tg(PiT1::EGFP-hATG7)* with *atg7* mutant zebrafish. For the rescue of LMW proteinuria, *atg7*^+/del4^, *Tg(PiT1::EGFP-hATG7)* was crossed with *atg7*^+/del4^, *Tg(lfabp::½vdbp-mCherry)* to produce double transgenic *atg7*^WT^ and *atg7*^KO^larvae. Overnight urine collection was performed at 10 and 14dpf for ELISA assay following the protocol described below.

### Swimming behaviour

The movement tracking of zebrafish larvae was conducted using a custom-built setup, comprising a projector (HP vp6111) for background light, an infrared light source for camera-tracking, and a high-speed camera (Allied Vision Stingray) equipped with a Fujinon 1:1.4/12.5mm objective (Fujinon). Both tracking and background light control were executed through the open-source tracking software Stytra. In each experiment, up to 10 larvae were situated in a specially designed arena featuring 10 lanes (1cm width x 16cm length), all filled with E3 medium. The tracking process involved monitoring larval swimming distances along each lane over 5 minutes. Larval identity was established by tracing a Region of Interest (ROI) on each lane, and the distance covered was calculated by summing the distances between each consecutive coordinate.

### Zebrafish urine collection and mCherry measurements

Zebrafish larvae (AB background) were placed in a 48-well microplate with one larva/500 mL facility water/well and kept at 28°C for 16 hours, followed by urine collection for ELISA assay. The levels of ½vdbp-mCherry were measured according to the manufacturer’s protocol. Briefly, 50 μl of fish pool water containing urine was distributed to a 96-well microplate pre-coated with anti-mCherry antibody. A 50 μL mixture of capture antibody and detector antibody was added to each well and incubated at room temperature for 1 hour. The wells were rinsed three times with washing buffer and incubated with 100 μL of TMB development solution at room temperature for 10 minutes. The reaction was stopped by adding 100μl of stop solution, followed by reading the absorbance of each well at 450 nm.

### Mouse models

The mouse lines were used: *Fip200*^fl/fl^, *Atg5*^fl/fl^, *Atg7*^fl/fl^ and γ*Gt1*-Cre. The *Fip200*^fl/fl^ and *Atg5*^fl/fl^ mouse lines were kindly provided by Dr. C. Münz (University of Zurich). The *Atg7*^fl/fl^ mice^10^ (C57BL/6J background) were obtained from the RIKEN repository (National BioResource Project, started by the Ministry for Education, Culture Sports, Science and Technology, Japan). The *Atg7*^fl/fl^ mice were generated by inserting two Lox P sites around exon 14, which encodes the active site cysteine essential for activation of the conjugation substrates. The Cre-expressing mouse line was obtained from The Jackson Laboratory (RRID: IMSR_JAX:012841). The rat proximal promoter to γ*Gt1* was used in a transgene construct to drive Cre recombinase expression.^22^ The γ*Gt1-Cre* promoter becomes active around postpartum day 14, coinciding with the completion of nephrogenesis and thus targeting cortical tubular epithelium in mature kidneys.^22^ The Cre-expressing mice (C57BL/6 x SJL background) were backcrossed (>10 times) into the C57BL/6 background. The PT-specific *Atg7*^KO^ mice were generated by crossing mice carrying the respective LoxP-flanked alleles with γ*Gt1*-Cre transgenic mice. All mice were housed at the animal service centre of the University of Zurich under specific pathogen-free conditions and maintained under controlled temperature (22-25°C) and humidity (50-60%), with 12-h dark/light cycle and ad libitum access to tap water and standard chow (Diet AO3, SAFE). Kidneys were collected for analysis at the time of sacrifice. Both female and male mice at 4, 12, and 24 weeks of age were used for kidney function studies, whereas the other experiments were performed using mice at 12 and 24 weeks of age. Littermates not carrying the γ*Gt1-Cre* transgenes were used as controls in all experiments.

### Kidney function and metabolic parameters

The mice were placed overnight in metabolic cages with ad libitum access to food and drinking water; urine was collected on ice, and body weight, water intake and diuresis were measured. Blood (from a sublingual vein) was obtained after anaesthesia with ketamine/xylazine or isoflurane. Urine and blood parameters were estimated using UniCel DxC 800 proSynchron (Beckman Coulter, Fullerton, CA, USA). Urinary levels of low-molecular-weight CC16 were measured by using an ELISA according to the manufacturer’s instructions (BIOMATIK EKU03200, Thermo Fischer Scientific, Waltham, MA). Albuminuria was measured via Coomassie Blue staining by using ProtoBlue Safe (National Diagnostics, EC-722, Atlanta, GA, USA) according to the manufacturer’s instructions.

### Amino acid measurements in the urine

Urinary amino acid concentrations were measured at the Functional Genomic Center Zurich, University of Zurich, using liquid chromatography-mass spectrometry (LC-MS/MS). Before injection, 50μL of urine was mixed with 10% sulfosalicylic acid, precipitated, and centrifuged for 10 minutes at 12000 rpm. The supernatant was then mixed with 30μL borate buffer, 5μL 500μM MSK-CAA-1 internal standard, and 10μL AQC solution, totalling 50μL. After a 10-minute incubation at 55°C, samples were loaded onto a Waters UPLC H-Class Plus system with an Acquity PLC Quadrupole Solvent Manager and Sample Manager. Derivatized amino acids were detected using a Waters Qda single quadrupole mass detector in positive mode with selected ion monitoring (SIR). The column was maintained at 55°C, and 1μL was injected. Gradient elution used 0.1% formic acid in water (eluent A) and in acetonitrile (eluent B) with a constant flow rate of 0.5 mL/min and the following gradient for solvent B: Initial: 1%, 0.0-1 min: 1%, 1-4 min: 13%, 4-8.5 min: 15%, 8.5-9.5 min: 95%, 9.5-11.5 min: 95%, 12-15 min: 1%. Data were analyzed using Masslynx 4.2 (Waters).

### Isolation of PT segments from mouse kidneys and primary cell cultures

Kidneys were harvested from Atg7^PT-KO^ mice and their control littermates or from *Atg5*^floxed/floxed^, *Atg7*^floxed/floxed^, and *Fip200*^floxed/floxed^ mice. One kidney was split transversally; one half was fixed and processed for immunostaining, and the other half was flash-frozen, homogenized in 1mL of RIPA buffer with protease and phosphatase inhibitors, and processed for western blot analysis. The contralateral kidney was used to generate primary cultures of mPTCs. Freshly microdissected PT segments were seeded onto chamber slides and/or 6- or 24-well plates and cultured at 37°C and 5% CO2 in DMEM/F12 with 0.5% dialyzed FBS, 15mM HEPES, 0.55mM sodium pyruvate, 0.1ml L^-1^ non-essential amino acids, hydrocortisone, human EGF, epinephrine, insulin, triiodothyronine, TF, and gentamicin/amphotericin, pH 7.40, 325mOsm kg^-1^. The medium was replaced every 48 hours. Confluent monolayers of mPTCs, expanded from the tubular fragments after 6-7 days, were characterized by a high endocytic uptake capacity and tested negative for mycoplasma contamination.

### Nutrient starvation protocols and cellular treatments

The withdrawal of serum, glucose, and amino acids was achieved by washing mPTCs with Hank’s balanced salt solution and placing them in either a normal growth or nutrient-deprived medium for the indicated times. Lysosomal proteolysis was inhibited, where indicated, by adding BafA1 (250nM in cell culture medium for 4 hours). The cells were then processed and analysed as described below.

### Adenovirus transduction

For expression studies, adenoviral constructs included a CMV control vector (Ad-CMV-GFP; #1060) and a vector carrying Cre-recombinase (Ad-Cre-GFP; #1700), both purchased from Vector Biolabs (University City Science Center, Philadelphia, USA). Adenovirus transduction was performed 24 hours after plating when the cells reached 70–80% confluence. Cells were incubated for 6 hours at 37°C with a culture medium containing the virus at a concentration of 20 x 10^5^ PFU/ml. Fresh culture medium was exchanged every 2 days, and the cells were cultured for 3 days before being collected for analysis.

## Proteomics

### Sample preparation

Cell pellets (∼5×10^5^ cells; n = 4 mice/group) were lysed in 100 μL buffer, sonicated with HIFU (1 min, 85% amplitude), and heated at 95°C for 10 min. Proteins were further extracted using a Tissue Lyser II (2 × 2 min at 30 Hz), heated again for 10 min, and subjected to an additional 1-min HIFU. After centrifugation (20,000 × g, 10 min), protein concentration was measured with a Lunatic instrument. Fifty micrograms of protein per sample was digested by adding 10 μL “Digest” solution and incubating at 37°C for 60 min; digestion was stopped with 100 μL Stop solution. The mixture was filtered (3,800 × g), and peptides were washed, eluted, dried, and reconstituted in 40μL of 3% acetonitrile/0.1% FA with 1μL iRT peptides (1:100 dilution).

### Liquid chromatography-mass spectrometry analysis

Peptides were analyzed on an Orbitrap Fusion Lumos (Thermo Scientific) with a Digital PicoView source (New Objective) and an M-Class UPLC (Waters). Solvent A was 0.1% FA; Solvent B was 0.1% FA in 99.9% acetonitrile. One microliter of peptides was loaded onto an MZ Symmetry C18 Trap Column (100LÅ, 5 μm, 180 μm × 20 mm) and separated on a nanoEase MZ C18 HSS T3 Column (100 Å, 1.8 μm, 75 μm × 250 mm). The elution gradient was: 5% B for 3 min, 5–22% B over 80 min, 22–32% B over 10 min, then washed at 95% B for 10 min and re-equilibrated for 10 min. Samples were run in randomized order. The instrument acquired full-scan MS spectra (m/z 300–1,500) at 120,000 resolution (target 500,000 ions) and data-dependent MS/MS in the ion trap (quadrupole isolation at 0.8 Da, HCD at 35% energy, target 10,000 ions, max 50ms injection time). Only precursors >5,000 intensity were selected (cycle time 3 s); charge states 1, unassigned, and >7 were excluded, with dynamic exclusion set to 20 s (10 ppm). Internal lock masses were m/z 371.1012 and 445.1200.

### Protein identification and label-free protein quantification

Raw data were processed with MaxQuant (v1.6.2.3) using Andromeda against the SwissProt canonical mouse proteome (2019-07-09), its reversed decoy, and common contaminants. Fixed modification was carbamidomethylation (C); variable modifications were methionine oxidation and N-terminal acetylation. Trypsin/P was used, allowing peptides ≥7 amino acids with up to 2 missed cleavages. FDR was set to 1% for peptides and 5% for proteins. Label-free quantification was enabled with a 2-minute match-between-runs window, and each file was processed separately for individual quantification. Protein fold changes were calculated from Intensity values in the proteinGroups.txt file, filtering for proteins with ≥2 peptides and ≤3 missing values. Data were normalized via a modified robust z-score, and p-values were computed using a pooled variance t-test. For proteins missing all measurements in one condition, a pseudo–fold change was calculated using the mean of the lowest 10% intensities. Finally, pathway analysis was performed with Network Analyst (http://www.networkanalyst.ca) on significantly changed proteins in mPTCs from *Atg7* mouse kidneys.

## Metabolomics

### Urine metabolite extraction and LC-MS analysis

#### Extraction

Twenty microliters of creatinine-normalized urine were mixed with 80 μL of pre-cooled methanol. The mixture was incubated at –20°C for 1 h, then centrifuged at 10,000 rpm for 10 min at 4°C. The supernatant was transferred to a clean test tube, dried under a nitrogen stream, and reconstituted in an injection buffer (90% acetonitrile). Following a second vortex and centrifugation (10,000 rpm, 4°C, 10 min), the clear supernatant was transferred to Total Recovery Vials (Waters) for LC-MS injection. Method blanks, standard mixes, and pooled samples were prepared identically to serve as quality controls.

#### LC-MS Data Acquisition

Urine metabolites were separated using a Thermo Vanquish Horizon Binary Pump equipped with a Waters Premier BEH Amide column (150 mm × 2.1 mm). A gradient was applied using solvent A (10 mM ammonium bicarbonate in 5% acetonitrile, pH 9.0) and solvent B (10 mM ammonium bicarbonate in 95% acetonitrile), transitioning from 99% B to 30% B over 12 min. The injection volume was 3.5 μL, with a flow rate of 0.4μL/min; the column was maintained at 40°C and the autosampler at 5°C. The LC was coupled via a HESI source to a Thermo Q Exactive mass spectrometer. MS1 and MS2 data were acquired in negative ion mode using Full MS/dd-MS² (Top5) over a mass range of 70–1050Lm/z, at a resolution of 70,000 for MS1 and >17,500 for MS2. Quality controls were performed using pooled samples, reference compound mixtures, and blanks to assess technical accuracy and stability.

#### Untargeted Metabolomics Data Analysis

Data were processed with Compound Discoverer (Thermo Scientific) in an untargeted workflow that included spectral selection, retention time alignment, compound detection and grouping, gap filling, background filtering, and constant median normalization. MS2-based identities were assigned by scoring fragmentation patterns against the mzCloud and mzVault databases. A filtering process then generated a manually annotated compound table, with features assigned the highest level of confidence based on the following criteria: signal-to-noise ratio > 3, mzCloud or mzVault match score > 50, mass error within ±5 ppm, match to an in-house MS1_RT library within ±10 sec, and acceptable chromatographic peak and MS2 spectral quality.

### Cellular metabolite extraction and LC-MS analysis

#### Sample Preparation

Cellular metabolism was quenched by adding 80% pre-cooled methanol to the cells. After vortexing, samples were incubated at –20°C for 1.5 h. Proteins were precipitated by centrifuging at 10,000 rpm for 20 min at 4°C, and the clear supernatant was transferred to a new vial. The supernatant was then dried under a nitrogen stream and reconstituted in 20 μL water mixed with 80 μL injection buffer (90% acetonitrile, 8.8% methanol, 50 mM ammonium acetate, pH 7). Following another round of vortexing and centrifugation (10,000 rpm, 4°C, 10 min), the clear supernatant was transferred to Total Recovery Vials (Waters) for LC-MS injection. Method blanks, standard mixes, and pooled samples were prepared identically for quality control.

#### LC-MS Data Acquisition

Cell metabolites were separated using a Waters nanoAcquity UPLC with a BEH Amide capillary column (150 μm × 50 μm × 1.7 μm). A gradient was applied using solvent A (5 mM ammonium acetate in water) and solvent B (5 mM ammonium acetate in acetonitrile), transitioning from 5% A to 50% A over 12 min. The injection volume was 1μL, with the flow rate adjusted from 3 to 2 μL/min. The UPLC was coupled to a Waters Synapt G2Si mass spectrometer via a nanoESI source. MS1 and MS2 data were acquired in negative ion mode using MSE over a mass range of 50 to 1200Lm/z at resolutions greater than 20,000.

#### Untargeted Metabolomics Data Analysis

Data were processed using Progenesis QI (Nonlinear Dynamics, Waters). Ion intensity maps were aligned to a reference dataset, and peak picking was performed on an aggregated map. Ions were identified by comparing accurate mass, adduct patterns, and isotope distributions with the Human Metabolome Database (HMDB) using a mass accuracy tolerance of 5LmDa; fragmentation patterns were also considered. All biological samples were analyzed in triplicate, with quality controls run on pooled samples and reference compound mixtures. Finally, pathway analysis of significantly altered metabolites in cells and urine samples was performed using MetaboAnalyst (http://www.metaboanalyst.ca) with default settings.

### Lipidomics

#### Extraction

Lipid extraction was performed following a previously described method^48^, with some modifications. A mixture of 1000 µL isopropanol: methanol (1:1) was added to each cell culture well. The plates were then incubated at -80 °C for 10 minutes. After incubation, the cells were scraped, and the resulting solution was transferred to Eppendorf tubes.

The samples were continuously mixed in a thermomixer (Eppendorf) at 25 °C (950 rpm) for 30 minutes. Protein precipitation was achieved by centrifugation at 16,000 g for 10 minutes at 25 °C. The resulting single-phase supernatant was collected, dried under nitrogen (N₂), and stored at -20 °C until further analysis. Before analysis, the dried lipids were reconstituted in 100 µL methanol: isopropanol (1:1).

#### LC-MS analysis

The lipidomics LC-MS method was performed as previously described, with some modification.^48^ A Waters Premier BEH C18 column (50μm × 2.1μm × 1.7µm) was used at 60°C for LC separation on a Thermo Vanquish Horizon tandem LC system. The injection volume was 2µL in positive mode and 5µL in negative mode, with a flow rate of 1 mL/min. A segmented gradient was applied, transitioning from 15% Buffer B to 99% Buffer B over a total gradient length of 7.5 minutes. The gradient segments were as follows: 0.0-1.0 min; 15-30%B,1.0-1.25 min; 30-48%B,1.25-5.0 min;48-82%B, 5.5-5.75; 82-99%B,5.75-6.0 min; isocratic at 99 %B,6.0-6.05; 99-15%B, 6.05-7.5; isocratic at 15%B. For the MS method, a Thermo Exploris 480 instrument was used in data-dependent acquisition mode (DDA) with top 5. Heated electrospray ionization (HESI) was used in positive and negative polarities in separate runs (sheath gas = 30, auxiliary gas = 13, spray voltage = 3800 and -3500 V, in positive and negative modes respectively), vaporizer temperature= 350C and ion transfer tube temp = 300°C). Internal mass calibration was used at the start of each run, and mild trapping was set to true to minimize in-source fragmentation (after the ion funnel). MS1 settings were as follows: resolution, 60000, scan range (150-2000 m/z), AGC target (1e6), max. Injection time (10ms). MS2 scan settings were as follows: isolation window (1 m/z), the stepped normalized collision energy was set to 10, 30, and 340 NCE %, Resolution = 15000, max injection time (10msec), AGC target (1e5), RF lens = 50%.

#### Data analysis

Data analysis was performed using Thermo Compound Discoverer software (version 3.3). Lipid identification was conducted by matching spectra against the LipidBlast library, followed by visual inspection of mirror plots (library vs. data) to confirm spectral alignment and the presence of diagnostic fragments. Identification was validated according to the Lipid Standards Initiative guidelines, ensuring compliance with established fragmentation rules. Only lipids that met the criteria for expected fragmentation spectra, mass accuracy threshold, and retention time (RT) pattern were included in the analysis.

Peak areas were normalized using median normalization, and statistical comparisons between groups were performed using a t-test after log transformation. Volcano plots and heatmaps were generated in R using the ggplot2 and ComplexHeatmap packages.

### RNA isolation, reverse transcription, and quantitative PCR

Total RNA was extracted from mouse tissues or zebrafish larvae using the AurumTM Total RNA Fatty and Fibrous Tissue Kit (Bio-Rad, Hercules, CA). To eliminate genomic DNA contamination, DNAse I treatment was performed. Total RNA was then extracted with an RNAqueousR kit (Applied Biosystems, Life Technologies). For cDNA synthesis, 1 µg of RNA was used with the iScript TM cDNA Synthesis Kit (Bio-Rad). Changes in mRNA levels of target genes were determined by relative RT-qPCR using a CFX96TM Real-Time PCR Detection System (Bio-Rad) and iQ TM SYBR Green Supermix (Bio-Rad). Analyses were performed in duplicate with 100nM of both sense and antisense primers in a final volume of 20 µL using iQTM SYBR Green Supermix (Bio-Rad). Specific primers were designed using Primer 3 (**Tables S3 and S4**). PCR conditions were 95°C for 3 min, followed by 40 cycles of 15 sec at 95°C and 30 sec at 60°C. PCR products were sequenced with the BigDye terminator kit (Perkin Elmer Applied Biosystems) using the ABI3100 capillary sequencer (Perkin Elmer Applied Biosystems). Primer efficiency was determined by dilution curves. The program geNorm version 3.4 was used to assess the expression stability of candidate reference genes in kidneys, and six reference genes were selected to calculate the normalization factor. Relative changes in targeted gene expression compared to *Hprt*, *Gapdh*, *Actb*, or *eef1a1a* mRNAs were calculated using the 2^-ΔΔCt^ formula.

### Endocytosis in mouse kidneys and cultured primary PT cells

#### Mouse kidneys

Receptor-mediated endocytosis and fluid-phase endocytosis were monitored in the PT segments of mouse kidneys by measuring the uptake of β-lactoglobulin (L3908, Sigma) and Alexa 647-conjugated dextran (D22914, Thermo Fisher Scientific), respectively. β-lactoglobulin was labelled with Cy5 using a TM2 Ab labelling kit (Amersham) according to the manufacturer’s instructions. Fifteen minutes after tail-vein injection of Cy5-β-lactoglobulin (0.8 mg/kg BW) or Alexa 647-conjugated dextran (1.2 mg/kg BW), all mice were anaesthetized, and their kidneys were harvested and processed for confocal microscopy.

#### Primary cells

Endocytic uptake was monitored in mPTCs by incubating them for 60 minutes at 4°C with specific markers: 20μg/mL BSA-Alexa-Fluor-647 (A34785, Thermo Fisher Scientific), 40μg/mL Lysozyme-647 (provided by Dr. A. Hall, University of Zurich), or 20μg/mL Dextran-Alexa-Fluor-647 (Invitrogen, D22914) in complete HEPES-buffered Dulbecco’s modified Eagle’s medium. After incubation, the cells underwent an acid wash and were warmed to 37°C in a growth medium for 20 minutes before fixation and processing for immunofluorescence analysis.

### Lysosomal degradation

The detection of lysosomal activity was performed in mPTCs by using Bodipy-FL-Pepstatin A (P12271, Thermo Fischer Scientific) to the manufacturer’s specifications. The cells were pulsed with 1μM Bodipy-FL-Pepstatin A in a Live Cell Imaging medium for 1h at 37°C, fixed and subsequently analyzed by confocal microscopy.

### Imaging of LDs and pulse-chase experiments

Cells were incubated for 30 min with 2.5μM BODIPY 493/503 (D3922, Thermo Fisher Scientific) the staining solution prepared in a growth medium. The cells were washed 3 times in PBS, fixed in 4% PFA for 10 min, stained with DAPI for another 5 min, washed in PBS, and mounted in a drop of Prolong® Gold (P36934, Thermo Fisher Scientific). For pulse-chase experiments, palmitate (C16) was conjugated to BSA following the standard protocol from Seahorse Biosciences. BODIPY (D3821, Thermo Fisher Scientific) and PAL were conjugated in a 0.1% FA-free bovine serum albumin solution for 30 min at 37°C. Cells were incubated with 0.5μM BODIPY-PAL for 24 hours and chased in a growth medium for 24 hours. The cells were washed 3 times in PBS, fixed in 4% PFA for 10 min, immunolabelled with a mitochondrial marker (TOMM20) and stained with DAPI, and analysed by confocal microscopy. HCS LipidTOX™ (H34477,

Thermo Fisher Scientific) was used for LD staining. Cells were fixed in 4% PFA for 10 min, stained with DAPI for another 5 min, washed in PBS, and incubated in the dark with HCS LipidTOX™ (1:250) diluted in PBS for 45 min. For pulse-chase experiments, cells were incubated for 5 h with 300 µM OA diluted in culture medium and then chased in starvation medium (RPMI containing 0.1 % FBS) for 16 h in presence of BfnA1(250 nM for 4h) or not. Cells were washed fixed in 4% PFA for 10 min, immunolabelled with an autophagic (Sqstm1) or lysosomal (Lamp1) marker, stained with HCS LipidTOX™ (1:250) for 45 min in the dark and analysed by confocal microscopy.

### Quantification of mitochondrial membrane potential and oxidative stress

Mitochondrial membrane potential (Δψ) was assessed following the manufacturer’s guidelines. Cells were treated with 50 nM Tetramethylrhodamine Methyl Ester Perchlorate (TMRM, T668, Thermo Fisher Scientific) and 1μM MitoTracker Red FM (Invitrogen, M22426) during a 30-minute live cell imaging session at 37°C. After washing, cells were analyzed using confocal microscopy (Leica SP8, Center for Microscopy and Image Analysis, University of Zurich) within a chamber maintained at 37°C and 5% CO2. Fluorescence intensity was quantified using the open-source image processing software Fiji (ImageJ, NIH), as described below.

## Immunofluorescence and confocal microscopy

### Mouse tissue slides

Fresh mouse kidneys were fixed by perfusion with 50-60 mL of 4% PFA (158127, Sigma-Aldrich), followed by dehydration and embedding in paraffin at 58°C. Paraffin blocks were sectioned into consecutive 5 µm-thick slices using a Leica RM2255 rotary microtome (Thermo-Fisher Scientific) on Superfrost Plus glass slides (Thermo-Fisher Scientific). Before staining, slides were deparaffinized in Xylenes (534056, Sigma-Aldrich) and rehydrated. Antigen retrieval was performed by heating the slides at 95°C for 10 min in a 10 mM sodium citrate buffer (pH 6.0). Slides were then quenched with 50 mM NH4Cl, blocked with 3% BSA in PBS Ca/Mg (D1283, Sigma-Aldrich) for 30 min, and incubated with primary antibodies diluted in blocking buffer overnight at 4°C. After two washes in 0.1% Tween 20 (v/v in PBS), slides were incubated with fluorophore-conjugated Alexa secondary antibodies (Invitrogen) diluted in a blocking buffer at room temperature for 1 hour. They were subsequently counterstained with 1µg Biotinylated Lotus Tetragonolobus Lectin (LTL; B-1325, Vector Laboratories) and 1µM 4’,6-Diamino-2-phenylindole dihydrochloride (DAPI; D1306, Thermo Fisher Scientific). Slides were mounted in Prolong Gold Anti-fade reagent (P36930, Thermo Fisher Scientific) and analysed by confocal microscopy using a Leica SP8 confocal laser scanning microscope (Center for Microscopy and Image Analysis, University of Zurich).

### Primary cultured cells

The cells were fixed for 10 minutes with 4% PFAin PBS, quenched with 50 mM NH_4_Cl, and permeabilized for 20 minutes in a blocking buffer solution (pH 7.4) containing 15 mM glycine (Carl Roth, 3908.2), 0.05% saponin (Sigma-Aldrich 47,036), 0.5% BSA (Carl Roth, 1ET6.3), 50 mM NH_4_Cl (Sigma-Aldrich, A9734) dissolved in PBS. Subsequently, cells were incubated overnight with the appropriate primary antibodies at 4°C. After repeated washing with PBS, the slides were incubated for 45 minutes with the suitable fluorophore-conjugated Alexa secondary antibodies (Invitrogen), counterstained with 1µM DAPI for 5 minutes, mounted with Prolong Gold Anti-fade reagent, and analyzed using a Leica SP8 confocal laser scanning microscope or a Zeiss LSM 980 confocal laser-scanning microscope equipped with an Airyscan 2 multiplex system (Center for Microscopy and Image Analysis, University of Zurich).

### Image processing and quantification

Images were acquired with a Leica APO 63x NA 1.4 oil immersion objective at a resolution of 1,024 x 1,024 pixels or a Zeiss 63x 1.40 oil immersion objective with a resolution of 3,072 x 3,072 pixels. Micrographs were processed using Adobe Photoshop CS5 software (Adobe Systems Inc., San Jose, USA). Quantitative image analysis involved selecting randomly 5-10 visual fields per slide containing at least 3-5 proximal tubules (LTL-positive or AQP1-positive as indicated) or 10-15 cells.

Setting parameters were consistently applied across analyses, including pinhole size, laser power, and offset gain, ensuring detector amplification remained below pixel saturation. Subcellular structure analyses were performed using the open-source software CellProfiler^49^, specifically utilising the “Measure-Object-Intensity-Distribution” module to quantify the number of LD, Pepstatin A- and Map1Lc3b- or Sqstm1- or BSA- or Lysozyme-positive structures. For measurements such as tubular height in mouse kidneys, mitochondrial morphology, and mitochondrial membrane potential, image analysis was carried out using Fiji v.2.0.0 (ImageJ, NIH). For the quantification of colocalization, images were imported into FIJI v.2.0.0-rc-69/1.52i, and individual channels were thresholded independently to eliminate background. Colocalization was assessed on individual cells with the JACOP plugin and Mander’s coefficients. The number of cells and/or fields of view used per condition and the number of biologically independent experiments are indicated in the figure legends.

### Imaging of zebrafish larvae

Quantitative analysis of fluorescent signals involved examining an entire pronephric tubule from each zebrafish larva, maintaining consistent setting parameters throughout the process. A multi-photon fluorescence microscope (Leica SP8 MP DIVE Falcon; Leica Microsystems, Heerbrugg, Switzerland) equipped with a 25/1.0 NA water immersion objective (HC IRAPO L, Leica) was used for capturing high-resolution images. The excitation wavelength was generated by a 1040 nm tunable laser for mCherry imaging. A light sheet fluorescence microscope (ZEISS Light sheet Z.1, Carl Zeiss Microscopy, Jena, Germany) equipped with a 20×/1.0 NA water immersion objective (W Plan APO, Zeiss) was used for detecting the human EGFP-ATG7 in mutant zebrafish larvae. Data acquired underwent processing with Huygens software (Scientific Volume Imaging, Hilversum, The Netherlands) for deconvolution, followed by segmentation using Ilastik software (EMBL, Heidelberg, Germany). Quantitative analysis of fluorescence intensity, vesicle number, and vesicle area in each image stack was conducted using Fiji (ImageJ, NIH), an open-source image processing software. For the segmentation of Map1Lc3b spots, image stacks were analysed using the object analyzer plugin within Huygens. This involved setting seed and threshold levels using algorithms, employing watershed segmentation, and filtering objects to isolate Map1Lc3b spots. A lower threshold was established to identify the entire kidney volume as a single object. The number of spots and extracted morphological parameters (such as length, volume, and sphericity) were exported in Excel format for subsequent statistical analysis. Surface images generated were used to create 3D volume-rendering images for visualisation purposes.

### Immunohistochemistry

Paraffin kidney sections were rehydrated, and antigen retrieval was performed either by heat-induced treatment in 10 mM sodium citrate buffer with 0.05% Tween-20 at pHL6.2 or by proteinaseLK treatment. Endogenous peroxidase activity was blocked using a peroxidase-blocking buffer for 15Lminutes at room temperature. Sections were then blocked in a solution containing 1% BSA, 0.2% fish-skin gelatine, 0.2% Triton-X-100, and 0.05% Tween-20 in PBS for 1 hour at room temperature. Following blocking, sections were incubated overnight at 4°C with a primary antibody against Cre (Cell Signaling 15036S, 1:200). The next day, sections were incubated with biotinylated anti-mouse IgG (H+L) secondary antibody (Vector Laboratories, BA-9200-1.5, diluted 1:1,000) or anti-Rabbit secondary antibody. Each staining was visualized using the ABC Kit Vectastain Elite (Vector, PK6100) and DAB substrate (Dako and Vector Laboratories).

### Histology and semiquantitative analysis of fibrosis

Picrosirius red staining (ab150681, Abcam, Cambridge, UK) was performed following the manufacturer’s instructions. Samples were rehydrated and incubated for 1 hour in picrosirius red solution. After incubation, samples were briefly rinsed in 0.5% acetic acid, dehydrated in absolute ethanol, cleared, and mounted. Evaluation of picrosirius red staining was conducted using a Zeiss AxioScanner.Z1 (Carl Zeiss, Oberkochen, Germany). Full scans of 24-week-old Atg7 kidneys were acquired at a resolution of 200× using a Zeiss Axio Scan.z1 slide scanner equipped with a Hitachi HV-F202FCL digital video camera (Hitachi, Tokyo, Japan). Images were captured and processed using ZENblue software (Carl Zeiss SpA).

## Electron microscopy

### Primary cells

Cultured mPTCs were fixed in 2.5% glutaraldehyde in 0.1 M cacodylate buffer (pH 7.4) at 37°C for 1 hour, followed by post-fixation in 1% osmium tetroxide and 1.5% potassium ferrocyanide in 0.1 M cacodylate on ice for 1 hour. After washing in distilled water, samples were stained with 0.5% uranyl acetate in water overnight at 4°C. Dehydration was performed in a graded ethanol series, followed by embedding in Epon 812 and curing at 60°C for 48 hours. Ultrathin sections (70–90nm) were cut using a Leica Ultracut UCT ultramicrotome, collected on copper grids, and stained with uranyl acetate and Sato’s lead citrate. Imaging was conducted using a FEI Talos 120kV transmission electron microscope with a 4k×4K Ceta CMOS camera. Autophagic vacuoles (AVs) were identified and categorised as autophagosomes or autolysosomes, based on established criteria.^17^ At least 20 randomly selected cellular profiles were analysed using ImageJ software to quantify AVs by calculating the percentage of cytosolic area occupied.

### Zebrafish larvae

Animals were fixed in 2.5% glutaraldehyde and 1.6% paraformaldehyde in 0.1 M cacodylate buffer (pH 7.4) overnight after tail cutting. After rinsing in 0.1 M cacodylate buffer, larvae were post-fixed in 1% osmium tetroxide in cacodylate buffer for 40 minutes at room temperature and stained with 1% aqueous uranyl acetate for 1 hour at room temperature. Samples were dehydrated through a graded ethanol series, infiltrated with Epon812 at 60°C for 28 hours, and then sectioned into 350 nm semi-thin sections using a Leica EM FCS ultramicrotome. Sections were stained with Toluidine blue solution. Ultra-thin sections of 60 nm were collected onto formvar-coated copper grids, stained with lead phosphate dilution in water, and examined using an electron microscope (Philips CM100) at 80kV.

### Sample preparation and immunoblotting

Proteins from animal tissues or cultured mPTCs were extracted using a lysis buffer containing protease inhibitors (Roche) and phosphatase inhibitors (PhosSTOP, Sigma). The lysates were sonicated and centrifuged at 10,000 × g for 10 min at 4°C. Protein concentrations were normalised (20 µg/lane), and samples were mixed with 4x Laemmli Sample Buffer (BioRad, 1610747) and separated by SDS-PAGE under reducing conditions. Proteins were then transferred onto PVDF membranes, which were blocked with 5% non-fat milk (Bio-Rad Laboratories, 1706404), followed by overnight incubation at 4°C with primary antibodies. After washing, membranes were incubated with peroxidase-labelled secondary antibodies, and protein bands were visualised using enhanced chemiluminescence (Millipore, WBKLS0050) and imaged with a ChemiDoc MP imaging system (Bio-Rad Laboratories). Signal intensities were quantified by measuring the relative density of each band normalised to β-actin using ImageJ software.

### Extracellular flux analysis and metabolic measurements

OCRs in mPTCs were assessed using XFp Extracellular Flux Analyzers from Agilent Seahorse Biosciences. Cells were cultured in XF-Base Medium (non-buffered RPMI 1640 supplemented with 2 mM L-glutamine, 1 mM sodium pyruvate, and 10 mM glucose, pH 7.4). OCR was measured under basal conditions and after the sequential addition of specific inhibitors using the XFp Cell Mito Stress Test Kit (Agilent Seahorse Biosciences). The inhibitors included 2μM Oligomycin (to inhibit ATP synthase), 0.5μM FCCP (to uncouple mitochondrial respiration), and 1μM Rotenone/Antimycin A (to inhibit complex I and III of the mitochondrial electron transport chain). All OCR measurements were normalised to cell numbers using the TC10TM automated cell counter from Bio-Rad. This method allows for the assessment of mitochondrial function and cellular respiration dynamics in mPTCs.

### Cell proliferation

To assess the rate of proliferation, we used the Click-iT Plus EdU Alexa Fluor 488 Imaging Kit (C10637 Life Technologies). Cells were incubated with EdU solution for 16 h at 37 °C and processed following the manufacturer’s instructions. DAPI (4′,6-diamino-2-phenylindole dihydrochloride) was used to counterstain nuclei. Analyses were performed by using the “Cytoplasm-Nucleus Translocation Assay” (CellProfiler^49^) to score the number of BrdU-positive nuclei.

### Analysis of TCGA gene expression on cell renal cell carcinoma samples and survival curves

The Genebass browser (https://app.genebass.org/) provided detailed coefficients and p-values from burden tests assessing the association between Chapter II neoplasms (labelled with ICD10 codes) and pLOF alleles in *ATG7*. The entire cohort for clear cell renal cell carcinoma (KIRC) was downloaded from TCGA (The Tissue Cancer Genome Atlas). It included transcriptomic (in FPKM) and clinical data from 537 patients. In the first step of filtering, data from patients with insufficient clinical data for survival analysis were removed. Additionally, data from patients who had multiple RNA-seq performed on the same sample from tumour tissue were excluded. 525 patients remained after filtering, and 72 had available data from adjacent normal tissue samples to the primary tumour (Normal Tissue). The total number of samples obtained amounts to 597 RNA-seq samples from 525 patients. The entire data analysis was performed locally with Python (v. 3.11.5) using Pandas (v. 2.2.3), Numpy (v. 1.24.3), and glob libraries. The division into tertiles was performed using Numpy, considering only the gene expression of *ATG7* in the primary tumour data. The normal tissue samples were assigned to the tertile of the corresponding primary tumour. Survival analyses were performed considering only the 525 transcriptomic data from tumour tissues (Primary Tumour). These analyses were performed using the lifelines library (v. 0.30.0). Violin plots were generated using the Seaborn (v. 0.11.2) and Matplotlib (v. 3.7.2) libraries, considering the transcriptomic data from both the 72 normal tissue samples and the 525 primary tumour samples.

### Data Collection and Integration Using PandaOmics AI Platform

The PandaOmics AI platform (Insilico Medicine, Hong Kong, China) is a cloud-based software tool that leverages artificial intelligence and bioinformatics to analyse multimodal omics and biomedical text data for the discovery of therapeutic targets and biomarkers.^50^ This platform was utilised to integrate omics datasets from the Gene Expression Omnibus and ArrayExpress, combining these with samples of clear cell renal cancer tissues and adjacent normal kidney samples. Gene expression data were uploaded to PandaOmics, normalised and pre-processed following the standard protocols provided by the platform.

### Differential expression and pathway enrichment analyses

The ccRCC gene signature was derived through a PandaOmics meta-analysis, which incorporated data from seven comparisons across seven distinct transcriptomic datasets. Differential expression analysis was conducted in PandaOmics utilising the limma R package, with each dataset processed following standard limma protocols. Gene-wise p-values were adjusted using the Benjamini-Hochberg correction method. PandaOmics calculated combined log-fold changes (LFC) across transcriptomics datasets using the ‘Expression’ feature. This feature calculates combined LFC values and Q-values across all datasets used for the analysis, employing min-max normalisation for LFC values and Stouffer’s method for p-value aggregation, followed by additional FDR correction. All differentially expressed genes (DEGs) were subsequently downloaded and used to construct the final gene expression signature.

Pathway enrichment analysis was performed with the GSEApy package prerank function according to standard protocols. For the prerank protocol, each gene was ranked according to the value derived from the multiplication of LogFoldChange and -log10(p-value) obtained from differential expression analysis results. MSigDB and Reactome databases were selected as the gene set from the GSEApy internal library for signalling pathway enrichment analysis.

### Single-cell RNA sequencing analysis

Single-cell RNA sequencing (scRNA-seq) data from 167,283 cells from multiple tumour regions, lymph nodes, normal kidneys, and peripheral blood of two immune checkpoint blockade (ICB)-naïve and four ICB-treated ccRCC patients were collected from the Krishna et al.^40^ Cell type annotations were derived from the original publication, as well as normalised counts, and dimensionality reduction post-batch correction for the UMAP. Differential expression between ccRCC and control samples for each cell type was calculated using the tl.rank_genes_groups function (method=’Wilcoxon’). ATG7 expression levels (non-zero values) were plotted with corresponding p-values (threshold: 0.05) using the Plotly package.

### Statistics and reproducibility

The data were presented as mean ± SEM. Statistical analyses were conducted using appropriate tests: one-way analysis of variance (ANOVA) with Tukey’s, Sidak’s, or Dunnett’s post hoc tests for multiple comparisons between experimental groups, as required. For comparisons between two groups, two-tailed Student’s t-tests were used as appropriate. Normality was assessed using the D’Agostino and Pearson omnibus test. Non-parametric data were analysed using the Kruskal-Wallis test with Dunn’s multiple comparison correction. Survival estimates were generated using the Kaplan–Meier method and compared using log-rank tests. Outliers were identified using the ROUT method (Q=1%). The sample size for each experimental group is detailed in the figure legends. Statistical significance levels are denoted by symbols, and the exact P-values are provided in the figure legends along with the specific statistical tests used. Each experiment was independently performed at least two to three times unless otherwise specified in the figure legends, and successful replication was achieved in all instances. Experimenters were not blinded to allocation during most experiments, except for experiments involving zebrafish larvae. GraphPad Prism software version 9.4.1 (GraphPad Software) was used for all statistical analyses and figure generation.

## Data Availability

All data produced in the present study are available upon reasonable request to the corresponding author.

## Acknowledgements

We thank Nadine Nägele and Marco Garbelli for technical help, the Center for Microscopy, and Image Analysis at the University of Zurich for providing equipment and confocal microscopy assistance, and the Functional Genomics Center at the University of Zurich for assistance. We thank C.W. Li, P. Nanni, and S. Streb (Functional Genomics Center Zurich, University of Zurich) for their advice and constructive suggestions. We acknowledge Euro-Bioimaging (www.eurobioimaging.eu) for providing access to imaging technologies and services via the Italian Node (ALEMBIC, Milan, Italy). We are grateful to the Cystinosis Research Foundation [Irvine, CA, USA; project grants CRFS-2017-007 (O.D. and A.L.), CRFS-2020-005 (O.D. and A.L.) and CRFS-2023-003 (O.D. and A.L.)], the Swiss National Science Foundation (project grant 310030_215715 to A.L.; and project grant 310030_204470 to C.M.), the University Research Priority Program of the University of Zurich (URPP) ITINERARE-Innovative Therapies in Rare Diseases (O.D. and A.L.). R.W.T. and R.M. are funded by the Wellcome Centre for Mitochondrial Research (203105/Z/16/Z), the Mitochondrial Disease Patient Cohort (UK) (G0800674), the Medical Research Council (MR/W019027/1), the Lily Foundation, the UK NIHR Biomedical Research Centre for Ageing and Age-related disease award to the Newcastle upon Tyne Foundation Hospitals NHS Trust and the UK NHS Highly Specialised Service for Rare Mitochondrial Disorders of Adults and Children. R.W.T. also receives additional funding from LifeArc and the Pathological Society. Postdoc Mobility-Stipendien of the Swiss National Science Foundation Grants P2ZHP3_195181 and P500PB_206851, Kidney Research UK Grant Paed_RP_001_20180925 support EO. Discovery Grant (NSERC), a Project Grant (CIHR) and a Research Chair in Precision Medicine (J. Louis Levesque Foundation – NB Research) support LRL.

## Authors contributions

A.L. and O.D. conceived and supervised the study. D.N., S.K., O.D., and A.L. designed the experiments. D.N., S.K., and L.P. conducted immunoblotting analysis, RNA interference, gene expression experiments, confocal microscopy, live-cell imaging, molecular biology studies, and data analysis. L.P. conducted confocal microscopy studies of lipid droplet biology and lipophagy flux, analysed and interpreted the data. Z.C. generated the *atg7* knockout zebrafish model and, together with D.N., performed complementation experiments in zebrafish larvae and data analysis. A.R. conducted electron microscopy studies and performed data analysis. V.B. performed deconvolution of MAP1BLC3-positive autophagosome structures in the zebrafish pronephros and analysed the data. M.B. and P.K. conducted differentiation and proliferation studies and data analysis. M.Z., M.R., and A.O. performed metabolomics- and lipidomics-based studies and data analysis. R.M., R.W.T., A.M.S., E.G.O., and J.A.S. provided urine samples from two patients with loss-of-function *ATG7* variants and intellectual feedback. S.N. supervised D.N. in zebrafish swimming behaviour analysis. C.M. provided *Fip200* and *Atg5* floxed mouse lines and intellectual feedback. A.U. and M.K. analysed publicly available microarrays and bulk and single-cell RNA sequencing of ccRCC patient samples using artificial intelligence and data analysis. R.P., F.C., and E.R. performed genome-wide and phenome-wide association studies, gene expression, and survival analyses in ccRCC patients, along with statistical testing. A.M.O. carried out lipid droplet, autophagy, and bioenergetic studies in worms and performed data analysis. L.R.L. supervised A.M.O. and provided intellectual feedback. A.L. and O.D. drafted and edited the manuscript, incorporating input and comments from all authors. All authors approved the final version of the manuscript.

## References

1. Palm, P. & Thompson, C.B. Nutrient acquisition strategies of mammalian cells. Nature 546, 234–242 (2017).

2. Keller, S. A. et al. Drug discovery and therapeutic perspectives for proximal tubulopathies. Kidney International 6, 1103–1112 (2023).

3. Nielsen, R., Christensen, E.I., Birn, H. Megalin and cubilin in proximal tubule protein reabsorption: from experimental models to human disease. Kidney International 89, 58–67 (2016).

4. Cullen, P. J. & Steinberg, F. To degrade or not to degrade: mechanisms and significance of endocytic recycling. Nature Reviews Molecular Cell Biology 19, 679–696 (2019).

5. Mizushima, N. & Levine, B. Autophagy in human diseases. New England Journal of Medicine 381, 1564–1576 (2020).

6. Festa, B. P. et al. Impaired autophagy bridges lysosomal storage disease and epithelial dysfunction in the kidney. Nature Communications 9, 161 (2018).

7. Berquez, M. et al. Lysosomal cystine export regulates mTORC1 signalling to guide kidney epithelial cell fate specialization. Nature Communications 14, 3994 (2023).

8. Yamamoto, H. et al. Autophagy genes in biology and disease. Nature Reviews Genetics 24, 382–400 (2023).

9. Nakatogawa, H. Mechanisms governing autophagosome biogenesis. Nature Reviews Molecular Cell Biology 21, 439–458 (2020).

10. Komatsu, M., Waguri, S., Ueno, T. et al. Impairment of starvation-induced and constitutive autophagy in Atg7-deficient mice. Journal of Cell Biology 169, 425–434 (2005).

11. Klionsky, D. J. et al. Guidelines for the use and interpretation of assays for monitoring autophagy (4^th^ edition). Autophagy 17, 1–382 (2021).

12. Collier, J. J. et al. Emerging roles of ATG7 in human health and disease. EMBO Molecular Medicine 13, e14824 (2021).

13. Komatsu, M. et al. Loss of autophagy in the central nervous system causes neurodegeneration in mice. Nature 441, 880–884 (2006).

14. Kimura, T. et al. Autophagy protects the proximal tubule from degeneration and acute ischemic injury. Journal of American Society of Nephrology 22, 902–913(2011).

15. Liu, S. et al. Autophagy plays a critical role in kidney tubule maintenance, aging, and ischemia-reperfusion injury. Autophagy 8, 826–837 (2012).

16. Wang, Z. M. et al. Specific metabolic rates of major organs and tissues across adulthood: evaluation by mechanistic model of resting energy expenditure. Am. J. Clin. Nutr. 92: 1369–1377 (2010).

17. van der Rijt, S. et al. Immunometabolic rewiring of tubular epithelial cells in kidney disease. Nature Reviews Nephrology 18, 588–603 (2022).

18. Bhargava, P. and Schnellman, R.G. Mitochondrial energetics in the kidney. Nature Reviews Nephrology 13, 629–646 (2017).

19. Dijon, P. et al. The nuclear receptor ESRRA protects from kidney disease by coupling metabolism and differentiation. Cell Metabolism 33, 379–384 (2021).

20. Doke, T., Mukerjee, S., Mukhi, D. et al. NAD+ precursor supplementation prevents mtRNA/RIG-I-dependent inflammation during kidney injury. Nature Metabolism 5, 414–430 (2023).

21. Collier, J. J. Et al. Developmental consequences of defective ATG7-mediated autophagy in humans. New England Journal of Medicine 384, 2406–2417(2021).

22. Iwano, M. et al. Evidence that fibroblasts derive from epithelium during tissue fibrosis. Journal of Clinical Investigation 110, 341–350 (2002).

23. Perez Bay, A. E., et al. The fast-recycling receptor Megalin defines the apical recycling pathway of epithelial cells. Nature Communications 7, 11550 (2016).

24. Jo, Y. S. et al. Phosphorylation of the nuclear receptor corepressor 1 by protein kinase B switches its corepressor targets in the liver of mice. Hepatology 62, 1606–1618 (2015).

25. Liu, G. Y. & Sabatini, D. M. mTOR at the nexus of nutrition, growth, ageing and disease. Nature Reviews Molecular Cell Biology 21, 183–203 (2020).

26. Singh, R. et al. Autophagy regulates lipid metabolism. Nature 458, 1131–1135 (2009).

27. Nguyen, T. B., et al. DGAT1-dependent lipid droplet biogenesis protects mitochondrial function during starvation-induced autophagy. Developmental Cell 42, 9–21 (2017).

28. Morant-Ferrando, B., et al. Fatty acid oxidation organizes mitochondrial supercomplexes to sustain astrocytic ROS and cognition. Nature Metabolism 5, 1290–1302 (2023).

29. Cunha, L. D. et al. LC3-associated phagocytosis in myeloid cells promotes tumor immune tolerance. Cell 175, 429–441 (2018).

30. Heckmann, B. L. et al. LC3-associated endocytosis facilitates beta-amyloid clearance and mitigates neurodegeneration in murine Alzheimer’s disease. Cell 178, 536–551(2019).

31. Kumar, A. V. et al. Lipid droplets modulate proteostasis, SQST-1/SQSTM1 dynamics, and lifespan in C. elegans. iScience 26, 107960 (2023).

32. MacRoe, C. A. & Peterson, R. T. Zebrafish as tools for drug discovery. Nature Reviews Drug Discovery 14, 721–730 (2015).

33. Chen, Z. et al. Transgenic zebrafish modelling low-molecular-weight proteinuria and lysosomal storage diseases. Kidney International 97, 1150–1163 (2020).

34. Verma, et al. Diversity and scale: Genetic architecture of 2068 traits in the VA Million Veteran Program. Science 385, 275 (2024).

35. Levine, B. & Kroemer, G. Biological functions of autophagy genes. Cell 176, 11–42 (2019).

36. Pan, X. et al. Molecular subtyping and characterisation of clear cell renal cell carcinoma by tumour differentiation trajectories. iScience 26, 108370 (2023).

37. Tan, S. K. et al. Fatty acid metabolism reprogramming in ccRCC: mechanisms and potential targets. Nature Reviews Urology 20, 48–59 (2023).

38. Karczewski, K. J., et al. Systematic single-variant and gene-based association testing of thousands of phenotypes in 394,841 UK Biobank exomes. Cell Genomics 2, 100168 (2022).

39. Tang, Z. et al. GEPIA2: an enhanced web server for large-scale expression profiling and interactive analysis. Nucleic Acids Research 47, W556–560 (2019).

40. Krishna, C. et al. Single-cell sequencing links multiregional immune landscapes and tissue-resident T cells in ccRCC to tumour topology and therapy efficacy. Cancer Cell 39, 662–677.e6 (2021).

41. Galluzzi, L. & Green, D. Autophagy-Independent Functions of the Autophagy Machinery. Cell 17, 1682–1699 (2019).

42. Mitrofanova, A., Merscher, S., and Fornoni, A. Kidney lipid dysmetabolism and lipid droplet accumulation in chronic kidney disease. Nature Reviews Nephrology 19, 629–645 (2023).

43. Miceli, C., et al. The primary cilium and lipophagy translate mechanical forces to direct metabolic adaptation of kidney epithelial cells. Nature Cell Biology 22, 1091–1102 (2020).

44. Geng, T.T., et al. Nuclear magnetic resonance-based metabolomics and risk of CKD. Am J Kidney Dis. 83, 9–17 (2024).

45. Ganley, I. The Importance of Being Autophagic. N. Engl. J. Med. 384, 2449–2450 (2021). .

46. Olzmann, J.A., & Carvalho, P. Dynamics and functions of lipid droplets. Nat. Rev. Mol. Cell Biol. 20, 137–155 (2019).

47. Liu, Y. et al. Hydroxysteroid dehydrogenase family proteins on lipid droplets in bacteria, C. elegans, and mammals. Biochim. Biophys. Acta Mol. Cell. Biol. Lipids 1863, 881–894 (2018).

48. Alshehry, Z. H., Barlow, C. K., Weir, J. M.; Zhou, Y., McConville, M. J., Meikle, P. J. An efficient single-phase method for extracting plasma lipids. Metabolites 5, 389–403 (2015).

49. Carpenter, A. E., Jones, T. R., Lamprecht, M. R. et al. CellProfiler: image analysis software for identifying and quantifying cell phenotypes. Genome Biol. 7, R100 (2006).

50. Kamya, P. et al. PandaOmics: An AI-driven platform for therapeutic target and biomarker discovery. J. Chem. Inf. Model. 64, 3961–3969 (2024).

