## Supplementary information for "Autophagy regulator ATG7 links lipid metabolism to cell-fate decisions in kidney tubule health and disease"

Nieri et al.

**Supplemental information**

Figures S1-S17

Table S1-S4

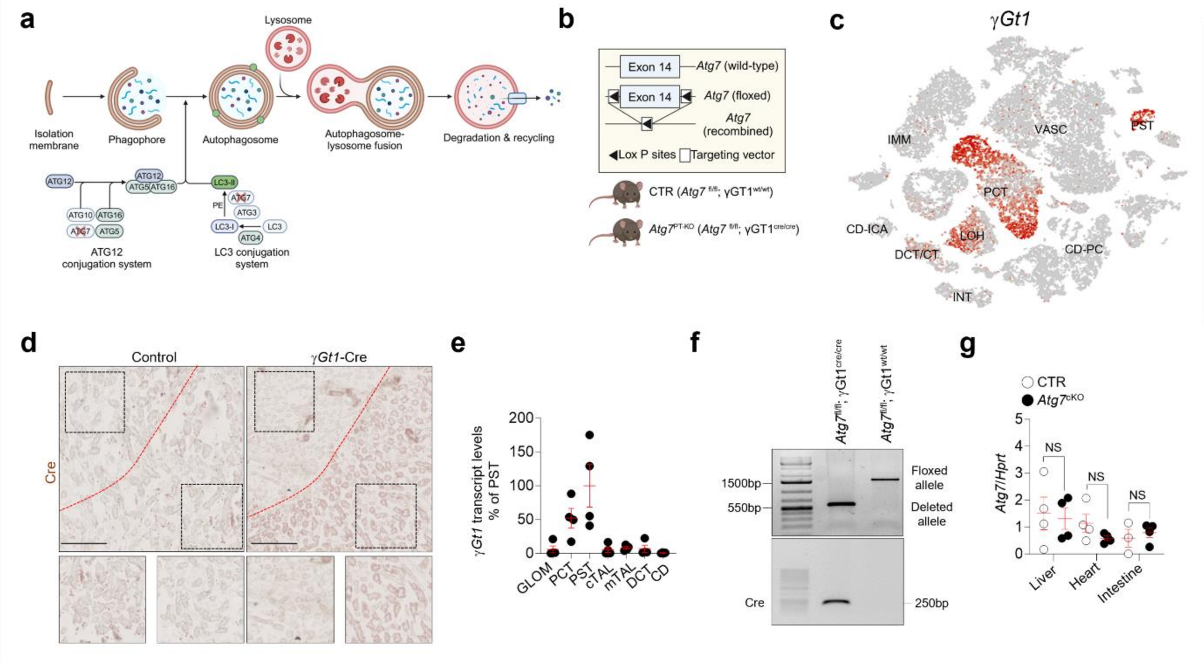


**Supplementary Fig. 1. Validation of *Atg7*$^{fl/fl}$ and γ*Gt1*-Cre mouse lines.** (**a**) Role of ATG7 in autophagy. Autophagy is activated by stress signals that initiate the process through the ULK1 complex, which integrates upstream signals to form a phagophore. This double-membrane structure incorporates ATG5-ATG12 and ATG8 proteins like Maplc3b-II, maturing into an autophagosome. ATG7 is essential for the conjugation systems that produce these components. The mature autophagosome fuses with the lysosome for the degradation of the cargo. (**b**) Diagram showing the strategy to generate *Atg7* floxed alleles and a mouse model with PT-specific (*Atg7*$^{fl/fl}$; γ*Gt1*$^{cre/cre}$) deletion of *Atg7*. (**c**) The t-SNE plot of the metabolic gene expression profile of γ*Gt1* in various cell types of the mouse kidneys based on data from Ransick et al (Dev. Cell 2019). CD: collecting duct (PC: principal cells, IC: intercalated cells (type A, Type B); DCT/CT: distal convoluted tubule/connecting tubule; INT: interstitial cells; LOH: loop of Henle; VASC: vasculature; IMM: immune cells. (**d**) mRNA levels of γ*Gt1* measured by RT-qPCR in microdissected nephron segments, n = 4 replicates from 2 biologically independent experiments. GLOM: glomerulus; PCT: proximal convoluted tubule; PST: proximal straight tubule; cTAL: cortical thick ascending limb; mTAL: medullary thick ascending limb; DCT: distal convoluted tubule; and CD: collecting duct. (**e**) Representative images of Cre expression in whole-kidney sections from γ*Gt1*-Cre mice. Insets show a high-magnification view of the marked area (black squares). Red lines indicate the boundary between the cortex and medulla. (**f**) Genotyping and PCR results from *Atg7*$^{fl/fl}$; γ*Gt1*$^{wt/wt}$ and *Atg7*$^{fl/fl}$; γ*Gt1*$^{cre/cre}$ mice using primers to detect floxed, deleted alleles and Cre recombinase. (**g**) mRNA levels of the indicated gene in the liver, heart, and intestine of 12-week-old *Atg7* mice, n = 4 biologically independent samples per genotype. Plots represent mean ± SEM. Statistics were calculated by an unpaired two-tailed Student's t-test. NS, not significant. Scale bar, 50µm in (**c**).

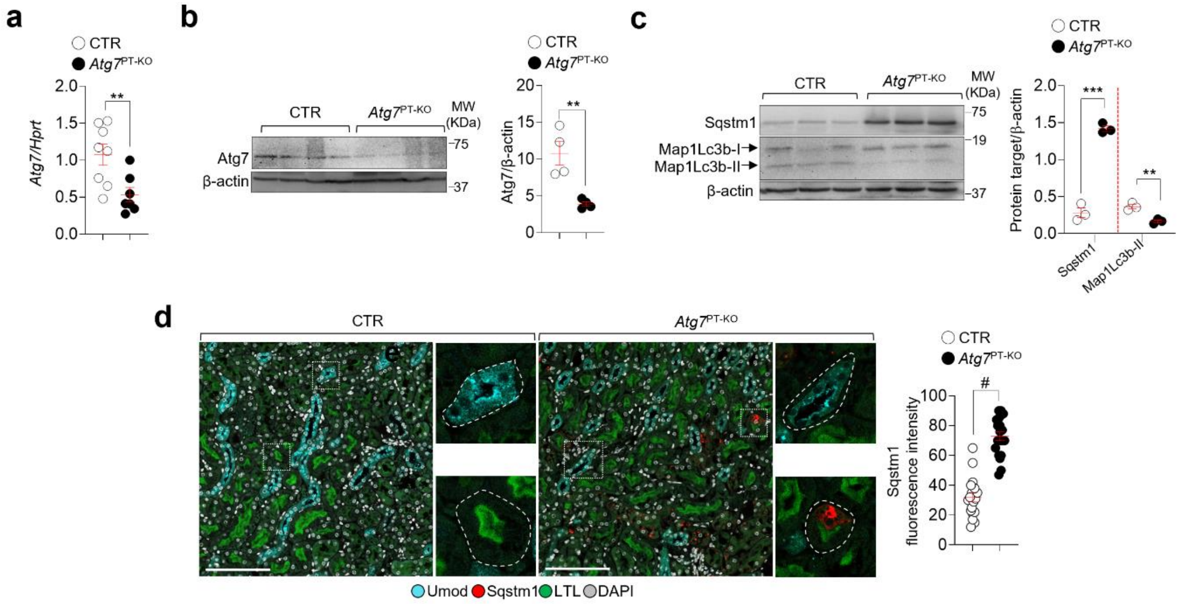


**Supplementary Fig. 2**. **ATG7 deficiency impairs autophagy in the PT of mouse kidneys.** (**a**) mRNA levels of the indicated genes in the kidney cortex of 12-week-old CTR (n = 8) and *Atg7*$^{PT\text{-}KO}$ (n = 7) mice. (**b-c**) Immunoblots and quantification of the indicated proteins in kidney cortex samples from 12-week-old CTR mice and *Atg7*$^{PT\text{-}KO}$; n = 4 animals and 3 animals per genotype in **b** and **c**, respectively. (**d,** left) Confocal imaging of whole-kidney sections immunostained with antibodies against Umod (cyan) and Sqstm1 (red). The PT segments and nuclei counterstained with Lotus tetragonolobus lectin (LTL; green) and 4′,6-diamidino-2-phenylindole (DAPI; grey), respectively. (**d,** right) Quantification of Sqstm1 fluorescence intensity, n = 30 PT segments from 3 animals per genotype. Bar graphs show mean ± SEM. Statistics were calculated using unpaired two-tailed Student's *t*-test, ** P < 0.01, ***P < 0.001 and $^{\#}$P < 0.0001 relative to CTR. Scale bar, 50μm.

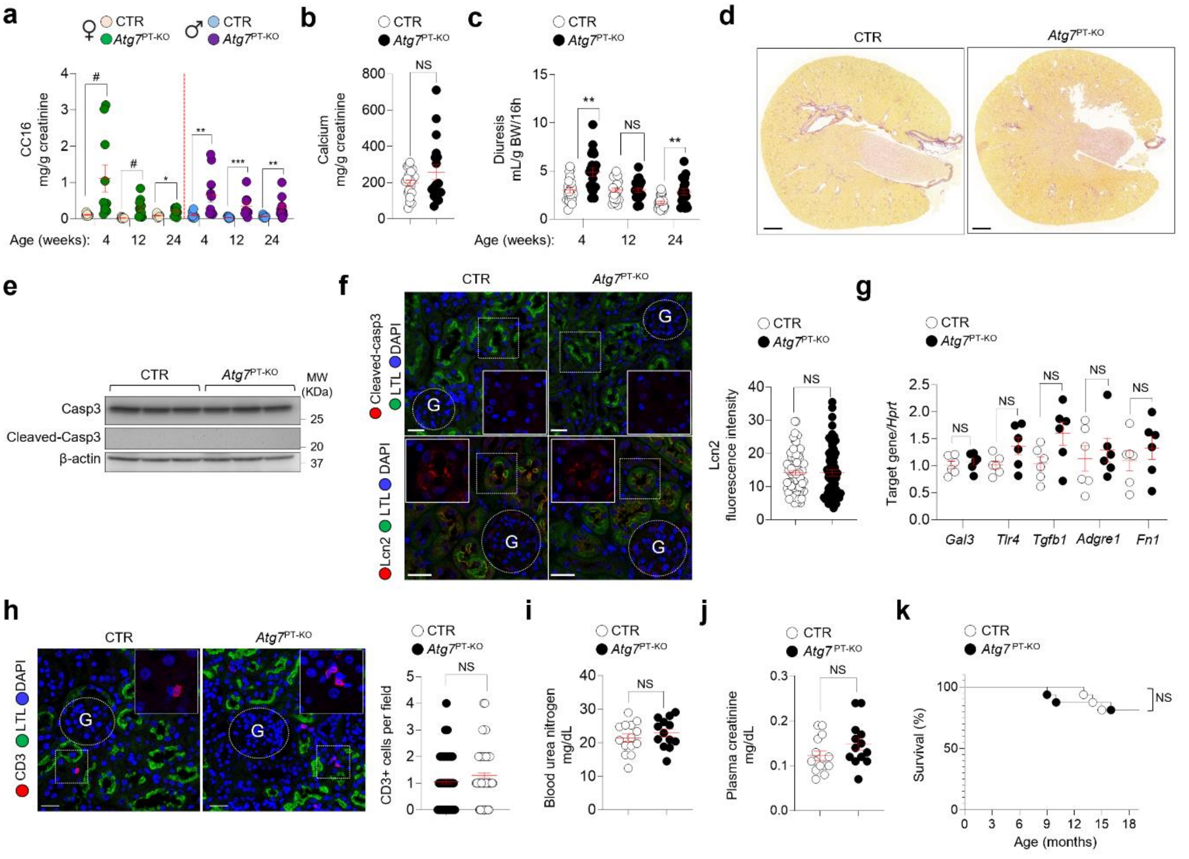


**Supplementary Fig. 3. ATG7 loss induces phenotypic changes in the mouse kidney.** (**a**) Urinary levels of low-molecular-weight protein CC16 at the indicated times in both female and male *Atg7* mice; n = 10 animals per gender and genotype. (**b**) Urinary calcium concentrations in 24-week-old *Atg7* mice; n = 20 animals per genotype. (**c**) Overnight urine volume (mL/16h) normalized to body weight, measured at 4, 12 and 24 weeks; n = 20 animals at 4 weeks of age; n = 20 CTR and n = 19 *Atg7*$^{PT\text{-}KO}$ mice at 12 weeks; n = 20 animals at 24 weeks of age. (**d**) Representative micrographs of Picrosirius Red staining in 24-week-old mice. (**e**) Immunoblots of the indicated proteins, n = 3 biologically independent experiments. (**f**) Representative micrographs of whole-kidney sections immunostained for cleaved caspase 3 (top panel, red) and lipocalin 2 (Lcn2; bottom panel, red) and quantification of Lcn2 fluorescence intensity in the kidney PT of 24-week-old mice. PT segments counterstained with Lotus tetragonolobus lectin (LTL, green) and nuclei with 4′,6-diamidino-2-phenylindole (DAPI, blue); n = 20 PT segments pooled from 4 biologically independent experiments. (**g**) mRNA levels of the indicated genes in the kidney cortex of 12-week-old mice, n=6 animals per genotype. (**h**) Representative micrographs and quantification of the number of CD3$^+$ cells (red) in the kidneys of 12-week-old mice. PT segments counterstained with LTL (green) and nuclei with DAPI (blue); n = 96-98 areas pooled from 4 biologically independent experiments. (**i-j**) Plasma levels of (**i**) blood urea nitrogen (BUN) and (**j**) creatinine in 24-week-old mice n = 14 CTR and 13 *Atg7*$^{PT\text{-}KO}$ mice. (**k**) Kaplan-Meier curves comparing CTR and *Atg7*$^{PT\text{-}KO}$ mice, n = 16 mice per genotype. Plots represent mean ± SEM. Statistics calculated by two-tailed unpaired Student's t-test, *P < 0.05, **P < 0.01, ***P < 0.001, $^{\#}$P < 0.0001 relative to CTR. NS, not significant. Scale bars, 100μm in (**d**) and 25μm in (**f**) and (**h**).

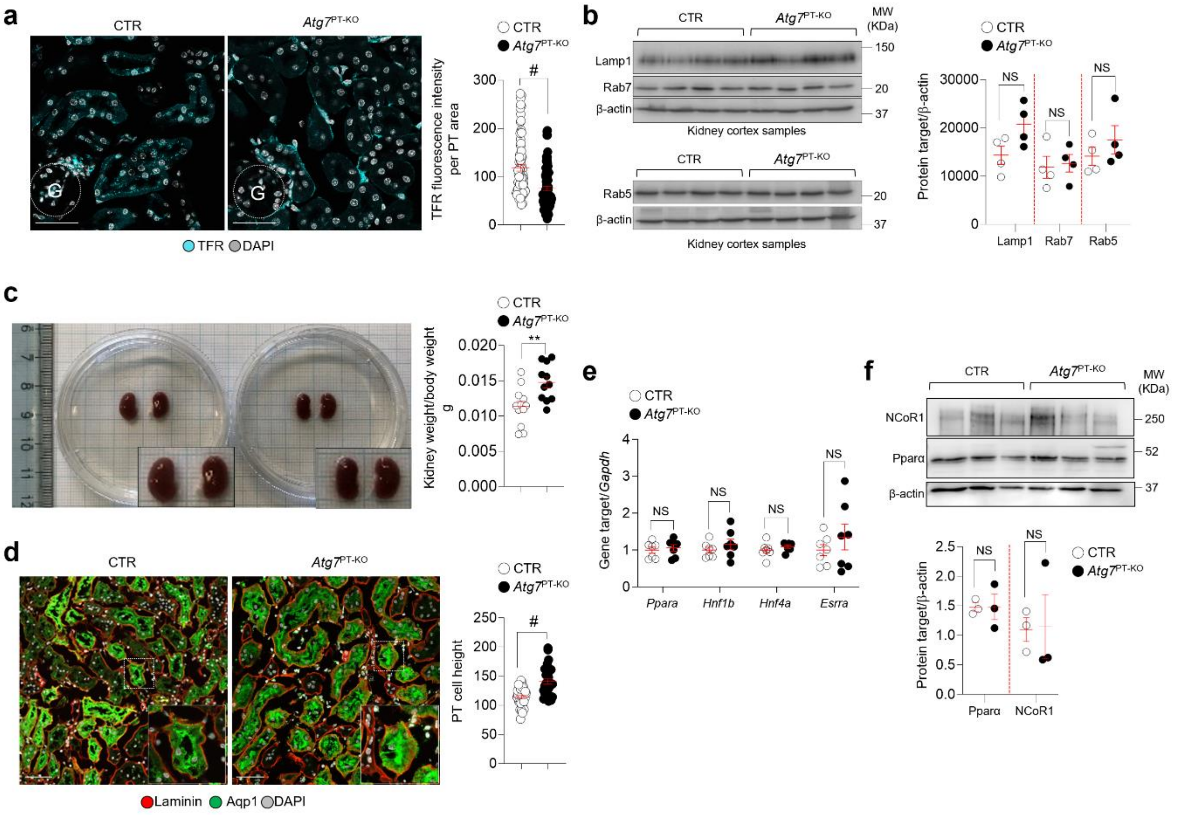


**Supplementary Fig. 4. Endocytosis, differentiation, and metabolism in the kidney PT of *Atg7* mice.** (**a**) Representative micrographs and quantification of transferrin receptor (TFR, cyan) fluorescence intensity in the kidney PT of CTR and $Atg7^{PT\text{-}KO}$ mice; n = 77 (CTR) or 83 ($Atg7^{PT\text{-}KO}$) PT segments pooled from 2 biologically independent experiments. (**b**) Immunoblots and quantification of the indicated proteins; n = 4 biologically independent animals per genotype. (**c**) Representative images and measurements of kidney-weight-to-body weight ratio in *Atg7* mice; n = 12 CTR and n = 11 $Atg7^{PT\text{-}KO}$ animals. (**d**) Whole-kidney sections were immunostained for apical (Aqp1, green) and basolateral markers (Laminin, red). Representative images and quantification of PT cell height in *Atg7* mice; n = 60 PT segments pooled from three biologically independent experiments. (**e**) Transcript levels of the indicated genes in the kidney cortex of 24-week-old mice; n = 7 biologically independent experiments. G: Glomerulus. Plots represent mean ± SEM. Statistical analysis was performed using an unpaired two-tailed Student's t-test, *P < 0.05, **P < 0.01, ***P < 0.001, and $^{\#}$P<0.0001 relative to CTR. NS, not significant. Nuclei (grey) were counterstained with DAPI. Scale bars, 25μm (**a**) and 50μm in (**d**).

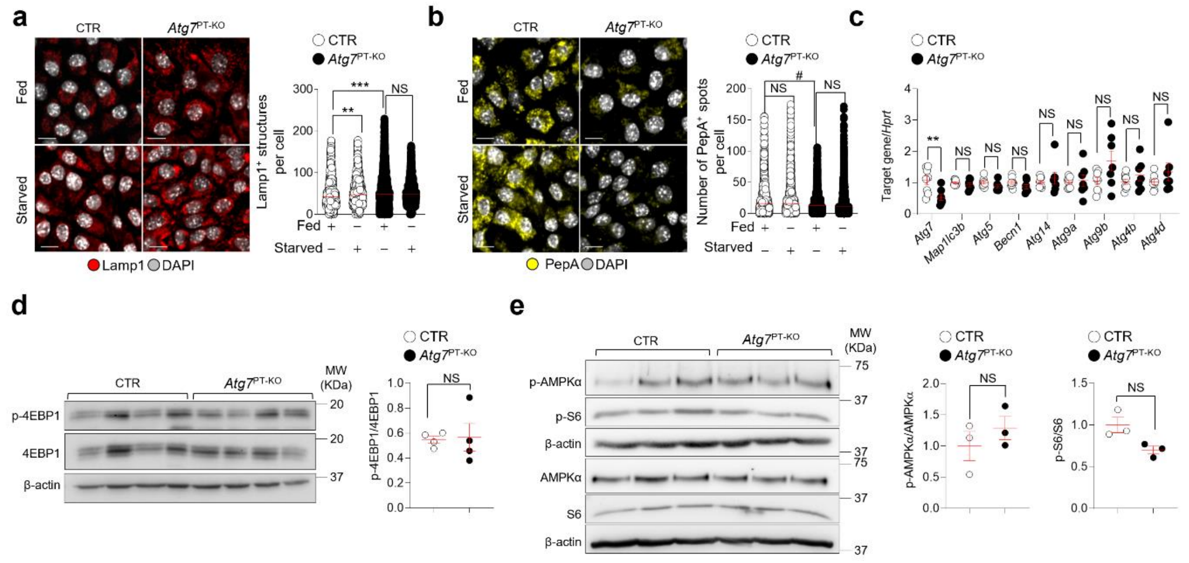


**Supplementary Fig. 5. Lysosome catabolism and nutrient sensing pathways in ATG7-deficient PT cells.** (**a-b**) CTR or *Atg7*$^{PT\text{-}KO}$ cells were cultured under fed or starved conditions for 8 hours, fixed and immunostained for Lamp1 (red) or stained with Pepstatin A (PepA, yellow), and analysed by confocal microscopy. Representative micrographs and quantification of the number of (**a**) Lamp1$^{+}$ and (**b**) PepA$^{+}$ structures per cell. For Lamp1$^{+}$ number: n = 1662 for CTR and 2073 for *Atg7*$^{PT\text{-}KO}$ in fed cells, and n = 2269 CTR and 1864 *Atg7*$^{PT\text{-}KO}$ in starved cells. For PepA$^{+}$ structures, n = 2404 CTR and 2467 *Atg7*$^{PT\text{-}KO}$ in fed cells, and n = 2925 CTR and 2434 *Atg7*$^{PT\text{-}KO}$ in starved cells, pooled from 4 biologically independent experiments. (**c**) mRNA levels of the indicated genes in *Atg7* mPTCs, n = 8 CTR and 7 *Atg7*$^{PT\text{-}KO}$ biologically independent samples. (**d-e**) Immunoblots and quantification of the indicated proteins, n = 3-4 biologically independent samples. The plots represent mean ± SEM, and statistical analysis was performed using a two-tailed unpaired Student's t-test, **P < 0.01, ***P < 0.001, and $^{\#}$P < 0.0001 relative to CTR cultured under fed conditions. Nuclei counterstained with DAPI (grey). NS, not significant. Scale bars, 25μm in (**a**) and (**b**).

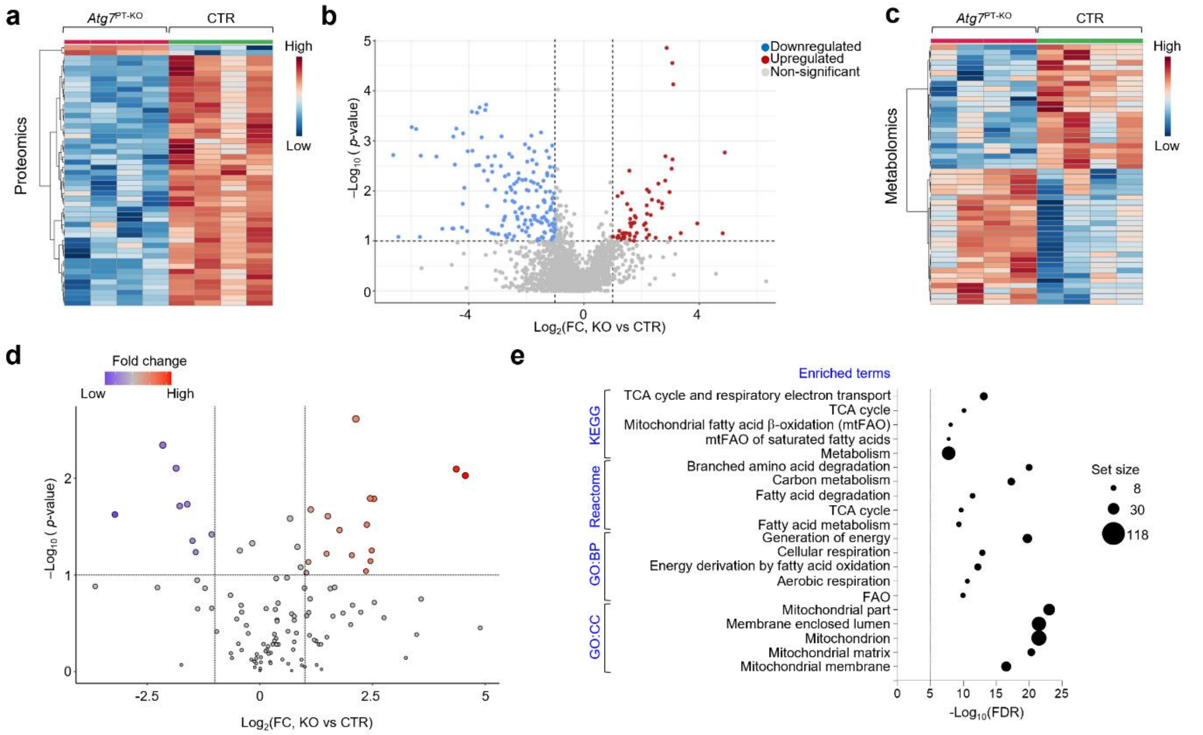


**Supplementary Fig. 6. Proteome and metabolome profiles of Atg7$^{PT-KO}$ vs. wild-type PT cells.** (**a**,**c**) Heat maps illustrating the abundance of the top 50 differentially expressed proteins (**a**) and metabolites (**c**) in *Atg7* mPTCs derived from kidneys of 24-week-old mice; n = 4 biologically independent experiments. (**b**,**d**) Volcano plots depicting the changes in (**b**) the proteome and (**d**) the metabolome in *Atg7*$^{PT-KO}$ vs CTR mPTCs. (**E**) Over-representation analysis of the top biological pathways — specifically Gene Ontology (GO) terms related to biological processes and cellular components, and pathways from Reactome and Kyoto Encyclopedia of Genes and Genomes (KEGG) databases. The analysis focused on differentially expressed proteins exhibiting a $\log_2$ fold change ($\log_2$FC) of less than -0.5 or greater than 1, with a P value of less than 0.1. This analysis utilised the Network Analyst toolkit (www.networkanalyst.ca), a web-based platform designed for biological network analysis that visualises genes and proteins within their respective biological contexts to investigate their statistical and functional relationships. The computational tool incorporates genes and proteins from 15 different databases to construct biological networks. These networks were analysed to identify genes, proteins, miRNAs, drugs, chemicals, or disease phenotypes with the most significant associations to the input list of genes (seed genes). The black dashed line represents the threshold for significant enrichment, while the size of the black circles indicates the enrichment score. P-values were derived from the enrichment pathway analysis performed in NetworkAnalyst.

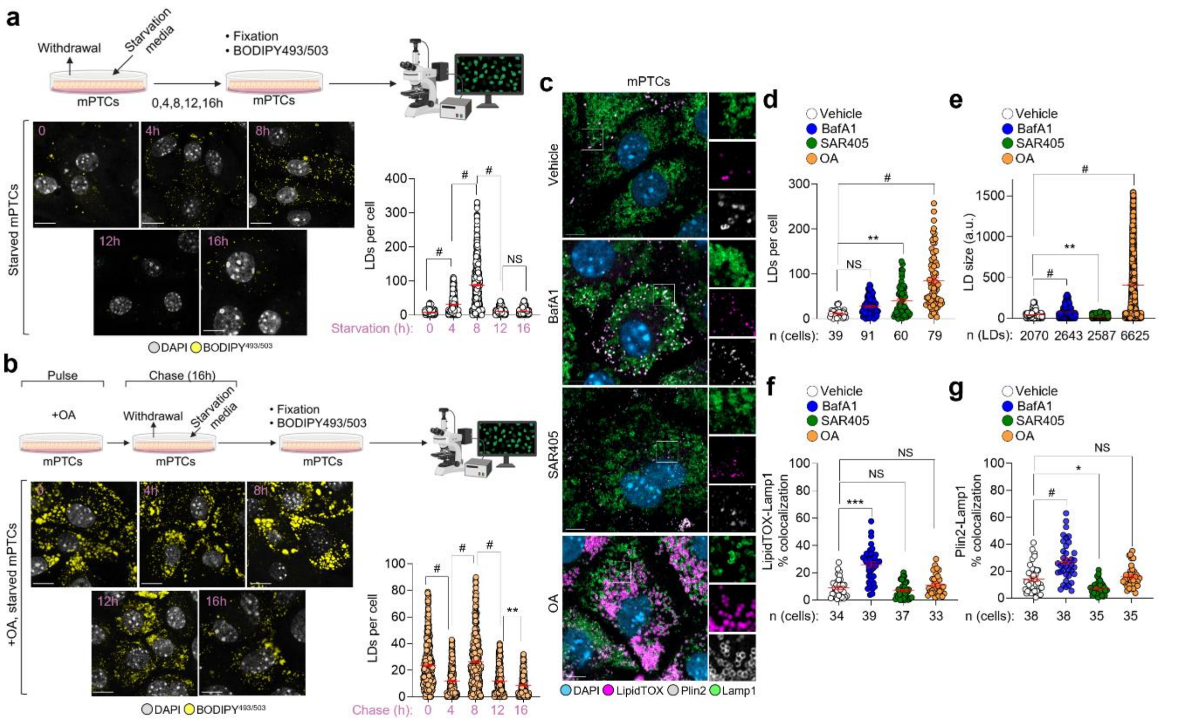


**Supplementary Fig. 7. Regulation of LD turnover in response to nutrient deprivation in cultured PT cells.** (**a**) Cells were starved for the indicated times, stained with BODIPY$^{493/503}$ (yellow), fixed, and analysed by confocal microscopy. Representative micrographs and quantification of the number of LDs per cell. Starved conditions: n = 234 cells (0h), 465 cells (4h), 470 cells (8h), 424 cells (12h), and 371 cells (16h). (**b**) Cells were pulsed with 300 µM oleic acid (OA) for 5h, chased for the indicated times (0,4,8,12 and 16 h) in starvation medium, stained with BODIPY$^{493/503}$ (yellow), fixed and analysed by confocal microscopy. Representative micrographs and quantification of the LD number per cell. OA-pulsed conditions: n = 459 cells (0h), 311 cells (4h), 442 cells (8h), 502 cells (12h), and 441 cells (16h). (**c**) mPTCs were pulsed with 300 µM OA for 24h or treated with 5 µM SAR405 for 16h or with 250 nM BafA1 for 4h, stained with LipidTOX (magenta), with anti-Lamp1 (green) and anti-Plin2 (grey), and analysed by confocal microscopy. Representative micrographs and quantification of (**d**) LD number and (**e**) size, and (**f**) LipidTOX/Lamp1 and (**g**) Plin2/Lamp1 colocalizations (%). Bar graphs show mean ± SEM. Statistics were calculated by one-way ANOVA followed by Tukey's multiple comparisons test, *P < 0.05, **P < 0.01, ***P < 0.001, $^{\#}$P < 0.0001 relative to vehicle-treated or starved cells (0h). Nuclei counterstained with DAPI (blue or grey). NS, not significant. Scale bars, 10µm in (**a**),(**b**), and (**c**).

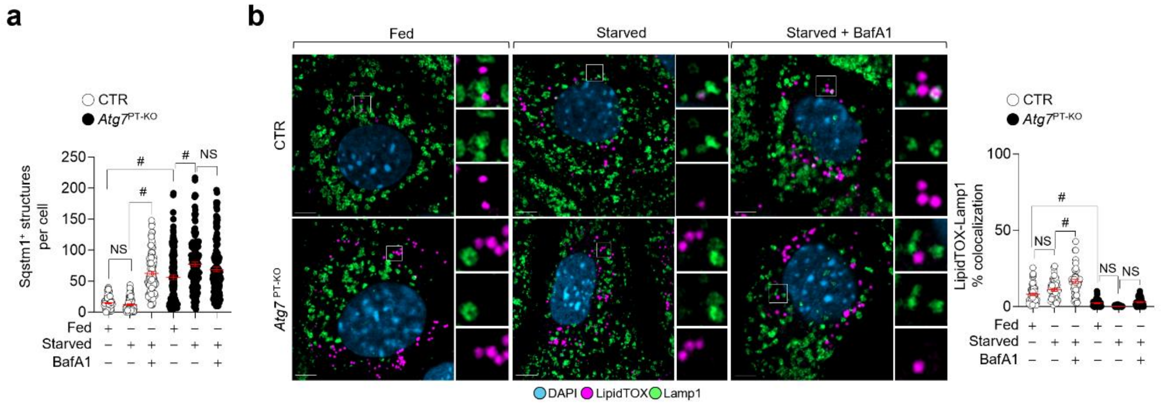


**Supplementary Fig. 8. LD turnover in ATG7-deficient mPTCs.** (**a**-**b**) Both CTR and *Atg7*$^{\text{PT-KO}}$ mPTCs were cultivated under fed or 16-hour starved conditions in the presence or absence of the lysosomal inhibitor BafA1 (250 nm for 4h). After treatment, the cells were fixed and stained with LipidTOX (magenta) and anti-Lamp1 (green) and analysed by confocal microscopy. (**a**) Quantification of the number of Sqstm1+ structures per cell (corresponding micrographs are shown in Fig. 4b) and (**b**) Representative micrographs and quantification of LipidTOX/Lamp1 colocalization (%). In (**a**): Number: n = 106 fed CTR, 108 starved CTR cells, 141 BafA1-treated, starved CTR cells, 141 fed *Atg7*$^{\text{PT-KO}}$, 148 starved *Atg7*$^{\text{PT-KO}}$, and 141 BafA1-treated, starved *Atg7*$^{\text{PT-KO}}$ cells. In (**b**): Colocalization (%): n = 48 fed and starved CTR, and 50 BafA1-treated, starved CTR; and n = 46 fed *Atg7*$^{\text{PT-KO}}$, starved *Atg7*$^{\text{PT-KO}}$, BafA1-treated, starved *Atg7*$^{\text{PT-KO}}$ cells. Bar graphs show mean ± SEM. Statistics were calculated using one-way ANOVA followed by Tukey's multiple comparisons test, $^{\#}P < 0.0001$ relative to fed/starved CTR or fed/starved *Atg7*$^{\text{PT-KO}}$ cells. Nuclei were counterstained with DAPI (blue). NS, not significant. Scale bars, 10µm.

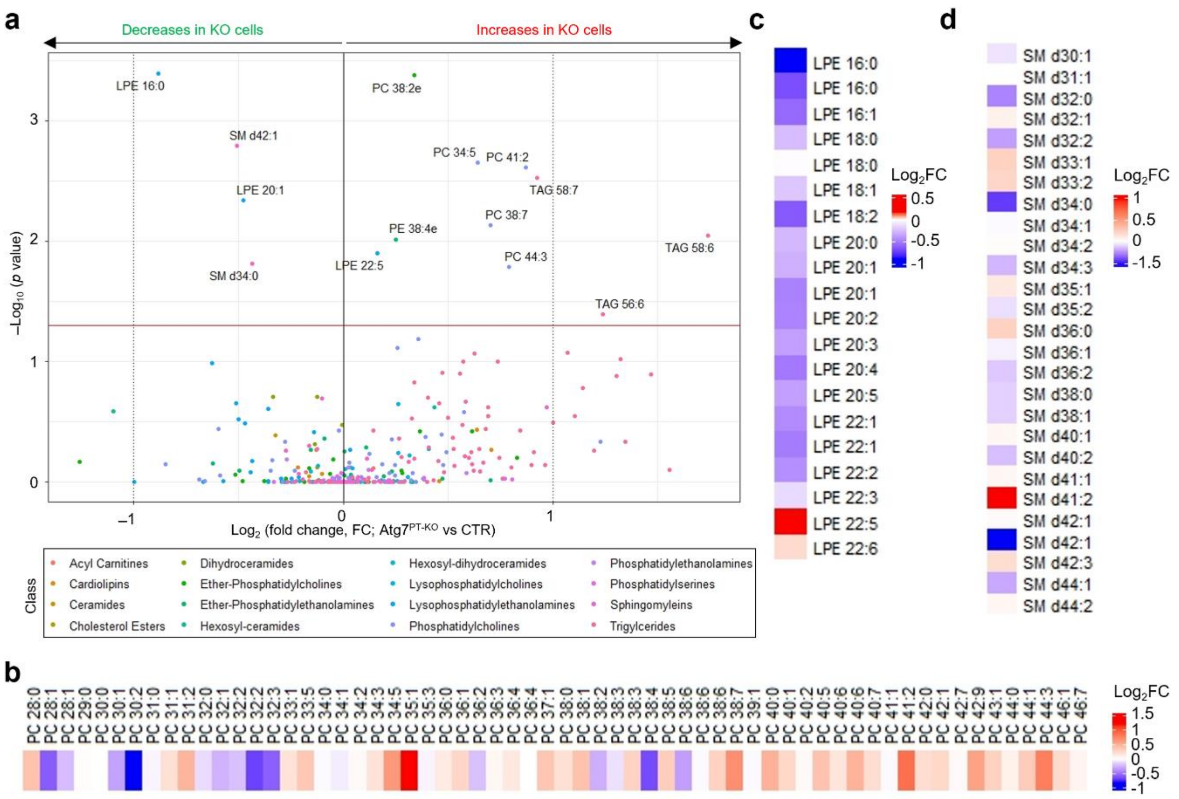


**Supplementary Fig. 9. Lipidomics-based profiling of PT cells deficient for ATG7 and autophagy.** (**a**) Volcano plot depicting the lipid changes resulting from ATG7 loss in PT cells; n = 4 biologically independent experiments (**b-d**) A heatmap illustrating the levels of b) phosphatidylcholine (PC), (**c**) lysophosphatidylethanolamine (LPE), and (**d**) sphingomyelin (SM), respectively.

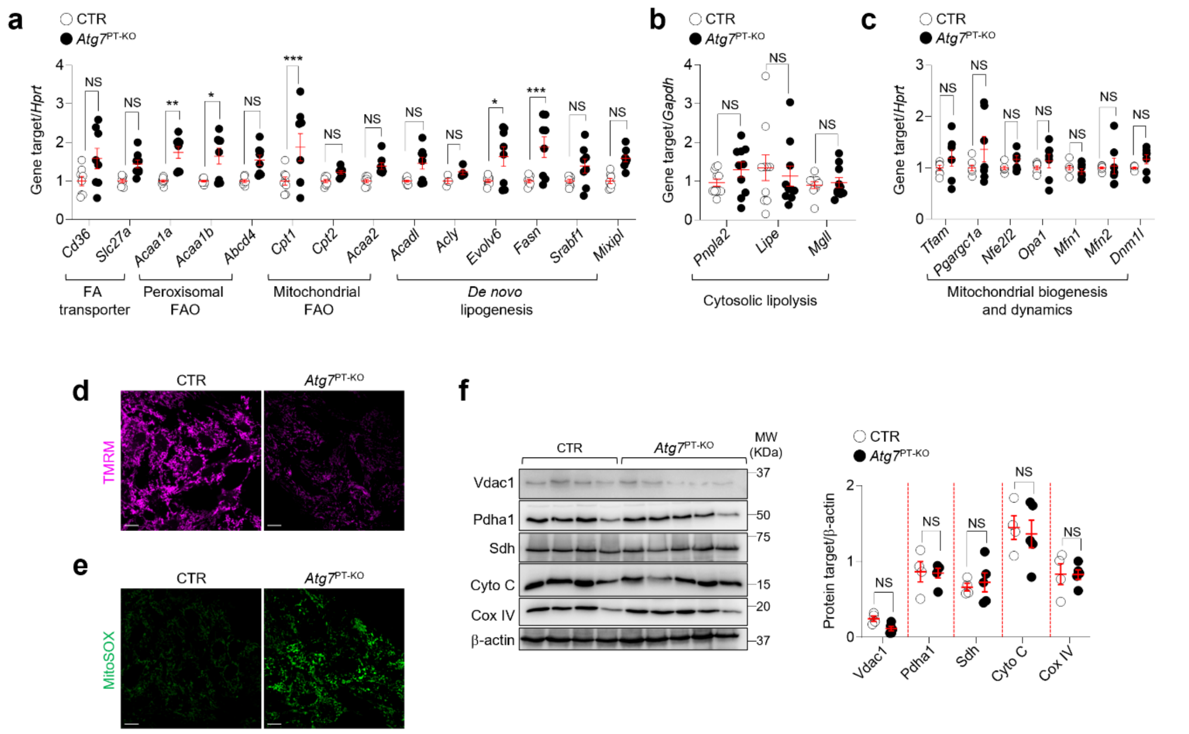


**Supplementary Fig. 10. Lipid metabolism gene signatures and mitochondrial mass in cultured PT cells** (**a-c**) mRNA levels of the indicated genes in CTR and *Atg7*$^{PT-KO}$ mPTCs, n = 7-9 replicates from two biologically independent experiments. (**d-e**) Representative micrographs of quantification shown in **Fig. 4g** and **4h**. (**f**) Immunoblot and quantification of the indicated proteins, n = 4-5 biologically independent experiments. The plots represent mean ± SEM. Statistical analysis was performed using unpaired two-tailed Student's t-test, *P < 0.05, **P < 0.01, and ***P < 0.001 relative to CTR cells or CTR mice. NS, not significant. Scale bars are 10μm in (**e**) and (**g**).

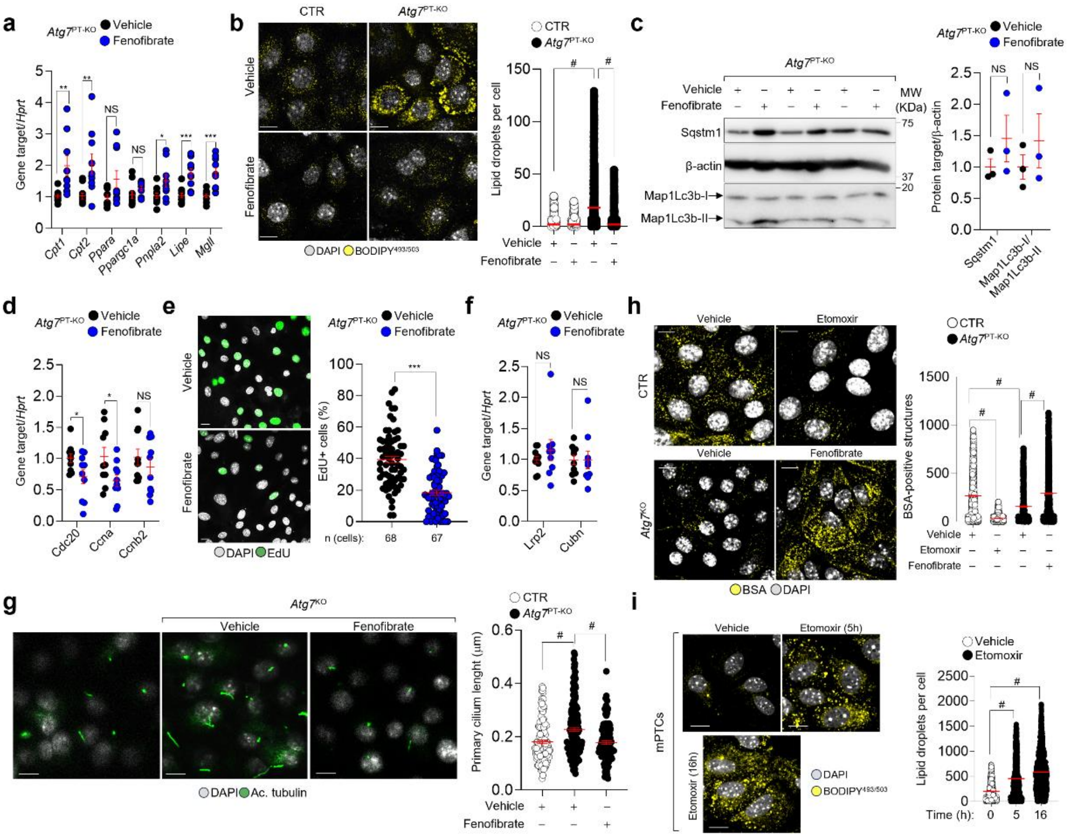


**Supplementary Fig. 11. Fenofibrate counteracts phenotypic changes induced by ATG7 loss in cultured PT cells.** (**a**-**g**) Both CTR and *Atg7*$^{PTKO}$ cells were treated with vehicle or fenofibrate (1μM for 16h). (**a**,**d**,**f**) mRNA levels of the indicated genes; n > 8 replicates pooled from 3 biologically independent experiments. (**b**) Cells were stained with BODIPY$^{493/503}$ (yellow), fixed and analysed by confocal microscopy. Representative micrographs and quantification for the number of LDs per cell; n = 4563 untreated CTR cells, 1643 fenofibrate-treated CTR cells, 4659 untreated *Atg7*$^{PT-KO}$ cells, and 3736 fenofibrate-treated *Atg7*$^{PT-KO}$ cells. (**c**) Representative immunoblot and quantification of the indicated proteins; n = 3 biologically independent experiments. (**e**) Representative micrographs and quantification of EdU$^{+}$ cells (%); n = 68 untreated and 67 fenofibrate-treated *Atg7*$^{PT-KO}$ cells. (**g**) Representative micrographs and quantification of BSA$^{+}$ structures per cell; n = 1970 untreated CTR cells, 1266 fenofibrate-treated CTR cells; and n = 1159 vehicle-treated *Atg7*$^{PT-KO}$ cells and 2741 fenofibrate-treated *Atg7*$^{PT-KO}$ cells. (**h**) Cells were fixed, stained with acetylated tubulin (ac-tubulin; green) analysed by confocal microscopy. Representative micrographs of the maximum intensity projection of image z-stacks and quantification of the primary cilium length; n = 128 vehicle-treated CTR cells, 197 vehicle-treated *Atg7*$^{PT-KO}$ cells, and 112 fenofibrate-treated *Atg7*$^{PT-KO}$ cells. (**i**) CTR cells were treated with vehicle or etomoxir for the indicated times, stained with BODIPY$^{493/503}$ (yellow), fixed, and analysed by confocal microscopy. Representative micrographs and quantification of the number of LDs; n = 808 vehicle-treated CTR cells, 571 etomoxir-treated CTR cells (5h), and 897 etomoxir-treated CTR cells (16h). Statistics were calculated using unpaired two-tailed Student's *t*-test in (**a**,**c**,**d-f**) and one-way ANOVA followed by Tukey's multiple comparisons test in (**b**,**g**,**h**,**i**), $^{\#}$P < 0.0001 relative to fed/starved CTR or *Atg7*$^{PT-KO}$ cells. Nuclei were counterstained with DAPI (blue or grey). NS, not significant. Scale bars, 10μm in (**b**,**e**,**g**,**i**).

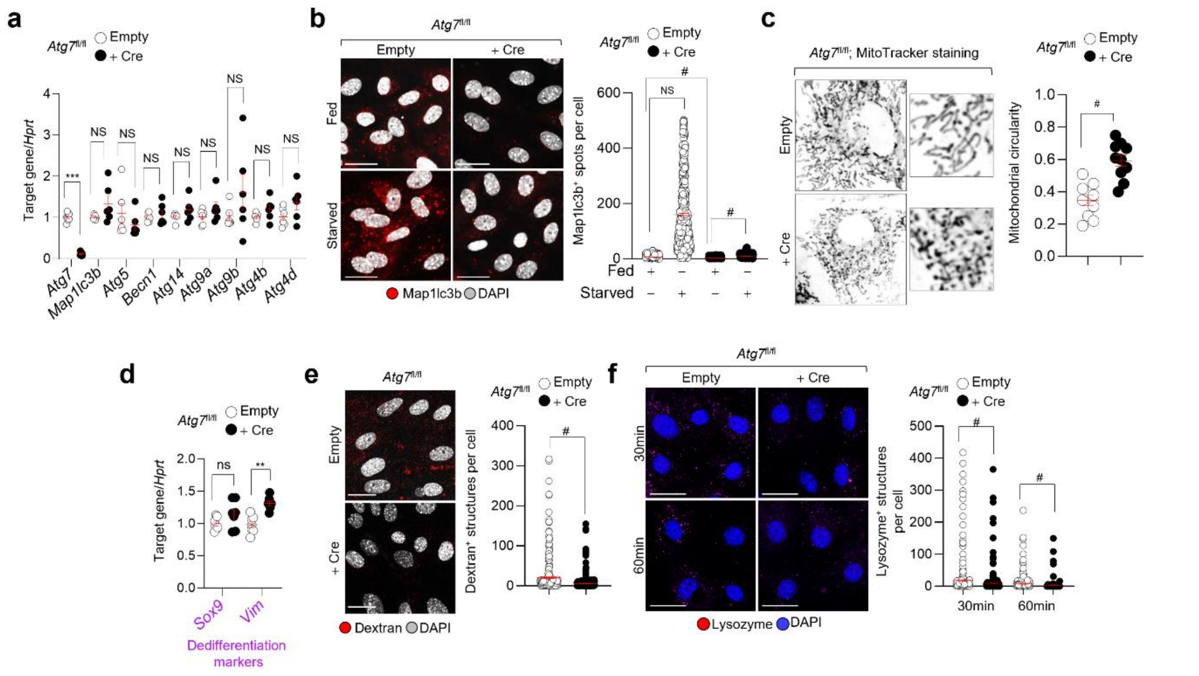


**Supplementary Fig. 12. Impaired autophagy and defective endocytosis in ATG7-deficient PT cells.** (**a**-**f**) mPTCs derived from microdissected PT of *Atg7* floxed kidneys were transduced with adenovirus bearing Empty or Cre-recombinase. (**a**) mRNA levels of the indicated genes; n = 6 Empty and n = 7 +Cre biologically independent experiments. (**b**) Representative micrographs and quantification of Map1lc3b$^+$ structures (red) in Empty and *Atg7*-deleted mPTCs cultured under fed and starved conditions; Fed conditions: n = 443 empty cells and 688 Cre-transduced cells. Starved conditions: n = 368 Empty cells and 465 Cre-transduced cells pooled from 2 biologically independent experiments. (**c**) Cells were stained with MitoTracker (1µM for 30 min at 37 °C) and analyzed by confocal microscopy. Representative micrographs and quantification of shape (expressed as circularity) of the mitochondrial network; each point represents the average values for circularity in a cell, n = 10 cells per condition, pooled from 2 biologically independent experiments. (**d**) mRNA levels of the indicated genes, n = 7 biologically independent experiments. (**e**) Cells were loaded with Alexa-647-Dextran (20µg × mL$^{-1}$ for min at 37 °C) and analyzed by confocal microscopy. Representative micrographs and quantification of Dextran$^+$ structures (red) in *Atg7* mPTCs; n = 358 Empty cells and 375 Cre-transduced cells pooled from 2 biologically independent animals. (**f**) The cells were loaded with Alexa-647- Lysozyme (40µg × mL$^{-1}$ for 30 and 60min at 37 °C) and analyzed by confocal microscopy. Representative micrographs and quantification of Lysozyme$^+$ structures (magenta); Timepoint, 30min: n = 315 Empty cells and 267 Cre-transduced cells. Timepoint, 60min: n = 240 Empty cells and 185 Cre-transduced cells pooled from 2 biologically independent animals. Plots represent mean ± SEM. Statistics were calculated using one-way ANOVA followed by Tukey's multiple comparisons test in (**b**,**f)** or by unpaired two-tailed Student's t-test in (**a**,**c**,**d**,**e)**; **P < 0.01; ***P < 0.001, and $^{\#}$P < 0.0001 relative to Empty cells cultured under fed or starved conditions, or to CTR cells. NS, not significant. Nuclei counterstained with DAPI (blue or grey). Scale bars, 10µm in (**b**,**f**,**g**).

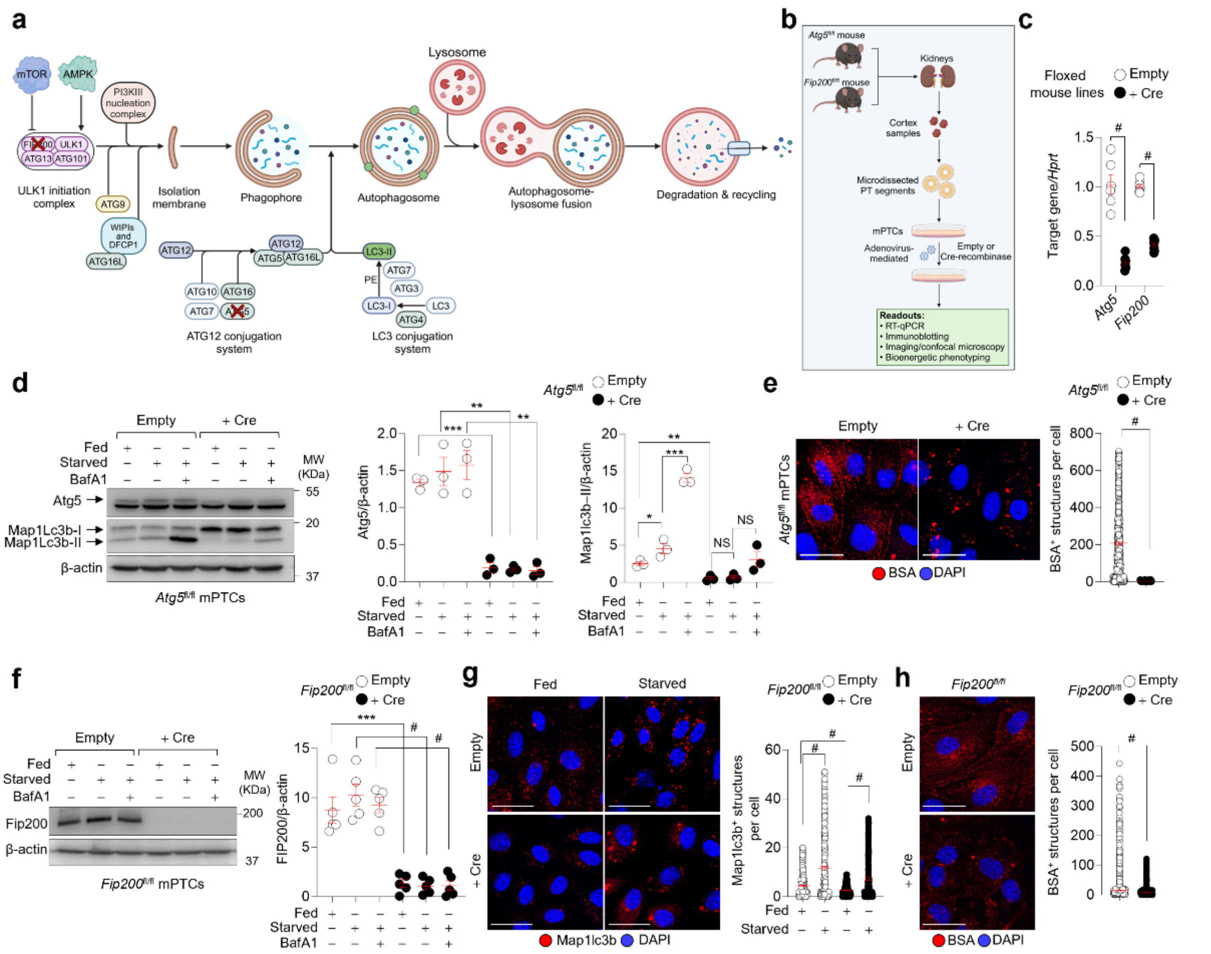


**Supplementary Fig. 13. Dysfunctional autophagy and altered endocytosis in FIP200- and in ATG5-deficient PT cells.** (**a**) Diagram illustrating the role of Unc-51-like kinase 1 (ULK1) complex that integrates upstream signals to initiate autophagy, leading to the formation of a phagophore, which expands through ATG5-ATG12 conjugates and ATG8 proteins (MapLc3b-II), eventually maturing into an autophagosome. (**b**) Experimental workflow for inactivating *Atg5* and *Fip200* in PT cells. (**c**) mRNA levels of the indicated genes in transduced *Atg5*$^{fl/fl}$ (n = 6 biologically independent experiments) and *Fip200*$^{fl/fl}$ mPTCs (n = 8 biologically independent experiments). (**d**,**f**,**g**) The transduced cells were cultivated under fed or starved conditions (8h), or starved conditions in the presence/absence of BafA1 (250 nM for 4h). (**d**,**f**) Immunoblot and quantification of indicated proteins; n = 3 (*Atg5*) and 5 (*Fip200*) biologically independent experiments. (**e**) Representative micrographs and quantification of BSA$^{+}$ structures; Empty: 539 cells and +Cre: n = 291 cells from 2 biologically independent experiments; each dot represents the number of BSA$^{+}$ structures in a cell. (**g**) Representative micrographs and quantification of Map1lc3b$^{+}$ structures (red); Fed conditions: Empty = 307 cells and +Cre = 338 cells. Starved conditions: Empty = 368 cells and Cre = 297 cells pooled from 2 biologically independent experiments. (**h**) Representative micrographs and quantification of BSA$^{+}$ structures in transduced *Fip200*$^{fl/fl}$ mPTCs; Empty = 1468 cells and +Cre = 1005 cells from 2 biologically independent experiments. Plots represent mean ± SEM. Statistics were calculated by an unpaired two-tailed Student's t-test in (**c**,**e**,**h**) or one-way ANOVA followed by Tukey's multiple comparisons test in (**d**,**f**,**g**), *P < 0.05; **P < 0.01; ***P <0.001, and $^{\#}$*P*< 0.0001 relative to Empty cells cultured under fed or starved conditions, or relative to +Cre cultured under fed conditions. Nuclei counterstained with DAPI (blue). Scale bars 25µm in (**e**,**g**,**h**).

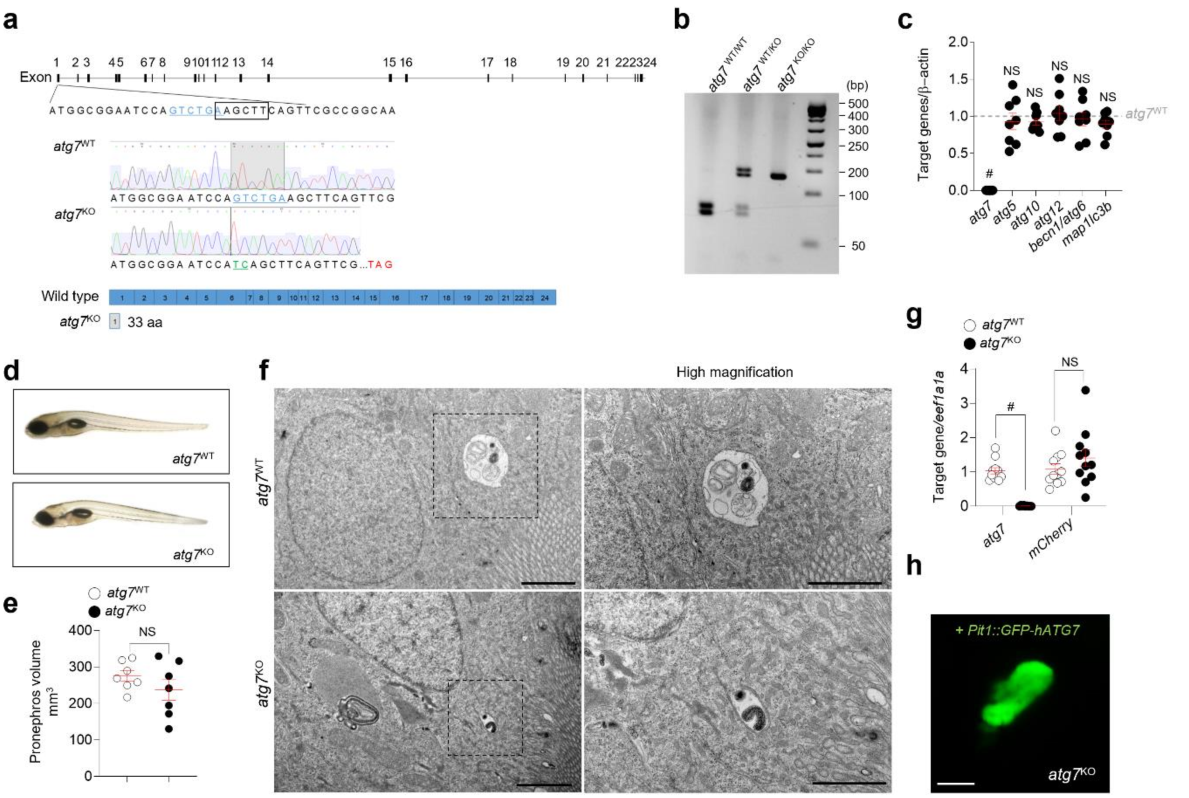


**Supplementary Fig. 14. Generation of the zebrafish model deficient for atg7.** (**a**) CRISPR-Cas9-induced deletion (blue) generates a frameshift in the open reading frame of the atg7 gene, resulting in a premature stop codon (TGA) within exon 1. This leads to the production of a truncated protein consisting of only 33 amino acids. (**b**) HindIII digestion of PCR products from the amplified CRISPR-Cas9 target region using genomic DNA extracted from the caudal region of wild-type (*atg7*$^{WT/WT}$), heterozygous (*atg7*$^{WT/KO}$) and homozygous (*atg7*$^{KO/KO}$) zebrafish larvae. The wild-type allele shows two lower bands (135 and 145 bp), representing PCR products cleaved by HindIII. The mutant allele shows a single upper band (276 bp), which is resistant to HindIII digestion. (**c**) mRNA levels for the indicated genes in *atg7* zebrafish larvae at 14dpf; n = 8 animals per genotype. (**d**) Representative images of *atg7*$^{WT}$ and *atg7*$^{KO}$ zebrafish larvae at 10dpf. (**e**) Quantification of pronephros volume (mm$^3$) in *atg7* zebrafish larvae at 5dpf; n = 7 zebrafish larvae per genotype. (**f**) Representative electron microscopy of pronephros from *atg7* zebrafish larvae at 14dpf. Insets show high magnification of the indicated area (black squares). (**g**) mRNA levels for the indicated genes in *atg7* zebrafish at 14dpf; n = 12 animals per genotype. (**h**) Light sheet microscopy and representative micrograph of Pit1::GFP-hATG7 expression in the pronephros of *atg7*$^{KO}$ zebrafish larvae at 14dpf. Plots represent mean ± SEM. Statistics calculated by two-tailed unpaired Student's t-test, $^{\#}P$ <0.0001 relative to *atg7*$^{WT}$. NS, not significant. Scale bars, 10μm (top panel) and 2μm (bottom panels) in (**f**), and 50μm in (**h**).

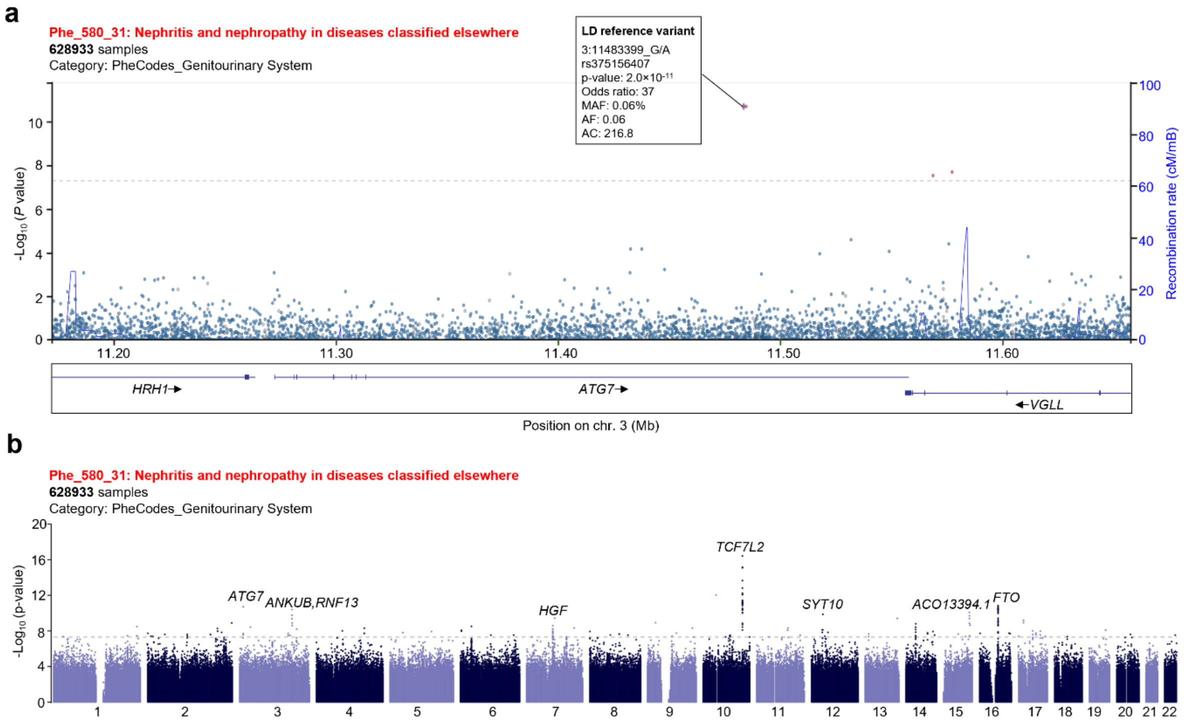


**Supplementary Fig. 15. Interactive views of associations between ATG7 and traits and diseases.** (**a**) Regional view (LocusZoom) for variant rs375156407 (purple diamond), with nearby variants present in the NHGRI-EBI GWAS Catalogue displayed. (**b**) Manhattan-plot view depicting GWAS results for “nephritis and nephropathy in diseases classified elsewhere” derived from 628993 samples in the VA Million Veteran (MVP) Program Biobank. The grey line shows the genome-wide significance threshold ($5 \times 10^{-8}$). Chromosome and position are displayed on the x-axis, and the two-sided −log10 P value is on the y-axis.

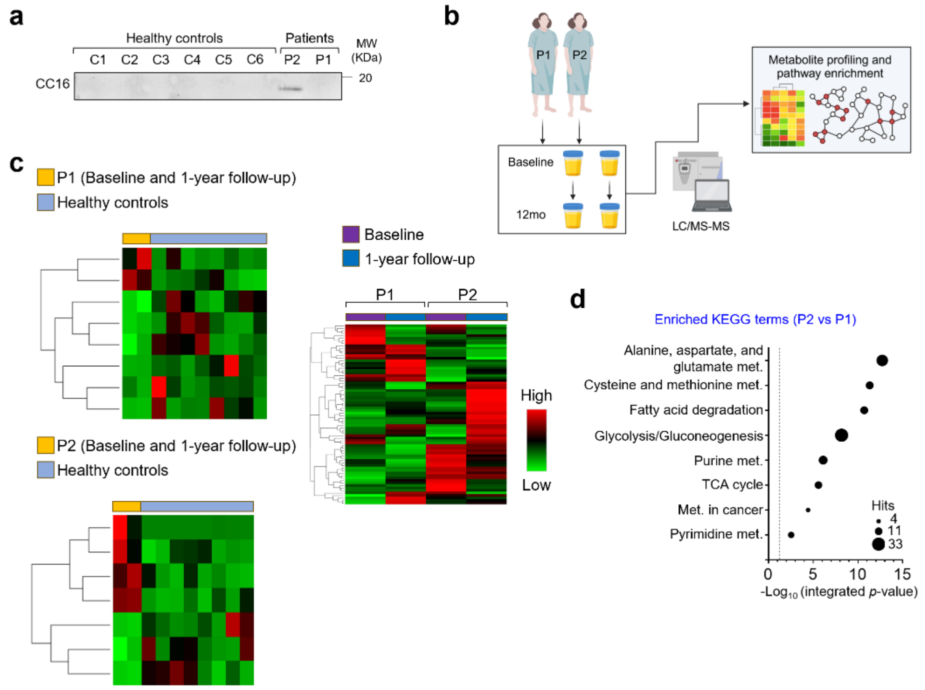


**Supplementary Fig. 16. Phenotypic changes in two female siblings with recessive pathogenic *ATG7* variants.** (**a**) Immunoblots of the low-molecular-weight protein CC16 in the urine samples of healthy subjects (n = 6) and two siblings with deleterious *ATG7* variants (n = 2). (**b**) Schematic of the experimental workflow for untargeted metabolomics and pathway enrichment analysis. (**c**) Heatmaps illustrate the levels of the top metabolites in P1 versus healthy controls, in P2 versus healthy controls, and in P1 versus P2. (**d**) Metabolite profiling and pathway enrichment analysis. The Black dashed line shows the threshold of significant enrichment. Black circles show the size of enriched proteins and metabolites, respectively. P-value was calculated from the enrichment analysis using Omicsnet2.0.

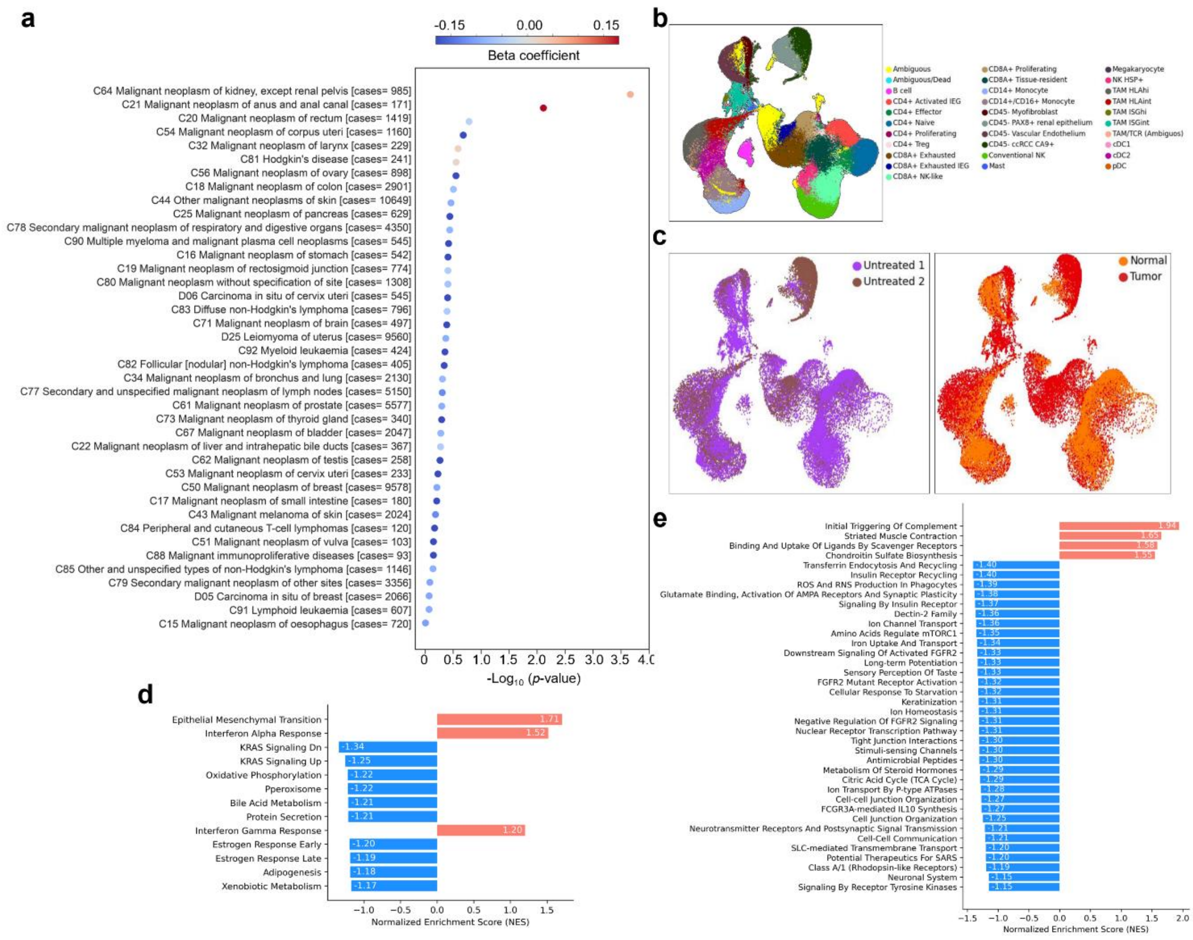


**Supplementary Fig. 17. Association of *ATG7* genetic variation with kidney neoplasms and pathway enrichment in single-cell transcriptional profiles of two patients with ccRCC.** (**a**) Burden test assessing the association between predicted loss-of-function (pLOF) alleles in *ATG7* and the prevalence of neoplasms. Beta coefficients and p-values are shown for the burden test linking Chapter II Neoplasms (ICD10 codes with case numbers in brackets) to *ATG7* pLOF, based on Genebass data. (**b**) UMAP embedding of transcriptional profiles from all patients and samples, with each dot representing a single cell. Colours denote clusters corresponding to inferred cell types. (**c**) UMAP embedding of single-cell transcriptional profiles from two patients with ccRCC (Untreated 1 and 2) across sample types (Normal and Tumour). (**d**-**e**) Pathway enrichment analysis of the PAX8-positive renal epithelial cell gene signature from Untreated Patient 2, identifying up- and down-regulated pathways, according to Reactome and other biological pathway databases.

**Supplementary Table 1:** Clinical and biochemical parameters in *Atg7*$^{PT\text{-}KO}$ mice

| | 4 weeks | | 12 weeks | | 24 weeks | |
|---|---|---|---|---|---|---|
| **Parameters** | CTR (n=10 ♂ and 10 ♀) | *Atg7*$^{PT\text{-}KO}$ (n=10 ♂ and 10 ♀) | CTR (n=10 ♂ and 10 ♀) | *Atg7*$^{PT\text{-}KO}$ (n=10 ♂ and 10 ♀) | CTR (n=10 ♂ and 10 ♀) | *Atg7*$^{PT\text{-}KO}$ (n=10 ♂ and 10 ♀) |
| Body weight (BW), g | 18.96 ± 0.57 | 17.63 ± 0.66 | 27.90 ± 0.95 | 21.93 ± 08 2$^{\#}$ | 37.10 ± 1.51 | 27.82 ± 1.53$^{\#}$ |
| **Urine** | | | | | | |
| Diuresis, μL·16h$^{-1}$·g BW$^{-1}$ | 3.01 ± 0.28 | 4.78 ± 0.45** | 3.00 ± 0.24 | 3.02 ± 0.24 | 1.74 ± 0.15 | 2.81 ± 0.31** |
| Creatinine, mg·dL$^{-1}$ | 39.23 ± 3.53 | 41.91 ± 4.49 | 24.11 ± 1.38 | 26.38 ± 1.81 | 23.26 ± 2.0 | 23.29 ± 1.94 |
| CC16, mg·g creat$^{-1}$ | 0.12 ± 0.02 | 0.63 ± 0.14$^{\#}$ | 0.03 ± 0.01 | 0.33 ± 0.06$^{\#}$ | 0.07 ± 0.01 | 0.22 ± 0.03$^{\#}$ |
| Albumin, mg·g creat$^{-1}$ | 5.13 ± 0.58 | 13.16 ± 1.85*** | 8.44± 0.63 | 25.51± 4.34*** | 20.63 ± 2.63 | 35.53± 3.64** |
| Glucose, g·g creat$^{-1}$ | 0.53 ± 0.07 | 0.91 ± 0.16 | 0.36 ±0.04 | 0.30 ± 0.03 | 0.14 ± 0.02 | 0.29 ± 0.05* |
| $Na^+$, g·g creat$^{-1}$ | 5.04 ± 0.26 | 5.42 ± 0.23 | 5.99 ± 0.37 | 6.37 ± 0.29 | 5.85 ± 0.29 | 6.97 ± 0.32* |
| $K^+$, g·g creat$^{-1}$ | 19.78 ± 0.48 | 19.87 ± 0.63 | 17.14 ± 0.78 | 18.62 ± 0.79 | 19.00 ± 0.95 | 24.00± 1.53** |
| $Cl^-$, g·g creat$^{-1}$ | 13.12 ± 0.37 | 14.08 ± 0.31 | 11.11 ± 0.73 | 11.138 ± 0.65* | 11.39 ± 0.67 | 15.72 ± 1.29** |
| $Ca^{2+}$, g·g creat$^{-1}$ | 165.76 ± 14.86 | 140.77 ± 17.29 | 136.46 ± 12.16 | 182.33 ± 16.90** | 216.92 ± 25.57 | 258.55 ± 41.22 |
| $P^{3-}$ , g·g creat$^{-1}$ | 2.32 ± 0.24 | 2.04 ± 0.20 | 3.64 ± 0.22 | 3.09 ± 0.35 | 2.24± 0.31 | 3.33 ± 0.34* |

Measurements were performed on *Atg7*$^{PT\text{-}KO}$ and CTR mice matched per gender and age. Two-tailed unpaired Student's t-test was applied between genotypes at the indicated time points. *$P < 0.05$, **$P < 0.01$, ***$P < 0.001$, $^{\#}P < 0.0001$ relative to CTR mice. n, number of mice; creat, creatinine.

**Supplementary Table 2:** Kidney function markers in 24-week-old *Atg7*$^{PT\text{-}KO}$ mice

| | 24 weeks | |
|---|---|---|
| **Plasma parameters** | CTR (n=8 ♂ and 5 ♀) | *Atg7*$^{PT\text{-}KO}$ (n=9 ♂ and 4 ♀) |
| Creatinine, mg·dL$^{-1}$ | 0.12 ± 0.01 | 0.15 ± 0.01 |
| BUN, mg·dL$^{-1}$ | 21.37 ± 1.22 | 22.09 ± 1.20 |

Measurements were performed on age- and gender-matched CTR and *Atg7*$^{PT\text{-}KO}$mice.
n, number of animals; BUN, Blood urea nitrogen.

**Supplementary Table 3**: Primer pairs for gene expression analysis in murine cells and kidneys

| Gene product | Forward primer (5'-3') | Reverse primer (5'-3') |
|---|---|---|
| *Atg7* | GCT AAT GGA CAC CAG GGA GA | CAG TC AGC AGG TGC TAC AA |
| *γGt1* | ACC TGT CTG CGG TTT CAG AG | TGA TGA AGT TGG CTG GTG AG |
| *Hprt* | ACA TTG TGG CCC TCT GTG TG | TTA TGT CCC CCG TTG ACT GA |
| *Map1lc3b* | CCG AGA AGA CCT TCA AGC AG | CCA GGA ACT TGG TCT TCT CC |
| *Atg5* | GAC CTT CTA CAC TGT CCA TCC A | TCA TTC TGC AGT CCC ATC C |
| *Fip200* | GAT GCC TGC AGT TTT CCA CA | CCA GAT TGG CCA TGA CA |
| *Becn1* | AGG AGC TGG AAG ATG TGG AA | ACT CCA GCT GCT GCC TTT TA |
| *Atg14* | AGA AGA TTC AGC GGC ACA AC | TCA CTT CGT CGA TTG GGA AT |
| *Atg9a* | CTC CGA GTG ATT CTT GCA CA | GAC GAT GGG ACT CAG CAA CT |
| *Atg9b* | CAC ATG CAT TAC CTC CCA GA | AGC AGA AAC AAT GGG GTC AG |
| *Atg4b* | CAG ATA GCG CAA ATG GGA GT | CCA TCA CCA CAG TGT TGT CC |
| *Atg4d* | TGC TAC CAT TTC GAG GGA GA | ATA CAT CCC CAG CCA CAG TC |
| *Lrp2* | CAG TGG ATT GGG TAG CAG | GCT TGG GGT CAA CAA CGA TA |
| *Cubn* | TCA TTG GCC TCA GAC ATT CC | CCC AGA CCT TCA CAA AGC TG |
| *Pcna* | TTG GAA TCC CAG AAC AGG AG | ATT GCC AAG CTC TCC ACT TG |
| *K167* | TGC AAA GGT AGA GGC TCC AT | CAG GTA GGC CAG AGC AA GT |
| *Scl34a1* | GGG AGA AGC TAT CCA GCT CA | ACA GCA AAC CAG CGG TAC TT |
| *Slc5a2* | TTG GGC ATC ACC ATG ATT TA | GCT CCC AGG TAT TTG TCG AA |
| *Ccna2* | CTT GGC TGC ACC AAC AGT AA | AGC AAT GAG TGA AGG CAG GT |
| *Cdc20* | ATG GAG CAG CCT GGA GAC TA | GCT TAC TCG AGC GGA GTG AC |
| *Ccnb2* | TGA AAC CAG TGC AGA TGG AG | CTG CAG AGC TGA GGG TTC TC |
| *Cpt1* | TGG CAG TCG ACT CAC CTT TC | ACA CCA TAG CCG TCA GC |
| *Cpt2* | TTG ACG CCA TTC AGT TTC AG | GCA GTG CTG CAG GAT TCA TA |
| *Sox9* | CAA GAA CAA GCC ACA CGT CA | GTG GTC TTT CTT GTG CTG CA |
| *Vim* | AAT GCT TCT CTG GCA CGT CT | AGT GAG GTC AGG CTT GGA AA |
| *Gal3* | GCC TAC CCC AGT GCT CCT | TTG CGT TGG GTT TCA CTG TG |
| *Tlr4* | GTG GCC CTA CCA AGT CTC AG | GAC CCA TGA AAT TGG CAC TC |
| *Tgfb1* | GTG GAA ATC AAC GGG ATC AG | GTT GGT ATC CAG GGC TCT C |
| *Adgre1* | CCA GGA GTG GCT TTT GTC TC | GGC TTG GAG AAG TCC TCC TT |

| Gene product | Forward primer (5'-3') | Reverse primer (5'-3') |
|---|---|---|
| *Fn1* | GCA AGC CAG TTT CCA TCA AT | CAT TTT TGG GAG TGG TGG TC |
| *Gapdh* | TGC ACC ACC AAC TGC TTA GC | GGA TGC AGG GAT GAT GTT CT |
| *Pcna* | TTG GAA TCC CAG AAC AGG AG | ATT GCC AAG CTC TCC ACT TG |
| *Pnpla2* | CAA CGC CAC TCA CAT CTA CG | ACC AG GTTG AAG GAG GGA TG |
| *Ppara* | ACT GCC GTT TTC ACA AGT GC | CGT GGA TTC TCT TGC CCA GA |
| *Ppargc1a* | TGG ATG AAG ACG GAT TGC CC | TGC TAA GAC CGC TGC ATT CA |
| *Lipe* | TGA GAT TGA GGT GCT GTC GT | AGT AC CTTG CTG TCC TGT CC |
| *Mgll* | AGT GAG GGA GAG AGG ATG GT | AGT CGA TGC AGA TTC CGG AT |
| *Hnf1b* | AGA TCA CAG TGT CGG GAG GA | GAG GTG TGG AGG CTC TGT G |
| *Hnf4a* | TGC GAA CTC CTT CTG GAT GAC C | CAG CAC GTC CTT AAA CAC CAT GG |
| *Esrra* | GGA AAG TGA ATG CCC AGG TG | CAG CAA AAG CAT CAC CCA GT |
| *CD36* | ACT GTG GGC TCA TTG CTG G | TGA TTT TGC TGC TGT TCT TTG C |
| *Slc27a* | GCT CCT GCG GCT TCA ACA | GCG CTA TCG CCC TTT CG |
| *Acaa1a* | GAC ATT GGC ATG GCC TGT GG | GCT TCT GCC GTG AAA TGC CA |
| *Acaa1b* | CTG CTT CAA GGA CAC CAC CC | TGC CTG CAG TCC CGA TGA AC |
| *Acaa2* | ACC CTT TGG AGC TTA CGG GG | TCC CAC TCG CAA ACC CAC AT |
| *Acadl* | ATG ACG GCG GGC AAG TGT AT | TCG AGC TTC ACG GTT GGT GA |
| *Abcd4* | GAG GAA GGA CCT CAC GGA GC | GCC AGC CTG TGC TTT GGA AG |
| *Acyl* | CTC ATT GAA CCC TTC GTC CC | CCT CGG TAT TCA GCT TTT CGT |
| *Evolv6* | GAC TCC GAA GAT CAG CCC CA | AAG ACG GCA AGA GTC AGC GA |
| *Fasn* | CTG CCT CCG TGG ACC TTA TC | GCA CAG ACA CCT TCC CGT CA |
| *Srabf1* | TTC TGG AGA CAT CGC AAA CAA | TGG TAG ACA ACA GCC GCA TC |
| *Mixipl* | GCT GCG GGA TGA AAT AGA GG | TCA AAT AAA GGT CGG ATG AGG A |
| *Tfam* | TAG GAA AAT TGC AGC CCT GT | CCT TCT CCA TAC CCA TGA GG |
| *Nfe2l2* | CAG CCA GCT GAC CTC CTT AG | TCA ATA GTC CCG TCC AGG AG |
| *Opa1* | GGA CCC AAG AGC AG GTG TT | TGG ATC GAC TTC CAC TCC TC |
| *Mfn1* | TGA TCT CAA TTG CCA CAA GC | ACC TGA AAG ATG GGC TCT GA |
| *Mfn2* | TGC AGA CTA TGC AGC AGG AC | CCT GAA AGT CAG CAC ACA GC |
| *Dnm1l* | TCA GAT CGT CGT AGT GGG AA | TCT TCT GGT GAA ACG TGG AC |

**Supplementary Table 4:** Primer pairs for gene expression analysis in zebrafish

| Gene product | Forward primer (5'-3') | Reverse primer (5'-3') |
|---|---|---|
| *atg7* | GGC GGA ATC CAG TCT GAA GC | CAG ACC AAC AGC ATC ACC GTT |
| *atg5* | CCC TAC TAT CTG CTC CTC CCA | CAA ACC ACA TTT CCT CCA CAT CC |
| *atg6/becn1* | ATG GTG GCT TTC CTT GAC TG | CTC CTG TGT CCT CAA TCT TTC |
| *atg10* | GAA GAA GAC TGT GCT CAT CCC T | ACT CCA GCC GTT TCA TCC TC |
| *atg12* | TCA CTC GCC CAG TTC ATC TC | CCG TCA CTT CCG AAA CAC TC |
| *map1lc3b* | ACT AAG TTT CTA GTT CCT GAC CAC | AGA TAC CAT GCT GTG ACC GT |
| *mCherry* | CCA TGT GAG AAT GGC TGC TA | ACC TTG AAG CGC ATG AAC TC |
| *β-actin* | TGA ATC CCA AAG CCA ACA GAG | TCA CAC CAT CAC CAG AGT CC |
| *eef1a1a* | TTC TCC GAG TAT CCT CCT CTG | CTT CTC CAC TCC TTT AAT CAC TCC |
